# When the Glass is Half Full: Early Life Experiences and Adult Optimism in 22 Countries

**DOI:** 10.1101/2025.03.14.25323965

**Authors:** Ying Chen, Laura D. Kubzansky, Eric S. Kim, Hayami Koga, Koichiro Shiba, R. Noah Padgett, Renae Wilkinson, Byron R. Johnson, Tyler J. VanderWeele

## Abstract

Little is known about early-life experiences that may lead to higher optimism levels in adulthood. Using data from 202,898 adults in 22 countries, we evaluated childhood candidate antecedents of optimism. We examined the associations between retrospectively reported childhood experiences and adult optimism levels in each country separately, and cross-nationally by pooling results across countries. Our pooled results suggest that higher adult optimism levels were associated with childhood experiences of having positive relationships with both parents, higher subjective financial status, better childhood self-rated health, frequent religious service attendance, an earlier year of birth, and being female. Conversely, lower adult optimism was associated with childhood experiences of parental divorce, abuse, financial hardship, and feeling like an outsider in the family. However, country-specific analyses showed substantial between-country variations in these associations, suggesting diverse societal influences. This study provides valuable insights into the association between early-life experiences and adult optimism levels across national contexts.

## INTRODUCTION

Governments worldwide are increasingly interested in incorporating indicators of well-being for assessing societal progress and policy making^1^. To this end, there has been increasing interest in identifying resources that may contribute to fostering well-being^2^. Dispositional optimism, defined as “a generalized expectation that good things will happen”^3^, is a potentially modifiable psychological attribute that has been linked with higher levels of multiple aspects of health and well-being^4^. However, social and structural conditions that may increase optimism are not well understood, with research on factors in childhood particularly limited.

Based on the Expectancy-Value Theory, optimistic individuals tend to perceive desired outcomes as attainable and are motivated to engage in goal-directed behaviors, leading to enhanced well-being^3^. Longitudinal evidence, mostly based on samples from the U.S. and European countries, suggests that more optimistic individuals, on average, have lower risks of mortality^5^, cardiovascular diseases^6^, and mental illness^7^, as well as healthier lifestyles^8^, when compared to their less optimistic counterparts. Given the benefits of optimism for mental and physical health, identifying factors that increase the likelihood individuals can maintain optimism over the life course may provide important targets for strategies that aim at improving population well-being.

One’s level of optimism typically remains stable in adulthood, and early life may be a crucial life period for cultivating an optimistic outlook^9^. While optimism is estimated to be approximately 25% heritable^10^, a number of individual-, interpersonal- and familial experience during early life may play a significant role in setting the developmental trajectory of optimism and ultimately adult levels of optimism. First, positive *family relationships* may help children foster positive expectations for their future. For instance, prior prospective studies have shown that experiencing parental warmth as well as an authoritative parenting style are each correlated with having higher optimism in later life, and that both maternal and paternal parenting practices contribute to fostering positive thinking in children^11,12^. Further, research has found that positive parent-child relationships help buffer the negative consequences of difficult circumstances such as parental divorce on the likelihood that children will be optimistic later in their lives^13^.

Relatedly, there is also evidence suggesting that experiencing a higher sense of family belonging is associated with greater well-being across family structures in early life^14^. Second, *adverse childhood experiences* may compromise the development of optimism. For instance, prior studies (mostly from Western countries) have suggested that experiencing child abuse is associated with lower optimism in adulthood^15,16^. Another form of childhood adversity is financial hardship, which prior work has shown is inversely associated with adult optimism, even after accounting for adult socioeconomic status (SES)^17,18^. Other difficult childhood psychosocial experiences, such as chronic stress related to one’s social identity (e.g., discrimination due to one’s race, ethnicity, or immigration status), can also negatively affect one’s outlook for the future and ultimately adult levels of optimism^19^. Third, optimism is a highly valued perspective among many religious communities^20^, and it is possible that having a *religious upbringing* may catalyze the cultivation of optimism. While few studies have directly examined linkages between childhood religious participation and adulthood optimism, some longitudinal evidence has found that a religious upbringing is positively associated with multiple facets of positive psychological functioning (e.g., happiness, purpose in life) that are closely related to optimism^21^. Fourth, *childhood health conditions* may also affect the likelihood of being optimistic in adulthood. A recent meta-analysis found that chronic physical illness in childhood is associated with greater psychological distress in adulthood^22^. However, to what extent childhood health conditions shape one’s level of optimism remains unclear.

The associations between individuals’ childhood experiences and adult levels of optimism may vary across national contexts. The Ecological Systems Theory posits that individuals’ development is shaped by cross-level interactions between influences from multiple interconnected systems, ranging from one’s immediate family environments to broader societal contexts^23^. In the case of optimism, prior studies found that one’s level of optimism is correlated with factors at multiple levels including personal characteristics, familial influences, neighborhood conditions, and cultural values and structural factors of the country where people live^24,25^. While the influences of cross-level interactions between these factors on optimism have seldom been studied empirically, based on the Ecological Systems Theory, country-level variations in how individual and familial experiences shape optimism are plausible. For instance, the stigma surrounding divorce and the level of societal support for children following parental divorce varies across countries, which may shape children’s psychosocial adjustment to parental divorce differently and may have long-term implications on their well-being^26^. As another example, the social welfare and child protection systems for supporting children from disadvantaged backgrounds (e.g., families of low socioeconomic status, victims of child abuse) and the policies and welfare states for migrant children vary widely across countries, which may have differential effects on shaping the influences of these childhood experiences on well-being later in life^27,28^. As a further example, countries that are considered more individualistic often emphasize autonomy and independence, whereas collectivist cultures tend to prioritize interpersonal relationships and group harmony, thus cultural values may shape the expectation for closeness and relatedness in parent-child relationships^29^. While positive parent-child relationships may be associated with better child well-being outcomes universally^29^, the strength of the association may vary across different cultures. Despite potential country variations in the influences of childhood experiences on well-being, to our knowledge country-level variations in childhood antecedents of optimism specifically remain understudied.

While past studies have identified some early life antecedents of optimism, several knowledge gaps remain. First, most prior studies used nonrepresentative samples, which limits generalizability of the study findings even within the countries in which these studies were conducted. Second, since most studies used data from the U.S. and European countries, there remains uncertainty in identifying likely childhood antecedents of optimism in other societies with varied cultures and norms. Cross-national differences are likely with regard to not only which early life factors matter but also the potency or extent to which any given factor contributes to shaping optimism over the life course. In addition to cultural values (e.g., collectivism/individualism, egalitarianism, uncertainty avoidance)^30^, societal structural factors will likely matter as well (e.g., GDP, income inequality, institutional trust)^31^. Greater understanding of the childhood antecedents of optimism across societies can inform the design of culturally relevant programs for increasing the likelihood that individuals can obtain and maintain optimism over the life course, which ultimately may contribute to enhanced population health and well-being.

To begin addressing these knowledge gaps, this study used data from 22 countries to examine the associations between a range of retrospectively reported childhood experiences and adult optimism levels, with nationally representative data within each country. Such knowledge would help provide initial evidence on potential targets for early interventions to enhance optimism later in life, including approaches that are universal across societies as well as strategies that need culturally specific adaptations. We hypothesized that these childhood personal attributes, and familial or social circumstances under consideration would have varied associations with adult optimism levels. We also anticipated that the strength of these associations would differ by country, reflecting diverse societal influences.

## METHODS

This study used data from the Global Flourishing Study (GFS). The description of the methods for the present study below has been adapted from VanderWeele et al.^32^ (the paper by VanderWeele et al. is scheduled to be published before the present study, and the reference will be updated when it becomes available). Further methodological detail regarding the Global Flourishing Study is available elsewhere^33–39^. The present study is a secondary analysis of the existing GFS data.

### Study Population

This study used wave 1 data from the Global Flourishing Study, a longitudinal study that enrolled 202,898 adults (age range: 18 to 99 years) from 22 culturally and geographically diverse countries, with nationally representative samples within each country. Survey items queried aspects of well-being including happiness, health status, sense of meaning and purpose, character, social relationships, and financial stability, along with a range of demographic, socioeconomic, political, religious, personality, childhood, community, and behavior factors. Cognitive interview and pilot-testing were conducted to validate the translation of survey questionnaires, and the translations were then sent to scholars in participating countries for evaluation^34^. Data collection was carried out by Gallup via a combination of modes (e.g., in-person, phone, web) that varied across countries^37^. Data for Wave 1 were collected principally during 2023, while some countries began data collection in 2022^33^. The following countries/territories were included in wave 1 data collection: Argentina, Australia, Brazil, Egypt, Germany, Hong Kong (Special Administrative Region of China), India, Indonesia, Israel, Japan, Kenya, Mexico, Nigeria, the Philippines, Poland, South Africa, Spain, Sweden, Tanzania, Turkey, United Kingdom, and the United States. The precise sampling design to ensure nationally representative samples varied by country^33^. The data are publicly available through the Center for Open Science (COS, https://www.cos.io/gfs). Details about the GFS study methodology and survey development are reported elsewhere^33,34^.

The present study used data from all GFS participants in wave 1 (N=202,898). Sampling weights were provided by Gallup (the weighting factors and weighting methods varied by country, depending on the specific sampling design in each country), and poststratification and nonresponse adjustments to ensure the sample was representative of the adult population in each country^33,37^. Further details regarding the GFS weighting procedure are provided elsewhere^37^.

Ethical approval was granted by the institutional review boards at Baylor University and Gallup, and all participants provided informed consent.

### Assessment of Optimism

Optimism was assessed with an item from the Revised Life Orientation Test^40^: “Overall, I expect more good things to happen to me than bad”. The response was rated on a Likert-type scale ranging from 0 (strongly disagree) to 10 (strongly agree). Specifically, participants chose an integer number from 0 to 10 to indicate their extent of agreement with the survey item statement on optimism, with a larger number indicating a higher level of optimism. The response was analyzed as a continuous score.

### Assessment of Childhood Factors

We investigated a range of individual-, interpersonal-, and familial-level childhood experiences, as described below. We also examined several childhood demographic factors (e.g., year of birth, gender, race/ethnicity), which are important demographic correlates of wellbeing. All childhood factors were retrospectively reported by the participants.

#### Relationship with parents

Participants reported their childhood relationships with parents in response to the question: “Please think about your relationship with your mother/father when you were growing up. In general, would you say that relationship was very good, somewhat good, somewhat bad, or very bad?”. Participants’ relationship with mother and father were queried separately. To reduce collinearity, in regression models the variables of relationship with mother and father were both dichotomized as “very/somewhat good” and “very /somewhat bad”. Responding “Does not apply” to either parental relationship indicator was treated as a dichotomous control variable for respondents who did not have a mother or father due to death or absence.

#### Parents’ marital status

Parents’ marital status was assessed with the question: “Were your parents married to each other when you were around 12 years old?”. Response options included “married”, “divorced”, “never married”, and “one or both of them had died”.

#### Subjective financial status

Participants reported the financial status of their family as it was when they were growing up, in response to the question: “which of these phrases comes closest to your own feelings about your family’s household income when you were growing up, such as when you were around 12 years old?”. Response options included “lived comfortably”, “got by”, “found it difficult”, and “found it very difficult”.

#### Childhood abuse

Childhood abuse victimization was assessed with the question: “Were you ever physically or sexually abused when you were growing up?”. Responses included “yes” and “no”. This information was not obtained in Israel due to restrictions on asking such questions.

#### Felt like an outsider in family

Participants’ sense of family belonging during childhood was assessed with the question: “When you were growing up, did you feel like an outsider in your family”. Response options included “yes” and “no”.

#### Self-rated health

Participants reported their health status during childhood in response to the question: “In general, how was your health when you were growing up? Was it excellent, very good, good, fair, or poor?”

#### Immigration status

Immigration status was assessed with the question “Were you born in this country?” The responses include “yes” and “no”.

#### Religious service attendance

Participants’ frequency of religious service attendance during childhood was assessed with a question: “How often did you attend religious services or worship at a temple, mosque, shrine, church or other religious building when you were around 12 years old”. The response options range from 1 (at least once a week) to 4 (never).

#### Birth Year

Participants reported their current age (in years), and their birth years were calculated as the year of data collection minus their current age.

#### Gender

Participants reported their gender, and the response options include “male”, “female”, and “other gender identities”.

#### Religious affiliation

Religious affiliation in childhood was measured with the question: “What was your religion when you were 12 years old”. Response options include 15 major religions (e.g., Christianity, Islam, Hinduism, Buddhism, Judaism etc.), “some other religion”, and “No religion/Atheist/Agnostic”. The exact response categories varied as appropriate for each country, and this variable was included in the country-specific analyses only but not in the meta-analysis across countries^41^. To reduce data sparsity, in regression models the response categories of religious affiliation with a prevalence <3% were collapsed. “No religion/Atheist/Agnostic” was used as the reference group when at least 3% of the observed sample within the country endorsed this category; otherwise, the most prominent religious group was used as the reference category.

#### Race and ethnicity

Race and ethnicity were assessed in most but not all countries (Germany, Japan, Spain, and Sweden had restriction on collecting such data). Response options varied as appropriate for each country, so this variable was included in the country-specific analyses only but not in the meta-analysis across countries. To reduce data sparsity, in regression models race and ethnicity was dichotomized as racial and ethnic plurality (the category with the largest proportion) and minority (collapsing other categories) in each country.

### Statistical Analyses

Descriptive analyses consider the distribution of childhood factors in individual countries and in the total combined sample. The results were weighted to be nationally representative within each country.

In the country-specific regression analyses, a weighted linear regression model with complex survey adjusted standard errors was used to regress optimism on *all childhood factors simultaneously* to evaluate their associations with optimism independently from each other. A Wald-type test was conducted to obtain a global (joint) test of the effect of all categories within a childhood factor resulting in a single global p-value for each childhood factor.

Random effects meta-analysis^42,43^ was conducted to pool regression coefficients from the country-specific analyses, along with confidence intervals, lower and upper limits 95% prediction intervals, heterogeneity (τ), and I^2^ for estimating variation within a given childhood factor category across countries^44^. Forest plots of estimates are presented in the online supplement. Because the variable of childhood abuse was not assessed in Israel, the meta-analysis pooled the country-specific estimates for childhood abuse across the other 21 countries. Religious affiliation and race and ethnicity (when available) were used as control variables within country (their coefficients are reported in the online supplement), but these coefficients themselves were not included in the meta-analyses because their response options varied by country. A pooled global p-value^45^ was reported across countries to evaluate if the association between each childhood factor and adult optimism holds within at least one country. Bonferroni corrected p-value thresholds are provided based on the number of childhood factors^46,47^.

Meta-analyses were conducted in R (R Core Team, 2024) using the metafor package^48^, and country-specific analyses were conducted using SAS 9.4 (SAS Institute, Inc). All statistical tests were 2-sided. For each childhood predictor, in both the meta-analyses and country-specific analyses we calculated E-values to evaluate sensitivity of the observed associations to potential unmeasured confounding. An E-value is the minimum strength of the association an unmeasured confounder must have with both the outcome and the predictor, above and beyond all measured covariates, for an unmeasured confounder to explain away the observed predictor-outcome association^49^. As a supplementary analysis, a population weighted meta-analysis was conducted to pool the regression coefficients from country-specific analyses. All analyses were pre-registered with COS prior to data access, with only slight subsequent modification in covariate adjustments due to multicollinearity (https://doi.org/10.17605/OSF.IO/FB254); all code to reproduce analyses are openly available in an online repository^36^.

### Missing Data

In the total combined sample, 0.41% of the participants had missing data on optimism, and missing data on childhood factors ranged from 0.01% to 5% (except race/ethnicity, which was not measured in some countries). We conducted multivariate imputation by chained equations (with 5 imputed datasets created) to impute missing data on all variables in each country separately^50–52^. We included sampling weights in the imputation model to account for missingness related to probability of inclusion.

### Accounting for Complex Sampling Design

The GFS used different sampling scheme across countries based on availability of existing panels and recruitment needs^33^. All analyses accounted for the complex survey design components by including weights, primary sampling units, and strata. Additional methodological details are reported elsewhere^36^.

## RESULTS

### Descriptive Analyses

In the combined cross-national sample, the weighted distribution of the childhood factors were as follows (Table 1): very/somewhat good relationships with mother (89%) and father (80%), parents were married (75%), high subjective financial status (lived comfortably/got by, 76%), no experience of child abuse (82%), no feeling like an outsider in the family (84%), excellent/very good childhood health (64%), being native-born (94%), and attending religious services at least once/week in childhood (41%). Respondents’ age ranged from 18 to 99 years and the weighted proportion of people from different birth cohorts was relatively similar (except for fewer participants from earlier birth cohorts with current age above 70 years). The weighted sample had a balanced ratio of female (51%) and male (48%) participants, and a small proportion (0.3%) who identified as other gender.

**Table 1.**
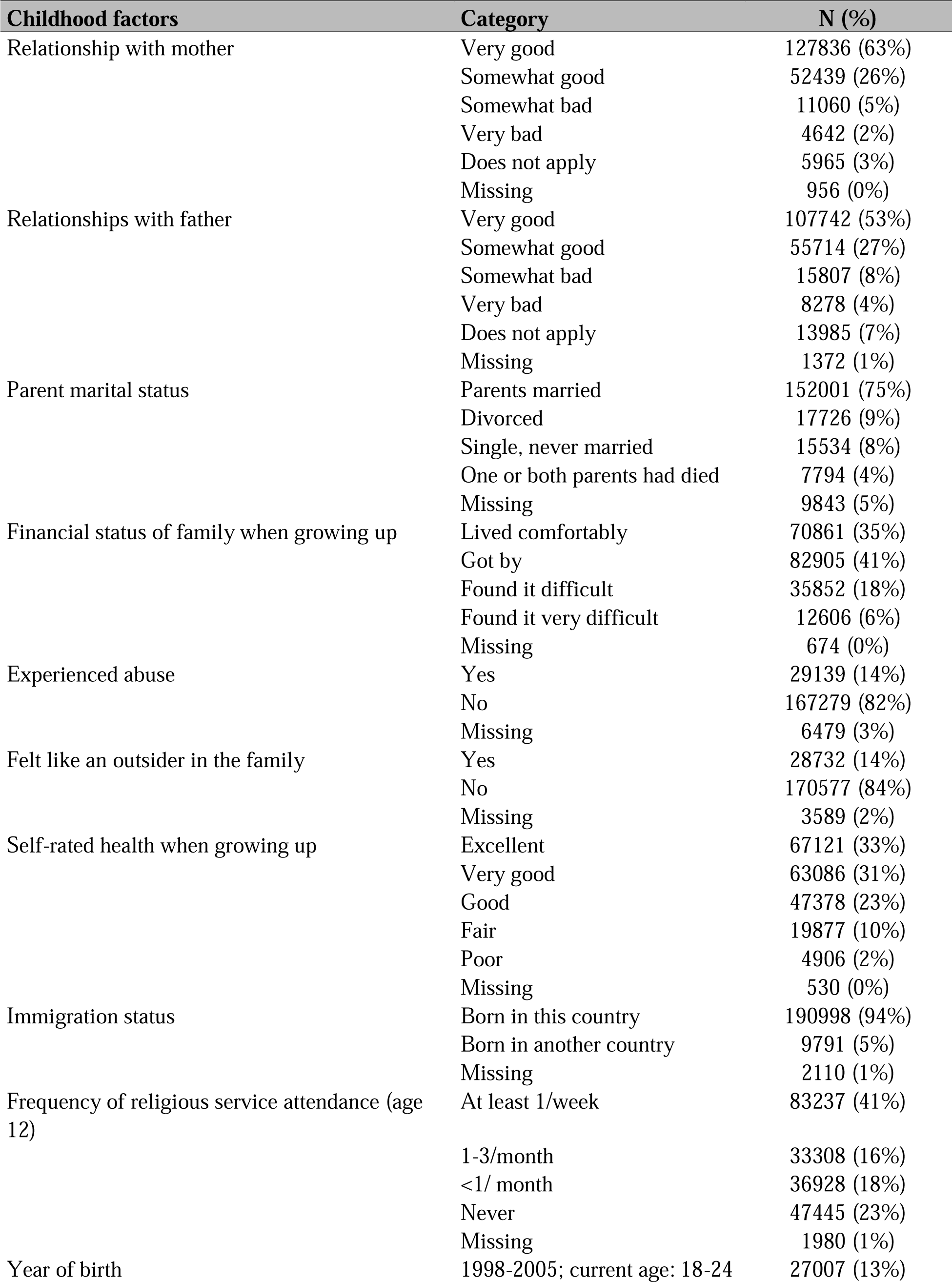
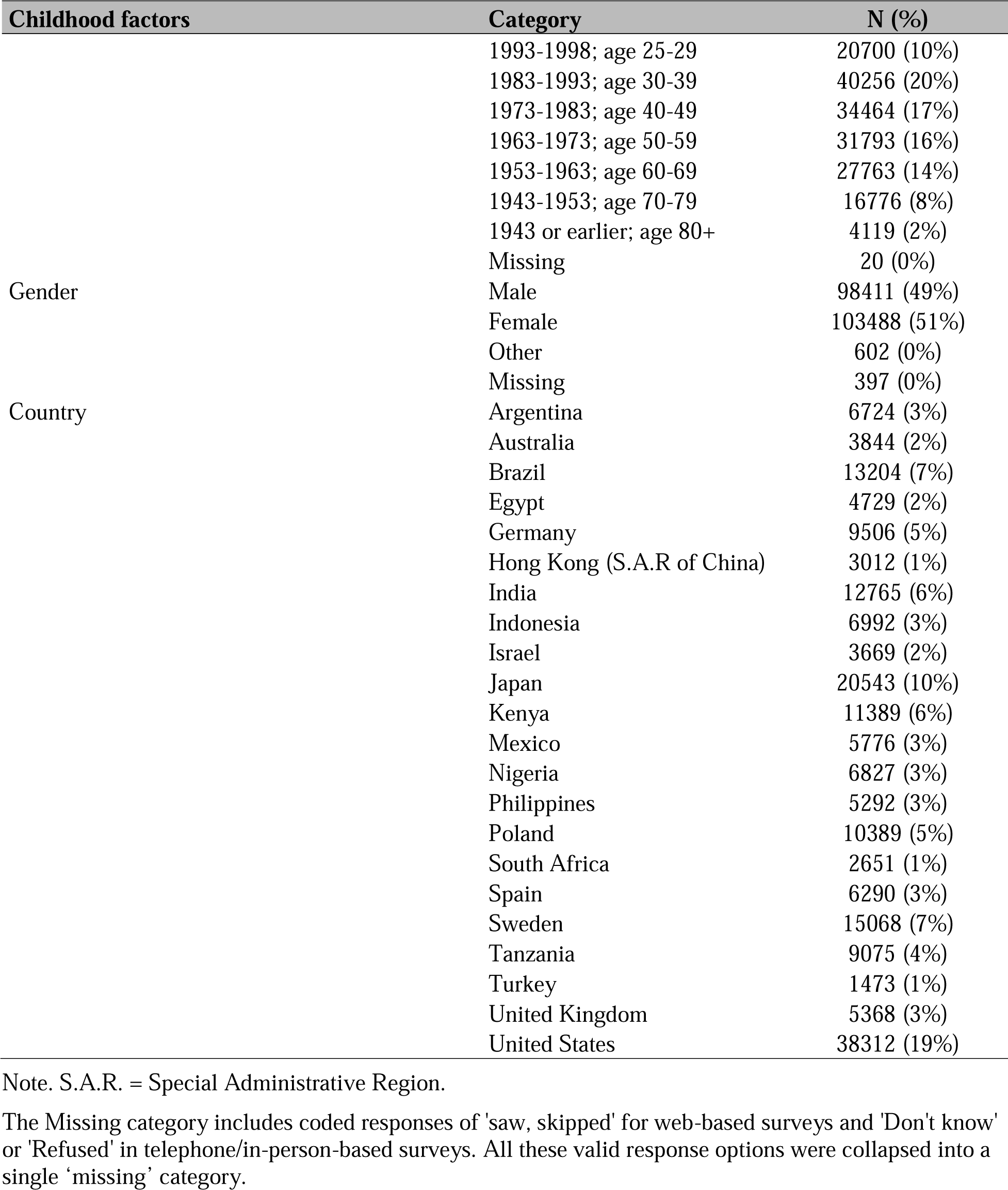
Distribution of Childhood Factors in the Overall Sample Combined Across 22 Countries (Weighted to Be Nationally Representative within Each Country, N=202,898).

The country-specific mean optimism score and standard deviation are reported in Supplementary Table S1. The weighted distribution of participant childhood characteristics in each individual country are reported in Supplementary Table S2A to S23A.

### Childhood Factors and Adult Optimism Levels: Pooled Estimates Across Countries

The random effect meta-analyses pooling regression coefficients from country-specific analyses suggest that most childhood factors under consideration are strongly associated with optimism levels in adulthood (see Table 2). All global p-values are below the Bonferroni corrected significance threshold (*p*<0.004), providing evidence that in at least one country each of these childhood factors are likely associated with adult optimism levels. For instance, on average across countries, the adult optimism score was on average 0.17 points higher (95% confidence interval [CI]: 0.07, 0.27) among those who had very/somewhat good relationships with mother in childhood, as compared to those who had very/somewhat bad relationship with mother, holding the values of all other covariates constant. Likewise, having very/somewhat good (vs. very/somewhat bad) relationship with one’s father was associated with higher levels of adult optimism (β=0.07, 95% CI=0.02, 0.13). Having divorced vs married parents was associated with lower optimism (β=-0.11, 95% CI: −0.21, −0.00). Experiencing childhood abuse was also associated with lower adult optimism levels (β=-0.24, 95% CI: −0.32, −0.15), as were feelings like an outsider in the family (β=-0.26, 95% CI: −0.35, −0.18). Furthermore, better childhood subjective financial status (e.g., β _lived comfortability vs. got by_=0.10, 95% CI: 0.03, 0.17), better self-rated health (e.g., β _excellent vs. good_=0.43, 95% CI: 0.25 0.61), and more frequent religious service attendance (e.g., β _at least once/week vs. never_=0.26, 95% CI: 0.17, 0.35) were each positively associated with higher optimism levels in a monotonic fashion. There was also evidence suggesting that earlier (vs. the most recent) birth cohorts reported higher adult optimism levels (e.g., β _birth cohort 1953-1963 vs. 1998-2005_=0.20, 95% CI: 0.02, 0.38). Females also reported higher optimism than males on average (β=0.18, 95% CI=0.11, 0.25). The confidence interval for the association of optimism with immigration status (native versus not native-born) included the null value but the global p-value was <.001, indicating immigration status was associated with optimism in at least one country but not on average across countries. In terms of the extent to which associations of childhood factors with adult optimism levels varied across countries, as assessed by T, the highest heterogeneity was evident for the childhood factors of parent marital status, childhood self-rated health, immigration status, birth cohort membership (especially the earlier birth cohorts), and gender (other gender identities vs. male in particular).

**Table 2.**
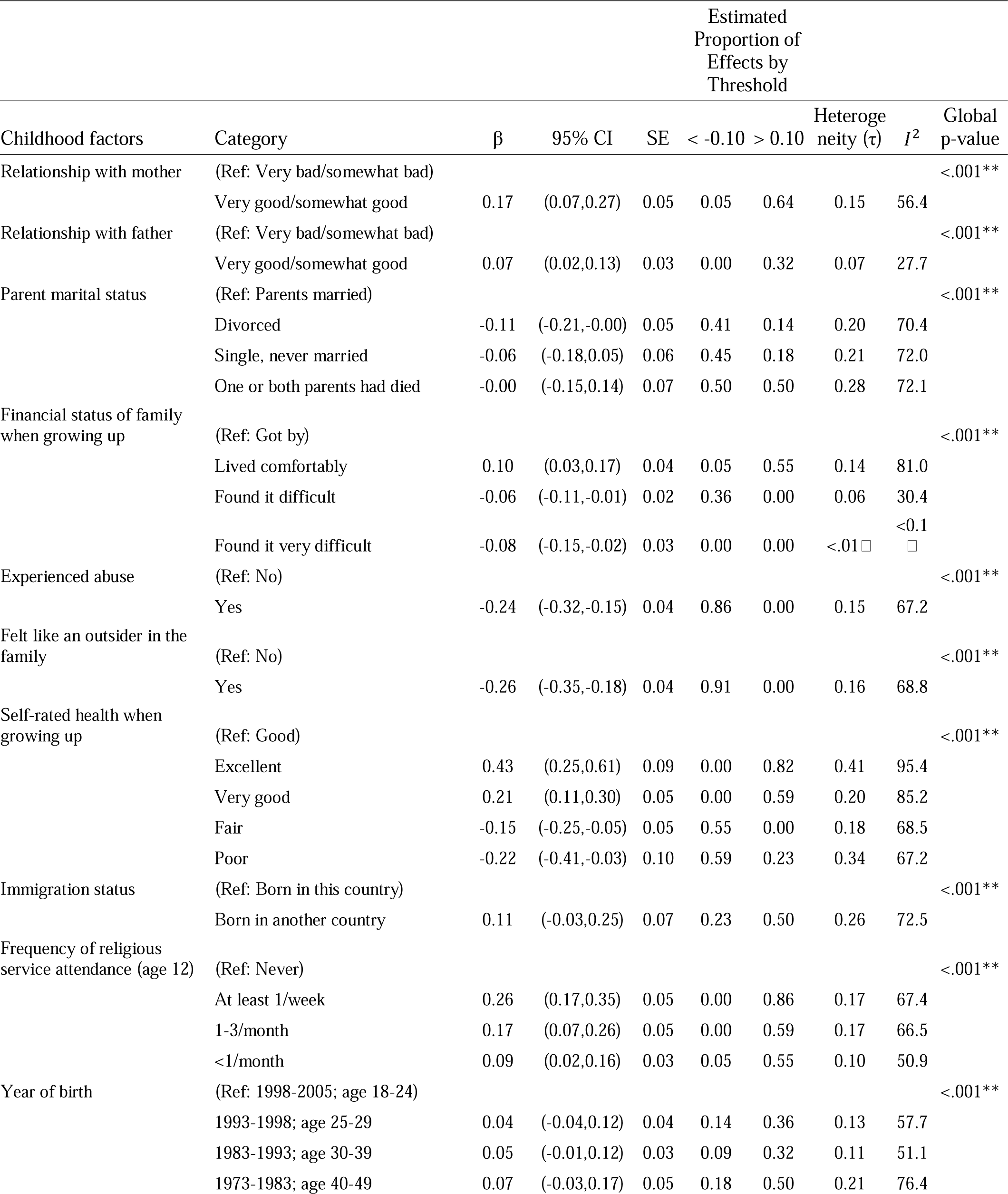
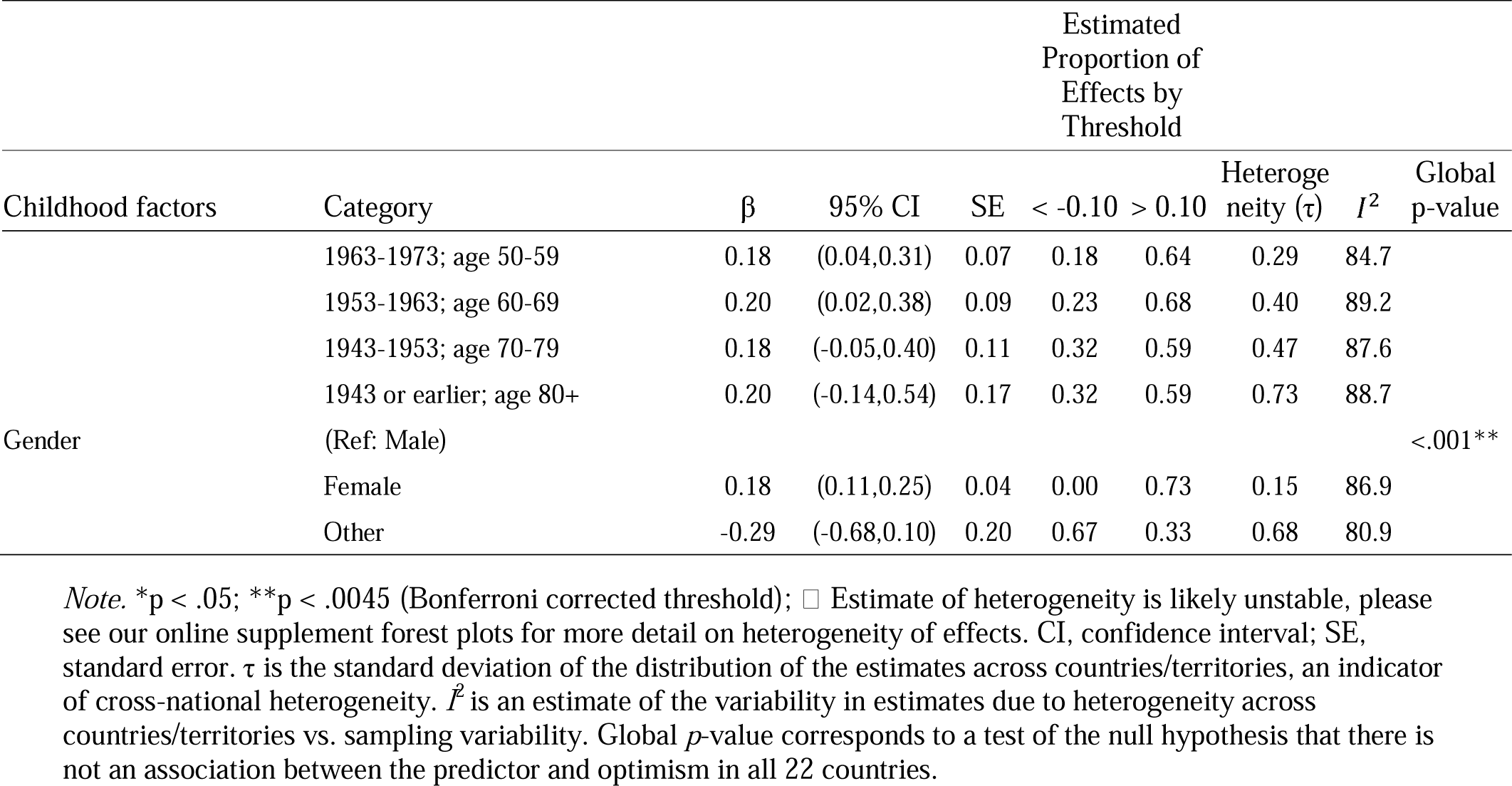
Random Effects Meta-Analysis of Association between Childhood Factors and Levels of Optimism in Adulthood.

The calculated “E values” provide evidence on the robustness of the observed associations between childhood factors and adult optimism in the meta-analysis to potential unmeasured confounding (Table 3). For instance, to explain away the positive association between having a better relationship with one’s mother and having higher adult optimism levels (β=0.17), an unmeasured confounder associated with both increased likelihood of having a positive relationship with mother and higher optimism by risk ratios of 1.36 each, above and beyond all measured covariates, could suffice but weaker joint confounder associations could not. Likewise, to shift the confidence interval to include the null value, an unmeasured confounder associated with both having a positive relationship with mother and higher optimism by risk ratios of 1.21-fold each could suffice, but weaker joint confounder association could not. The E-values for several other childhood factors (e.g., childhood abuse, self-rated health, religious service attendance) were of similar or larger magnitude than for relationship with mother. However, the association for some other childhood categories (e.g., some categories of parental marital status and year of birth) are likely to be explained away by unmeasured confounding.

**Table 3.** Sensitivity of Meta-Analyzed Associations between Childhood Factors and Optimism in Adulthood to Potential Unmeasured Confounding.

| Childhood factors | Category | E-value for $\beta$ | E-value for 95% Confidence interval |
| --- | --- | --- | --- |
| Relationship with mother | (Ref: Very bad/somewhat bad) |  |  |
|  | Very good/somewhat good | 1.36 | 1.21 |
| Relationship with father | (Ref: Very bad/somewhat bad) |  |  |
|  | Very good/somewhat good | 1.21 | 1.09 |
| Parent marital status | (Ref: Parents married) |  |  |
|  | Divorced | 1.27 | 1.03 |
|  | Single, never married | 1.20 | 1.00 |
|  | One or both parents had died | 1.03 | 1.00 |
| Financial status of family when growing up | (Ref: Got by) |  |  |
|  | Lived comfortably | 1.26 | 1.14 |
|  | Found it difficult | 1.19 | 1.08 |
|  | Found it very difficult | 1.23 | 1.10 |
| Experienced Abuse | (Ref: No) |  |  |
|  | Yes | 1.45 | 1.34 |
| Felt like an outsider in the family | (Ref: No) |  |  |
|  | Yes | 1.49 | 1.38 |
| Self-rated health when growing up | (Ref: Good) |  |  |
|  | Excellent | 1.70 | 1.47 |
|  | Very good | 1.41 | 1.28 |
|  | Fair | 1.33 | 1.18 |
|  | Poor | 1.43 | 1.14 |
| Immigration status | (Ref: Born in this country) |  |  |
|  | Born in another country | 1.27 | 1.00 |
| Frequency of religious service attendance (age 12) | (Ref: Never) |  |  |
|  | At least 1/week | 1.48 | 1.36 |
|  | 1-3/month | 1.36 | 1.21 |
|  | <1/month | 1.24 | 1.11 |
| Year of birth | (Ref: 1998-2005; age 18-24) |  |  |
|  | 1993-1998; age 25-29 | 1.14 | 1.00 |
|  | 1983-1993; age 30-39 | 1.17 | 1.00 |
|  | 1973-1983; age 40-49 | 1.21 | 1.00 |
|  | 1963-1973; age 50-59 | 1.37 | 1.16 |
|  | 1953-1963; age 60-69 | 1.40 | 1.10 |
|  | 1943-1953; age 70-79 | 1.37 | 1.00 |
|  | 1943 or earlier; age 80+ | 1.40 | 1.00 |
| Gender | (Ref: Male) |  |  |
|  | Female | 1.37 | 1.27 |
|  | Other | 1.52 | 1.00 |

Notes: The *E*-value for the effect estimate is the minimum strength of association (on the risk ratio scale) that an unmeasured confounder would need to have with both the predictor and the outcome to entirely explain away the observed association between them, conditional on the measured covariates. The *E*-value for the limit of the 95% confidence interval closest to the null denotes the minimum strength of association (on the risk ratio scale) that an unmeasured confounder would need to have with both the predictor and the outcome to shift the confidence interval to include the null value, conditional on the measured covariates.

### Childhood Factors and Adult Optimism Levels: Country-Specific Estimates

In country-specific analyses (Supplementary Tables S2B to S23B; Supplementary Figures S1 to S27), childhood self-rated health, year of birth, gender, and religious affiliation were each associated with adult optimism levels in more than half of the countries evaluated (global p-values all <.05). However, the magnitude of the associations varied widely across countries for each of these childhood factors. For instance, while better childhood self-rated health (excellent vs. good) was associated with substantially higher levels of optimism in adulthood across a range of culturally diverse countries (e.g., Hong Kong, Japan, Sweden, U.S., Brazil, Poland), the association was particularly strong in high-income countries.

Several other childhood factors also had a p value <.05 in many but fewer than half of the countries. These include: parent marital status, subjective financial status, childhood abuse, felt like an outsider in the family, religious service attendance, and race and ethnicity. Specifically, in most countries, reporting having a parent divorced or being single/never married (vs. married) was associated with lower optimism in adulthood; however, there were exceptions whereby having parents who were single/never married (e.g., in Argentina, Australia, Egypt) or divorced (e.g., in Japan, Sweden) was related to higher adult optimism levels. Interestingly, subjective financial status in childhood was not associated with adult optimism in most low- or middle-income countries (e.g., Argentina, India, Kenya, Poland, Turkey), whereas there was a positive association in several high-income countries/territories (e.g., Germany, Hong Kong, Japan, Sweden, U.S.). When associations were evident, experiencing childhood abuse and feeling like an outsider in the family were each associated with lower adult optimism; however, in countries where the associations included the null value, it is worth noting that the prevalences of abuse and feeling like an outsider were mostly low (mostly ≤ 10%, e.g., in Egypt, Indonesia, Poland, Spain, Tanzania), which may affect statistical power. Interestingly, regular religious service attendance in childhood was associated with higher adult levels of optimism in some of the most secular countries^53^ (e.g., Japan, Hong Kong, Germany, Spain, U.K.).

Contrary to findings in the meta-analysis, in country-specific analyses the associations between having a positive relationship with either mother or father and adult optimism levels often included the null value. This might be due to the small number of participants who reported “very/somewhat bad relationships” in individual countries (in over half of the countries <5% and <10% of the participants reported having very/somewhat bad relationship with mother and with father). Furthermore, the associations of immigration status with optimism were evident in only a handful of countries; among these, being born in another country was generally associated with being more optimistic (except in Brazil).

Results of the sensitivity analyses for unmeasured confounding (i.e., the E-values) for the country-specific analyses are presented in Supplementary Tables S2C to S23C. The population weighted meta-analysis that pooled results across countries considering population sizes in each country yielded similar results as the random-effects meta-analysis, except that the associations for relationships with mother and father became weaker (Supplementary Tables S24 and S25).

This may be due to the heavy weight in the population weighted meta-analysis given to India, where very few participants reported having very/somewhat bad relationships with mother and father.

## DISCUSSION

Having an optimistic mindset may be a desirable health and well-being asset^54^. However, childhood origins of optimism remain understudied^55^. This study extends the literature by examining a range of individual-, interpersonal-, and familial-level factors during childhood as candidate antecedents of adult optimism across 22 culturally diverse countries, with nationally representative samples from within each country.

Congruent with prior research^13,56–58^, this study suggests that familial relationships in early life may have long-term associations with individuals’ optimism; this study extends such evidence to a wide range of diverse countries. Specifically, in the pooled results across countries felt like an outsider in the family is one of the childhood factors that is most strongly associated with lower levels of optimism in adulthood; conversely, having a positive relationship with mother and father are each related to higher adult optimism levels. Such findings may be understood through the lens of the Attachment Theory^59^: love and responsiveness from caregivers provide a base of secure attachment, contributing to the child’s development of trust, self-worth, and possibly other psychological attributes such as optimism and resilience^13^.

Considering the country-specific estimates, felt like an outsider in the family was consistently associated with substantially lower levels of adult optimism across many culturally diverse countries (e.g., Australia, U.S., Germany, Brazil, Japan, India, Turkey), suggesting that enhancing a sense of family belonging in early life may be a universal approach to fostering optimism across societies.

Consistent with prior literature^16,17,60^, this study finds that adverse childhood experiences (e.g., childhood abuse, experiencing parental divorce, financial hardship) are associated with lower optimism levels in adulthood on average across countries. Childhood traumatic experiences, such as childhood abuse and parental divorce, may induce generalized negative cognitive schemas and alter neurobiological development, leading to a reduced sense of mastery and lower capacities to envision positive future outcomes^15^. The findings of childhood financial hardship and lower adult optimism levels may be contextualized through the lens of the Reserve Capacity Framework^61^, which posits that individuals who grow up with greater socioeconomic disadvantage are more likely to encounter major life stressors and have fewer opportunities to develop psychosocial resources (e.g., adaptive coping strategies, sense of purpose, optimism) that help buffer stress. It is noteworthy, however, that some of these associations with specific adverse childhood experiences varied by country. For instance, individuals with divorced parents reported lower adult optimism pooled across the 22 countries and, for instance, in India (a country with a very low national divorce rate), whereas higher adult optimism in Sweden (a country with a high divorce rate and a prominent singlehood culture)^62^, suggesting that social norms around marriage may shape individuals’ experiences of parental divorce and its influence on optimism later in life.

This study also suggests that greater resources in material dimensions during childhood (e.g., financial resources, good health status) are strongly associated with higher levels of adult optimism. In the pooled estimates across countries, living financially comfortably in childhood and having excellent/good health in childhood were each associated with substantially higher adult optimism levels. Individuals with more material resources are less likely to encounter stressors and have more opportunities to develop reserve capacity (e.g., optimism) for coping with stress^61^; meanwhile, material resources in childhood may also contribute to fostering optimism indirectly through promoting success in important facets of life (e.g., educational achievement, social network, job security)^61^. It is worth noting that the positive associations of high childhood financial status and optimal childhood health with adult optimism levels are particularly strong in some of the wealthiest countries/territories (e.g., Japan, Sweden, U.S., Germany, Hong Kong). We speculate that in these high-income countries, the overall wealth and stability may amplify the positive impact of a financially comfortable upbringing and optimal childhood health on optimism, whereby children are more likely to deploy material resources (e.g., money, knowledge, prestige, power, social connections) that help them navigate challenges effectively. Financial resources might thereby contribute to not only “resourced” forms of optimism, but also greater agentive optimism as well^63^. Conversely, the countries with the weakest associations between a financially comfortable upbringing, optimal childhood health, and adult optimism (e.g., India, Nigeria, Israel, Tanzania) are mostly countries that have lower economic development levels and/or have faced political or economic conflicts in the recent past. In these societies, the lack of stability and ongoing conflicts might disrupt individuals’ access to and/or the ability to effectively deploy material resources, thereby diminishing the potential benefits of a financially comfortable upbringing and optimal childhood health.

Optimism holds a deep-rooted tradition in many world religions, especially in the so-called “salvation religions” (e.g., Christianity, Islam, Jainism, Buddhism, etc.) that emphasize liberation through means such as faith and good deeds^64^. Prior empirical evidence has suggested that having a religious upbringing is associated with greater likelihood of exhibiting elements of positive psychological functioning in adulthood (e.g., greater purpose in life)^21^, although its association with optimism has seldom been examined directly. This study provides novel evidence on a strong dose-response association between religious service attendance during childhood and higher levels of adult optimism on average across countries. Interestingly, the country-specific results further indicate that the associations hold in some of the most secular countries^53^ (e.g., Japan, Sweden, Hong Kong, U.K.). We speculate that individuals who participate in religious congregations in secular regions are likely those with a particularly strong sense of faith, meaning, and hope.

This study has some limitations that need to be considered. First, optimism was assessed with a single item from the LOT-R. Although the LOT-R has been validated in many countries, the GFS cognitive interview found that some respondents had difficulty in understanding the optimism question/translation^34^, suggesting the findings may be affected by measurement error.

Second, our adult respondents reported on all childhood experiences retrospectively at the same time as when they reported their optimism levels, which raises concerns about recall bias. For recall bias to completely explain away the observed associations will require that the effect of adult optimism on biasing retrospective assessments of the childhood factors would essentially have to be at least as strong as the observed associations themselves^65^, and many of these are quite substantial. It is, however, important to consider potential recall bias when interpreting the findings, and it is also worth noting that causality cannot be inferred from this observation study. Third, there is potential collinearity between some childhood factors (e.g., relationship with mother, relationship with father, felt like an outsider in the family, childhood abuse) when they were simultaneously included in the regression models. We took several approaches to reducing concerns about collinearity (e.g., collapsing response categories, removing some variables from the analyses; see details in our pre-registration), but the possibility of collinearity cannot be completely ruled out. Fourth, while we conducted sensitivity analysis to evaluate robustness of the results to unmeasured confounding, the possibility of unmeasured confounding cannot be ruled out and it is a limitation that needs to be considered in all observational studies. Fifth, we considered the optimism score as a continuous variable and used linear regression model in our analyses. Future studies may consider replicating the findings with alternative modeling strategies such as the ordinal regression model. Finally, there are other important potential childhood factors that may shape adult optimism levels that are not examined in this study, such as school experiences, peer relationships, neighborhood conditions, community and cultural support during childhood^25,66^, which warrant further investigation in future studies.

Several low-cost approaches to enhancing optimism in adulthood have been developed such as the Best Possible Self intervention and mindfulness-based programs^67,68^. A prior meta-analysis has suggested short-term effectiveness of such programs in increasing optimism^4^, although other work suggests such interventions may have small effects and more substantial interventions are needed. As a result, identifying and targeting social and cultural conditions that increase the prevalence and levels of optimism in the population may prove a more effective approach to enhancing this facet of positive psychological function and facilitating a more equitable distribution of optimism in diverse populations. Findings from the current study are in line with the growing body of literature suggesting that an optimistic mindset may be cultivated starting from an early age, and point to a variety of intrapersonal, interpersonal, familial, societal, and cultural factors during childhood that may matter. To our knowledge, this is the first study that examines a wide range of childhood correlates of adult optimism simultaneously across multiple countries. The findings in this study need to be replicated in future studies with longitudinal data. Further investigations on whether, when, and how childhood factors can meaningfully shape adult levels of optimism, accounting for varied social and cultural conditions in individual countries, will advance our understanding of culturally appropriate approaches to fostering an optimistic mindset, a promising psychological asset for enhancing population health and well-being.

## Acknowledgements

The Global Flourishing Study was supported by funding from the John Templeton Foundation (grant #61665), Templeton Religion Trust (#1308), Templeton World Charity Foundation (#0605), Well-Being for Planet Earth Foundation, Fetzer Institute (#4354), Well Being Trust, Paul L. Foster Family Foundation, and the David and Carol Myers Foundation. The funding source had no impact on the study design; on the collection, analysis and interpretation of data; on the writing of the report; or on the decision to submit the article for publication.

## Author Contributions

B.R. J., and T.J.V. developed the study concept. Y.C., L.D.K., E.S.K., H.K., K.S., R.N.P., R.W., B.R. J., and T.J.V. contributed to the study design. Y.C. and R.N.P. had full access to the data, conducted data analyses, and take responsibility for the integrity of the data and accuracy of the data analysis. Y.C. drafted the manuscript. L.D.K., E.S.K., H.K., K.S., R.N.P., R.W., B.R. J., and T.J.V. provided critical revisions, and approved the final submitted version of the manuscript.

## Ethics approval and consent to participate

Ethical approval was granted by the institutional review boards at Baylor University (IRB Reference #: 6431841317) and Gallup (IRB Reference #: 2021-11-02), and all participants provided informed consent.

## Data Availability

Data for Wave 1 of the Global Flourishing Study is available through the Center for Open Science upon submission of a pre-registration (https://doi.org/10.17605/OSF.IO/3JTZ8), and will be openly available without pre-registration beginning February 2025. Please see https://www.cos.io/gfs-access-data for more information about data access.

## Code Availability

All analyses were pre-registered with COS prior to data access (https://doi.org/10.17605/OSF.IO/FB254). All code to reproduce analyses are openly available in the online OSF repository (https://doi.org/10.17605/osf.io/vbype).

## Completing Interests

Tyler VanderWeele reports consulting fees from Gloo Inc., along with shared revenue received by Harvard University in its license agreement with Gloo according to the University IP policy. Other authors have no conflicts of interest to declare.

