## Supplementary file for "When the Glass is Half Full: Early Life Experiences and Adult Optimism in 22 Countries"

- Table S1: Mean optimism score and standard deviation in each country.
- Table S2A – S23C: Supplementary tables for country-specific analyses.
- Table S24: Supplementary table for population weighted meta-analysis.
- Table S25: Supplementary table for population weighted meta-analysis of E-values.
- Figure S1– S27: Supplementary figures for estimates of childhood predictor categories.

***Supplementary Table S1. Means of Optimism with Standard Deviations by Country.***

| **Country** | **Mean** | **Standard Deviation** |
| --- | --- | --- |
| Argentina | 8.86 | 1.83 |
| Australia | 7.41 | 2.27 |
| Brazil | 9.22 | 1.62 |
| Egypt | 7.67 | 2.57 |
| Germany | 7.50 | 2.11 |
| Hong Kong (S.A.R of China) | 7.23 | 2.10 |
| India | 8.14 | 2.90 |
| Indonesia | 9.15 | 1.69 |
| Israel | 8.38 | 1.78 |
| Japan | 6.74 | 2.27 |
| Kenya | 8.92 | 2.20 |
| Mexico | 9.06 | 1.66 |
| Nigeria | 8.90 | 1.77 |
| Philippines | 8.79 | 2.01 |
| Poland | 7.72 | 1.83 |
| South Africa | 8.22 | 2.22 |
| Spain | 8.25 | 1.97 |
| Sweden | 7.30 | 2.32 |
| Tanzania | 8.75 | 2.36 |
| Türkiye | 7.66 | 2.49 |
| United Kingdom | 6.99 | 2.44 |
| United States | 7.65 | 2.29 |

Note. S.A.R. = Special Administrative Region.

***Supplementary Table S2A. Nationally Representative Childhood Descriptive Statistics for Argentina***

| **Variable** | **Category** | **N (%)** |
| --- | --- | --- |
| Relationship with mother | Very good | 4463 (66%) |
|  | Somewhat good | 1436 (21%) |
|  | Somewhat bad | 299 (4%) |
|  | Very bad | 216 (3%) |
|  | Does not apply | 273 (4%) |
|  | Missing | 36 (1%) |
| Relationships with father | Very good | 3612 (54%) |
|  | Somewhat good | 1537 (23%) |
|  | Somewhat bad | 440 (7%) |
|  | Very bad | 401 (6%) |
|  | Does not apply | 694 (10%) |
|  | Missing | 39 (1%) |
| Parent marital status | Parents married | 4110 (61%) |
|  | Divorced | 637 (9%) |
|  | Single, never married | 1368 (20%) |
|  | One or both parents had died | 199 (3%) |
|  | Missing | 410 (6%) |
| Subjective financial status of family growing up | Lived comfortably | 2042 (30%) |
|  | Got by | 2305 (34%) |
|  | Found it difficult | 1789 (27%) |
|  | Found it very difficult | 569 (8%) |
|  | Missing | 19 (0%) |
| Abuse | Yes | 1302 (19%) |
|  | No | 5271 (78%) |
|  | Missing | 151 (2%) |
| Outsider growing up | Yes | 1165 (17%) |
|  | No | 5458 (81%) |
|  | Missing | 101 (2%) |
| Self-rated health growing up | Excellent | 2402 (36%) |
|  | Very good | 1819 (27%) |
|  | Good | 1830 (27%) |
|  | Fair | 505 (8%) |
|  | Poor | 156 (2%) |
|  | Missing | 12 (0%) |
| Immigration status | Born in this country | 6346 (94%) |
|  | Born in another country | 348 (5%) |
|  | Missing | 29 (0%) |
| Age 12 religious service attendance | At least 1/week | 2601 (39%) |
|  | 1-3/month | 1204 (18%) |
|  | <1/ month | 1059 (16%) |
|  | Never | 1808 (27%) |
|  | Missing | 53 (1%) |
| Year of birth | 1998-2005; current age: 18-24 | 1108 (16%) |
|  | 1993-1998; age 25-29 | 719 (11%) |
|  | 1983-1993; age 30-39 | 1432 (21%) |
|  | 1973-1983; age 40-49 | 1254 (19%) |
|  | 1963-1973; age 50-59 | 1014 (15%) |
|  | 1953-1963; age 60-69 | 730 (11%) |
|  | 1943-1953; age 70-79 | 356 (5%) |
|  | 1943 or earlier; age 80+ | 112 (2%) |
| Gender | Male | 3143 (47%) |
|  | Female | 3542 (53%) |
|  | Other | 21 (0%) |
|  | Missing | 18 (0%) |
| Religious affiliation at age 12 | Christianity | 5805 (86%) |
|  | Islam | 11 (0%) |
|  | Hinduism | 2 (0%) |
|  | Buddhism | 3 (0%) |
|  | Judaism | 51 (1%) |
|  | Sikhism | 5 (0%) |
|  | Taoism | 1 (0%) |
|  | Primal, Animist, or Folk religion | 17 (0%) |
|  | Some other religion | 10 (0%) |
|  | No religion/Atheist/Agnostic | 697 (10%) |
|  | Missing | 122 (2%) |
| Race/ethnicity | Asian | 43 (1%) |
|  | Black | 95 (1%) |
|  | Indigenous | 129 (2%) |
|  | Mestizo(a) | 1801 (27%) |
|  | Mullato(a) | 75 (1%) |
|  | White | 3406 (51%) |
|  | Other | 104 (2%) |
|  | Missing | 1070 (16%) |

***Supplementary Table S2B. Regression of Optimism on Childhood Predictors for Argentina***

| **Variable** | **Category** | **Estimate** | **SE** | **95% CI** | **Global P-value** |
| --- | --- | --- | --- | --- | --- |
| Relationship with mother (ref: Very bad/Somewhat bad) | Very good/Somewhat good | 0.32 | 0.13 | (0.06,0.58) | 0.015 |
| Relationship with father (ref: Very bad/Somewhat bad) | Very good/Somewhat good | 0.02 | 0.11 | (-0.19,0.23) | 0.830 |
| Parent marital status (ref: Married) | Divorced | 0.03 | 0.12 | (-0.20,0.27) | 0.067 |
|  | Single, never married | 0.19 | 0.09 | (0.01,0.37) |  |
|  | One or both parents had died | 0.29 | 0.16 | (-0.03,0.60) |  |
| Subjective financial status of family growing up (ref: Got by) | Lived comfortably | 0.04 | 0.07 | (-0.10,0.19) | 0.528 |
|  | Found it difficult | -0.03 | 0.08 | (-0.19,0.12) |  |
|  | Found it very difficult | 0.15 | 0.13 | (-0.10,0.39) |  |
| Abuse (ref: No) | Yes | -0.05 | 0.08 | (-0.21,0.12) | 0.567 |
| Outsider growing up (ref: No) | Yes | -0.20 | 0.10 | (-0.39,-0.01) | 0.040 |
| Self-rated health growing up (ref: Good) | Excellent | 0.29 | 0.08 | (0.13,0.45) | <.001 |
|  | Very good | 0.02 | 0.09 | (-0.15,0.19) |  |
|  | Fair | -0.18 | 0.15 | (-0.48,0.12) |  |
|  | Poor | 0.01 | 0.27 | (-0.52,0.54) |  |
| Immigration status (ref: Born in this country) | Born in another country | 0.06 | 0.14 | (-0.22,0.34) | 0.650 |
| Age 12 religious service attendance (ref: Never) | At least 1/week | 0.15 | 0.08 | (-0.01,0.30) | 0.004 |
|  | 1-3/month | -0.10 | 0.10 | (-0.29,0.09) |  |
|  | <1/month | -0.11 | 0.10 | (-0.31,0.09) |  |
| Birth year (ref: 1998-2005; current age: 18-24) | 1993-1998; age 25-29 | 0.41 | 0.13 | (0.16,0.67) | 0.006 |
|  | 1983-1993; age 30-39 | 0.30 | 0.12 | (0.06,0.53) |  |
|  | 1973-1983; age 40-49 | 0.42 | 0.12 | (0.18,0.67) |  |
|  | 1963-1973; age 50-59 | 0.51 | 0.12 | (0.26,0.75) |  |
|  | 1953-1963; age 60-69 | 0.38 | 0.14 | (0.10,0.66) |  |
|  | 1943-1953; age 70-79 | 0.41 | 0.17 | (0.08,0.75) |  |
|  | 1943 or earlier; age 80+ | 0.11 | 0.26 | (-0.40,0.62) |  |
| Gender (ref:Male) | Female | 0.39 | 0.06 | (0.27,0.52) | <.001 |
|  | Other | 0.87 | 0.47 | (-0.05,1.80) |  |
| Religious affiliation at age 12 (ref: No religion/Atheist/Agnostic) | Christianity | 0.36 | 0.13 | (0.12,0.61) | <.001 |
|  | Collapsed affiliations with prevalence<3% | -0.46 | 0.38 | (-1.21,0.29) |  |
| Race/ethnicity (ref: plurality) | Race/ethnicity minority | 0.13 | 0.08 | (-0.02,0.28) | 0.031 |

***Supplementary Table S2C. E-Values for Estimates and CI for Argentina***

| **Variable** | **Category** | **E-Value for Estimate** | **E-Value for 95% CI** |
| --- | --- | --- | --- |
| Relationship with mother (ref: Very bad/Somewhat bad) | Very good/Somewhat good | 1.62 | 1.21 |
| Relationship with father (ref: Very bad/Somewhat bad) | Very good/Somewhat good | 1.10 | 1.00 |
| Parent marital status (ref: Married) | Divorced | 1.15 | 1.00 |
|  | Single, never married | 1.43 | 1.08 |
|  | One or both parents had died | 1.57 | 1.00 |
| Subjective financial status of family growing up (ref: Got by) | Lived comfortably | 1.17 | 1.00 |
|  | Found it difficult | 1.15 | 1.00 |
|  | Found it very difficult | 1.36 | 1.00 |
| Abuse (ref: No) | Yes | 1.18 | 1.00 |
| Outsider growing up (ref: No) | Yes | 1.44 | 1.06 |
| Self-rated health growing up (ref: Good) | Excellent | 1.59 | 1.34 |
|  | Very good | 1.11 | 1.00 |
|  | Fair | 1.41 | 1.00 |
|  | Poor | 1.08 | 1.00 |
| Immigration status (ref: Born in this country) | Born in another country | 1.21 | 1.00 |
| Age 12 religious service attendance (ref: Never) | At least 1/week | 1.36 | 1.00 |
|  | 1-3/month | 1.28 | 1.00 |
|  | <1/month | 1.30 | 1.00 |
| Birth year (ref: 1998-2005; current age: 18-24) | 1993-1998; age 25-29 | 1.76 | 1.38 |
|  | 1983-1993; age 30-39 | 1.59 | 1.20 |
|  | 1973-1983; age 40-49 | 1.77 | 1.41 |
|  | 1963-1973; age 50-59 | 1.90 | 1.54 |
|  | 1953-1963; age 60-69 | 1.71 | 1.28 |
|  | 1943-1953; age 70-79 | 1.76 | 1.25 |
|  | 1943 or earlier; age 80+ | 1.30 | 1.00 |
| Gender (ref:Male) | Female | 1.73 | 1.55 |
|  | Other | 2.46 | 1.00 |
| Religious affiliation at age 12 (ref: No religion/Atheist/Agnostic) | Christianity | 1.69 | 1.31 |
|  | Collapsed affiliations with prevalence<3% | 1.82 | 1.00 |
| Race/ethnicity (ref: plurality) | Race/ethnicity minority | 1.34 | 1.00 |

***Supplementary Table S3A. Nationally Representative Childhood Descriptive Statistics for Australia***

| **Variable** | **Category** | **N (%)** |
| --- | --- | --- |
| Relationship with mother | Very good | 2554 (66%) |
|  | Somewhat good | 925 (24%) |
|  | Somewhat bad | 218 (6%) |
|  | Very bad | 107 (3%) |
|  | Does not apply | 32 (1%) |
|  | Missing | 7 (0%) |
| Relationships with father | Very good | 2032 (53%) |
|  | Somewhat good | 1144 (30%) |
|  | Somewhat bad | 315 (8%) |
|  | Very bad | 196 (5%) |
|  | Does not apply | 148 (4%) |
|  | Missing | 9 (0%) |
| Parent marital status | Parents married | 3048 (79%) |
|  | Divorced | 462 (12%) |
|  | Single, never married | 187 (5%) |
|  | One or both parents had died | 96 (2%) |
|  | Missing | 52 (1%) |
| Subjective financial status of family growing up | Lived comfortably | 1756 (46%) |
|  | Got by | 1496 (39%) |
|  | Found it difficult | 422 (11%) |
|  | Found it very difficult | 154 (4%) |
|  | Missing | 16 (0%) |
| Abuse | Yes | 995 (26%) |
|  | No | 2790 (73%) |
|  | Missing | 59 (2%) |
| Outsider growing up | Yes | 756 (20%) |
|  | No | 3062 (80%) |
|  | Missing | 26 (1%) |
| Self-rated health growing up | Excellent | 1736 (45%) |
|  | Very good | 1087 (28%) |
|  | Good | 603 (16%) |
|  | Fair | 308 (8%) |
|  | Poor | 106 (3%) |
|  | Missing | 4 (0%) |
| Immigration status | Born in this country | 2953 (77%) |
|  | Born in another country | 885 (23%) |
|  | Missing | 6 (0%) |
| Age 12 religious service attendance | At least 1/week | 1362 (35%) |
|  | 1-3/month | 486 (13%) |
|  | <1/ month | 600 (16%) |
|  | Never | 1307 (34%) |
|  | Missing | 90 (2%) |
| Year of birth | 1998-2005; current age: 18-24 | 345 (9%) |
|  | 1993-1998; age 25-29 | 282 (7%) |
|  | 1983-1993; age 30-39 | 641 (17%) |
|  | 1973-1983; age 40-49 | 618 (16%) |
|  | 1963-1973; age 50-59 | 691 (18%) |
|  | 1953-1963; age 60-69 | 589 (15%) |
|  | 1943-1953; age 70-79 | 498 (13%) |
|  | 1943 or earlier; age 80+ | 178 (5%) |
|  | Missing | 2 (0%) |
| Gender | Male | 1861 (48%) |
|  | Female | 1941 (50%) |
|  | Other | 36 (1%) |
|  | Missing | 6 (0%) |
| Religious affiliation at age 12 | Christianity | 2678 (70%) |
|  | Islam | 48 (1%) |
|  | Hinduism | 39 (1%) |
|  | Buddhism | 16 (0%) |
|  | Judaism | 29 (1%) |
|  | Sikhism | 6 (0%) |
|  | Baha’i | 5 (0%) |
|  | Taoism | 1 (0%) |
|  | Primal, Animist, or Folk religion | 4 (0%) |
|  | Some other religion | 8 (0%) |
|  | No religion/Atheist/Agnostic | 990 (26%) |
|  | Missing | 21 (1%) |
| Race/ethnicity | Aboriginal | 53 (1%) |
|  | Australian | 1946 (51%) |
|  | Australian British/European | 1047 (27%) |
|  | Chinese | 75 (2%) |
|  | Indian | 58 (2%) |
|  | Japanese | 1 (0%) |
|  | Malay | 11 (0%) |
|  | Sinhalese | 1 (0%) |
|  | Spanish | 2 (0%) |
|  | Sri Lankan Moor | 1 (0%) |
|  | Sri Lankan Tamil | 7 (0%) |
|  | Vietnamese | 7 (0%) |
|  | Russian | 7 (0%) |
|  | Samoan | 4 (0%) |
|  | New Zealander | 91 (2%) |
|  | Other European | 357 (9%) |
|  | Other | 163 (4%) |
|  | Missing | 14 (0%) |

***Supplementary Table S3B. Regression of Optimism on Childhood Predictors for Australia***

| **Variable** | **Category** | **Estimate** | **SE** | **95% CI** | **Global P-value** |
| --- | --- | --- | --- | --- | --- |
| Relationship with mother (ref: Very bad/Somewhat bad) | Very good/Somewhat good | 0.01 | 0.20 | (-0.38,0.40) | 0.937 |
| Relationship with father (ref: Very bad/Somewhat bad) | Very good/Somewhat good | 0.18 | 0.17 | (-0.16,0.52) | 0.282 |
| Parent marital status (ref: Married) | Divorced | 0.08 | 0.19 | (-0.29,0.44) | 0.203 |
|  | Single, never married | 0.61 | 0.29 | (0.04,1.19) |  |
|  | One or both parents had died | 0.20 | 0.31 | (-0.41,0.80) |  |
| Subjective financial status of family growing up (ref: Got by) | Lived comfortably | 0.12 | 0.09 | (-0.07,0.30) | 0.075 |
|  | Found it difficult | -0.31 | 0.17 | (-0.65,0.03) |  |
|  | Found it very difficult | -0.06 | 0.34 | (-0.72,0.61) |  |
| Abuse (ref: No) | Yes | -0.53 | 0.12 | (-0.77,-0.30) | <.001 |
| Outsider growing up (ref: No) | Yes | -0.85 | 0.15 | (-1.15,-0.55) | <.001 |
| Self-rated health growing up (ref: Good) | Excellent | 0.69 | 0.13 | (0.43,0.95) | <.001 |
|  | Very good | 0.24 | 0.15 | (-0.04,0.53) |  |
|  | Fair | -0.48 | 0.25 | (-0.97,-0.00) |  |
|  | Poor | -0.85 | 0.37 | (-1.58,-0.12) |  |
| Immigration status (ref: Born in this country) | Born in another country | 0.23 | 0.12 | (0.01,0.46) | 0.041 |
| Age 12 religious service attendance (ref: Never) | At least 1/week | 0.31 | 0.13 | (0.06,0.56) | 0.092 |
|  | 1-3/month | 0.12 | 0.17 | (-0.21,0.44) |  |
|  | <1/month | 0.13 | 0.15 | (-0.16,0.42) |  |
| Birth year (ref: 1998-2005; current age: 18-24) | 1993-1998; age 25-29 | 0.34 | 0.27 | (-0.18,0.87) | <.001 |
|  | 1983-1993; age 30-39 | 0.57 | 0.23 | (0.12,1.01) |  |
|  | 1973-1983; age 40-49 | 0.70 | 0.22 | (0.26,1.14) |  |
|  | 1963-1973; age 50-59 | 0.74 | 0.21 | (0.32,1.16) |  |
|  | 1953-1963; age 60-69 | 0.97 | 0.21 | (0.56,1.39) |  |
|  | 1943-1953; age 70-79 | 1.20 | 0.22 | (0.76,1.64) |  |
|  | 1943 or earlier; age 80+ | 1.02 | 0.25 | (0.53,1.52) |  |
| Gender (ref:Male) | Female | 0.38 | 0.09 | (0.20,0.56) | <.001 |
|  | Other | -0.71 | 0.63 | (-1.95,0.53) |  |
| Religious affiliation at age 12 (ref: No religion/Atheist/Agnostic) | Christianity | 0.08 | 0.14 | (-0.19,0.35) | 0.646 |
|  | Collapsed affiliations with prevalence<3% | -0.11 | 0.27 | (-0.65,0.42) |  |
| Race/ethnicity (ref: plurality) | Race/ethnicity minority | 0.20 | 0.10 | (0.00,0.41) | 0.046 |

***Supplementary Table S3C. E-Values for Estimates and CI for Australia***

| **Variable** | **Category** | **E-Value for Estimate** | **E-Value for 95% CI** |
| --- | --- | --- | --- |
| Relationship with mother (ref: Very bad/Somewhat bad) | Very good/Somewhat good | 1.07 | 1.00 |
| Relationship with father (ref: Very bad/Somewhat bad) | Very good/Somewhat good | 1.36 | 1.00 |
| Parent marital status (ref: Married) | Divorced | 1.21 | 1.00 |
|  | Single, never married | 1.88 | 1.15 |
|  | One or both parents had died | 1.38 | 1.00 |
| Subjective financial status of family growing up (ref: Got by) | Lived comfortably | 1.27 | 1.00 |
|  | Found it difficult | 1.52 | 1.00 |
|  | Found it very difficult | 1.18 | 1.00 |
| Abuse (ref: No) | Yes | 1.78 | 1.51 |
| Outsider growing up (ref: No) | Yes | 2.16 | 1.81 |
| Self-rated health growing up (ref: Good) | Excellent | 1.96 | 1.66 |
|  | Very good | 1.44 | 1.00 |
|  | Fair | 1.72 | 1.02 |
|  | Poor | 2.16 | 1.28 |
| Immigration status (ref: Born in this country) | Born in another country | 1.42 | 1.05 |
| Age 12 religious service attendance (ref: Never) | At least 1/week | 1.52 | 1.17 |
|  | 1-3/month | 1.27 | 1.00 |
|  | <1/month | 1.29 | 1.00 |
| Birth year (ref: 1998-2005; current age: 18-24) | 1993-1998; age 25-29 | 1.56 | 1.00 |
|  | 1983-1993; age 30-39 | 1.82 | 1.28 |
|  | 1973-1983; age 40-49 | 1.97 | 1.46 |
|  | 1963-1973; age 50-59 | 2.02 | 1.53 |
|  | 1953-1963; age 60-69 | 2.32 | 1.81 |
|  | 1943-1953; age 70-79 | 2.61 | 2.05 |
|  | 1943 or earlier; age 80+ | 2.38 | 1.77 |
| Gender (ref:Male) | Female | 1.60 | 1.38 |
|  | Other | 1.99 | 1.00 |
| Religious affiliation at age 12 (ref: No religion/Atheist/Agnostic) | Christianity | 1.22 | 1.00 |
|  | Collapsed affiliations with prevalence<3% | 1.27 | 1.00 |
| Race/ethnicity (ref: plurality) | Race/ethnicity minority | 1.39 | 1.03 |

***Supplementary Table S4A. Nationally Representative Childhood Descriptive Statistics for Brazil***

| **Variable** | **Category** | **N (%)** |
| --- | --- | --- |
| Relationship with mother | Very good | 8369 (63%) |
|  | Somewhat good | 3559 (27%) |
|  | Somewhat bad | 483 (4%) |
|  | Very bad | 214 (2%) |
|  | Does not apply | 507 (4%) |
|  | Missing | 73 (1%) |
| Relationships with father | Very good | 6364 (48%) |
|  | Somewhat good | 3654 (28%) |
|  | Somewhat bad | 1035 (8%) |
|  | Very bad | 756 (6%) |
|  | Does not apply | 1303 (10%) |
|  | Missing | 93 (1%) |
| Parent marital status | Parents married | 8546 (65%) |
|  | Divorced | 1384 (10%) |
|  | Single, never married | 1985 (15%) |
|  | One or both parents had died | 508 (4%) |
|  | Missing | 781 (6%) |
| Subjective financial status of family growing up | Lived comfortably | 4998 (38%) |
|  | Got by | 4616 (35%) |
|  | Found it difficult | 2484 (19%) |
|  | Found it very difficult | 1027 (8%) |
|  | Missing | 79 (1%) |
| Abuse | Yes | 2606 (20%) |
|  | No | 10147 (77%) |
|  | Missing | 451 (3%) |
| Outsider growing up | Yes | 1659 (13%) |
|  | No | 11234 (85%) |
|  | Missing | 311 (2%) |
| Self-rated health growing up | Excellent | 5312 (40%) |
|  | Very good | 3392 (26%) |
|  | Good | 2873 (22%) |
|  | Fair | 1368 (10%) |
|  | Poor | 228 (2%) |
|  | Missing | 30 (0%) |
| Immigration status | Born in this country | 12688 (96%) |
|  | Born in another country | 153 (1%) |
|  | Missing | 363 (3%) |
| Age 12 religious service attendance | At least 1/week | 6306 (48%) |
|  | 1-3/month | 2491 (19%) |
|  | <1/ month | 2629 (20%) |
|  | Never | 1707 (13%) |
|  | Missing | 71 (1%) |
| Year of birth | 1998-2005; current age: 18-24 | 1986 (15%) |
|  | 1993-1998; age 25-29 | 1468 (11%) |
|  | 1983-1993; age 30-39 | 2908 (22%) |
|  | 1973-1983; age 40-49 | 2638 (20%) |
|  | 1963-1973; age 50-59 | 2131 (16%) |
|  | 1953-1963; age 60-69 | 1435 (11%) |
|  | 1943-1953; age 70-79 | 510 (4%) |
|  | 1943 or earlier; age 80+ | 126 (1%) |
| Gender | Male | 6320 (48%) |
|  | Female | 6820 (52%) |
|  | Other | 35 (0%) |
|  | Missing | 30 (0%) |
| Religious affiliation at age 12 | Christianity | 11403 (86%) |
|  | Islam | 15 (0%) |
|  | Hinduism | 1 (0%) |
|  | Buddhism | 27 (0%) |
|  | Judaism | 40 (0%) |
|  | Baha’i | 1 (0%) |
|  | Jainism | 4 (0%) |
|  | Shinto | 4 (0%) |
|  | Taoism | 1 (0%) |
|  | Confucianism | 7 (0%) |
|  | Primal, Animist, or Folk religion | 17 (0%) |
|  | Spiritism | 336 (3%) |
|  | Umbanda, Candomblé, and other African-derived religions | 262 (2%) |
|  | Some other religion | 87 (1%) |
|  | No religion/Atheist/Agnostic | 908 (7%) |
|  | Missing | 94 (1%) |
| Race/ethnicity | Branca | 5169 (39%) |
|  | Preta | 1615 (12%) |
|  | Parda | 5125 (39%) |
|  | Amarela | 238 (2%) |
|  | Indígena | 131 (1%) |
|  | Other | 61 (0%) |
|  | Missing | 865 (7%) |

***Supplementary Table S4B. Regression of Optimism on Childhood Predictors for Brazil***

| **Variable** | **Category** | **Estimate** | **SE** | **95% CI** | **Global P-value** |
| --- | --- | --- | --- | --- | --- |
| Relationship with mother (ref: Very bad/Somewhat bad) | Very good/Somewhat good | -0.02 | 0.07 | (-0.17,0.12) | 0.763 |
| Relationship with father (ref: Very bad/Somewhat bad) | Very good/Somewhat good | -0.00 | 0.06 | (-0.12,0.11) | 0.886 |
| Parent marital status (ref: Married) | Divorced | -0.10 | 0.07 | (-0.25,0.05) | 0.091 |
|  | Single, never married | -0.03 | 0.06 | (-0.15,0.10) |  |
|  | One or both parents had died | -0.27 | 0.14 | (-0.54,0.00) |  |
| Subjective financial status of family growing up (ref: Got by) | Lived comfortably | -0.02 | 0.04 | (-0.10,0.06) | 0.575 |
|  | Found it difficult | -0.07 | 0.06 | (-0.19,0.04) |  |
|  | Found it very difficult | 0.02 | 0.08 | (-0.14,0.19) |  |
| Abuse (ref: No) | Yes | -0.10 | 0.05 | (-0.20,0.00) | 0.059 |
| Outsider growing up (ref: No) | Yes | -0.39 | 0.07 | (-0.52,-0.26) | <.001 |
| Self-rated health growing up (ref: Good) | Excellent | 0.41 | 0.05 | (0.30,0.52) | <.001 |
|  | Very good | 0.15 | 0.06 | (0.04,0.27) |  |
|  | Fair | 0.03 | 0.08 | (-0.13,0.19) |  |
|  | Poor | 0.03 | 0.16 | (-0.29,0.34) |  |
| Immigration status (ref: Born in this country) | Born in another country | -0.30 | 0.16 | (-0.61,0.00) | 0.046 |
| Age 12 religious service attendance (ref: Never) | At least 1/week | 0.09 | 0.06 | (-0.04,0.21) | 0.072 |
|  | 1-3/month | -0.03 | 0.07 | (-0.17,0.12) |  |
|  | <1/month | -0.01 | 0.07 | (-0.15,0.13) |  |
| Birth year (ref: 1998-2005; current age: 18-24) | 1993-1998; age 25-29 | 0.14 | 0.07 | (-0.00,0.29) | <.001 |
|  | 1983-1993; age 30-39 | 0.21 | 0.06 | (0.09,0.33) |  |
|  | 1973-1983; age 40-49 | 0.25 | 0.07 | (0.12,0.38) |  |
|  | 1963-1973; age 50-59 | 0.39 | 0.07 | (0.26,0.52) |  |
|  | 1953-1963; age 60-69 | 0.36 | 0.08 | (0.20,0.52) |  |
|  | 1943-1953; age 70-79 | 0.24 | 0.13 | (-0.01,0.49) |  |
|  | 1943 or earlier; age 80+ | 0.33 | 0.23 | (-0.11,0.78) |  |
| Gender (ref:Male) | Female | 0.27 | 0.04 | (0.19,0.34) | <.001 |
|  | Other | -0.63 | 0.62 | (-1.84,0.58) |  |
| Religious affiliation at age 12 (ref: No religion/Atheist/Agnostic) | Christianity | 0.27 | 0.09 | (0.09,0.45) | 0.014 |
|  | Collapsed affiliations with prevalence<3% | 0.24 | 0.11 | (0.03,0.46) |  |
| Race/ethnicity (ref: plurality) | Race/ethnicity minority | 0.17 | 0.04 | (0.09,0.25) | <.001 |

***Supplementary Table S4C. E-Values for Estimates and CI for Brazil***

| **Variable** | **Category** | **E-Value for Estimate** | **E-Value for 95% CI** |
| --- | --- | --- | --- |
| Relationship with mother (ref: Very bad/Somewhat bad) | Very good/Somewhat good | 1.12 | 1.00 |
| Relationship with father (ref: Very bad/Somewhat bad) | Very good/Somewhat good | 1.05 | 1.00 |
| Parent marital status (ref: Married) | Divorced | 1.30 | 1.00 |
|  | Single, never married | 1.14 | 1.00 |
|  | One or both parents had died | 1.60 | 1.00 |
| Subjective financial status of family growing up (ref: Got by) | Lived comfortably | 1.11 | 1.00 |
|  | Found it difficult | 1.25 | 1.00 |
|  | Found it very difficult | 1.13 | 1.00 |
| Abuse (ref: No) | Yes | 1.31 | 1.00 |
| Outsider growing up (ref: No) | Yes | 1.80 | 1.58 |
| Self-rated health growing up (ref: Good) | Excellent | 1.83 | 1.65 |
|  | Very good | 1.40 | 1.17 |
|  | Fair | 1.14 | 1.00 |
|  | Poor | 1.14 | 1.00 |
| Immigration status (ref: Born in this country) | Born in another country | 1.65 | 1.00 |
| Age 12 religious service attendance (ref: Never) | At least 1/week | 1.28 | 1.00 |
|  | 1-3/month | 1.14 | 1.00 |
|  | <1/month | 1.08 | 1.00 |
| Birth year (ref: 1998-2005; current age: 18-24) | 1993-1998; age 25-29 | 1.39 | 1.00 |
|  | 1983-1993; age 30-39 | 1.50 | 1.29 |
|  | 1973-1983; age 40-49 | 1.56 | 1.35 |
|  | 1963-1973; age 50-59 | 1.79 | 1.58 |
|  | 1953-1963; age 60-69 | 1.75 | 1.49 |
|  | 1943-1953; age 70-79 | 1.55 | 1.00 |
|  | 1943 or earlier; age 80+ | 1.70 | 1.00 |
| Gender (ref:Male) | Female | 1.60 | 1.47 |
|  | Other | 2.20 | 1.00 |
| Religious affiliation at age 12 (ref: No religion/Atheist/Agnostic) | Christianity | 1.59 | 1.28 |
|  | Collapsed affiliations with prevalence<3% | 1.56 | 1.15 |
| Race/ethnicity (ref: plurality) | Race/ethnicity minority | 1.43 | 1.28 |

***Supplementary Table S5A. Nationally Representative Childhood Descriptive Statistics for Egypt***

| **Variable** | **Category** | **N (%)** |
| --- | --- | --- |
| Relationship with mother | Very good | 4110 (87%) |
|  | Somewhat good | 505 (11%) |
|  | Somewhat bad | 21 (0%) |
|  | Very bad | 10 (0%) |
|  | Does not apply | 83 (2%) |
| Relationships with father | Very good | 3713 (79%) |
|  | Somewhat good | 683 (14%) |
|  | Somewhat bad | 56 (1%) |
|  | Very bad | 30 (1%) |
|  | Does not apply | 233 (5%) |
|  | Missing | 14 (0%) |
| Parent marital status | Parents married | 4049 (86%) |
|  | Divorced | 131 (3%) |
|  | Single, never married | 9 (0%) |
|  | One or both parents had died | 485 (10%) |
|  | Missing | 55 (1%) |
| Subjective financial status of family growing up | Lived comfortably | 1251 (26%) |
|  | Got by | 2352 (50%) |
|  | Found it difficult | 857 (18%) |
|  | Found it very difficult | 268 (6%) |
|  | Missing | 1 (0%) |
| Abuse | Yes | 405 (9%) |
|  | No | 4293 (91%) |
|  | Missing | 30 (1%) |
| Outsider growing up | Yes | 260 (5%) |
|  | No | 4456 (94%) |
|  | Missing | 13 (0%) |
| Self-rated health growing up | Excellent | 2687 (57%) |
|  | Very good | 1174 (25%) |
|  | Good | 497 (11%) |
|  | Fair | 265 (6%) |
|  | Poor | 106 (2%) |
|  | Missing | 1 (0%) |
| Immigration status | Born in this country | 4713 (100%) |
|  | Born in another country | 16 (0%) |
|  | Missing | 1 (0%) |
| Age 12 religious service attendance | At least 1/week | 2307 (49%) |
|  | 1-3/month | 570 (12%) |
|  | <1/ month | 629 (13%) |
|  | Never | 1165 (25%) |
|  | Missing | 57 (1%) |
| Year of birth | 1998-2005; current age: 18-24 | 960 (20%) |
|  | 1993-1998; age 25-29 | 607 (13%) |
|  | 1983-1993; age 30-39 | 1204 (25%) |
|  | 1973-1983; age 40-49 | 897 (19%) |
|  | 1963-1973; age 50-59 | 613 (13%) |
|  | 1953-1963; age 60-69 | 387 (8%) |
|  | 1943-1953; age 70-79 | 54 (1%) |
|  | 1943 or earlier; age 80+ | 7 (0%) |
| Gender | Male | 2394 (51%) |
|  | Female | 2334 (49%) |
|  | Missing | 0 (0%) |
| Religious affiliation at age 12 | Christianity | 123 (3%) |
|  | Islam | 4602 (97%) |
|  | Jainism | 1 (0%) |
|  | Taoism | 0 (0%) |
|  | Missing | 3 (0%) |
| Race/ethnicity | Arab | 4585 (97%) |
|  | Turkish | 9 (0%) |
|  | Greek | 1 (0%) |
|  | Bedouin Arab | 4 (0%) |
|  | Nubian | 27 (1%) |
|  | Missing | 102 (2%) |

***Supplementary Table S5B. Regression of Optimism on Childhood Predictors for Egypt***

| **Variable** | **Category** | **Estimate** | **SE** | **95% CI** | **Global P-value** |
| --- | --- | --- | --- | --- | --- |
| Relationship with mother (ref: Very bad/Somewhat bad) | Very good/Somewhat good | 0.46 | 0.50 | (-0.53,1.44) | 0.361 |
| Relationship with father (ref: Very bad/Somewhat bad) | Very good/Somewhat good | 0.37 | 0.31 | (-0.24,0.99) | 0.228 |
| Parent marital status (ref: Married) | Divorced | 0.00 | 0.27 | (-0.54,0.55) | 0.121 |
|  | Single, never married | 1.21 | 0.52 | (0.19,2.23) |  |
|  | One or both parents had died | 0.12 | 0.16 | (-0.19,0.43) |  |
| Subjective financial status of family growing up (ref: Got by) | Lived comfortably | 0.12 | 0.10 | (-0.08,0.33) | 0.092 |
|  | Found it difficult | -0.21 | 0.13 | (-0.47,0.06) |  |
|  | Found it very difficult | -0.21 | 0.19 | (-0.59,0.16) |  |
| Abuse (ref: No) | Yes | -0.27 | 0.19 | (-0.65,0.11) | 0.143 |
| Outsider growing up (ref: No) | Yes | -0.08 | 0.20 | (-0.46,0.31) | 0.697 |
| Self-rated health growing up (ref: Good) | Excellent | 0.14 | 0.21 | (-0.27,0.55) | 0.234 |
|  | Very good | 0.03 | 0.21 | (-0.38,0.43) |  |
|  | Fair | -0.32 | 0.29 | (-0.89,0.25) |  |
|  | Poor | 0.32 | 0.36 | (-0.39,1.03) |  |
| Immigration status (ref: Born in this country) | Born in another country | 0.76 | 0.50 | (-0.24,1.75) | 0.134 |
| Age 12 religious service attendance (ref: Never) | At least 1/week | 0.28 | 0.12 | (0.03,0.52) | 0.075 |
|  | 1-3/month | 0.24 | 0.16 | (-0.09,0.56) |  |
|  | <1/month | 0.05 | 0.15 | (-0.24,0.35) |  |
| Birth year (ref: 1998-2005; current age: 18-24) | 1993-1998; age 25-29 | -0.34 | 0.18 | (-0.70,0.03) | 0.007 |
|  | 1983-1993; age 30-39 | -0.20 | 0.13 | (-0.46,0.07) |  |
|  | 1973-1983; age 40-49 | -0.02 | 0.15 | (-0.31,0.28) |  |
|  | 1963-1973; age 50-59 | 0.30 | 0.17 | (-0.05,0.64) |  |
|  | 1953-1963; age 60-69 | 0.17 | 0.21 | (-0.24,0.58) |  |
|  | 1943-1953; age 70-79 | 0.13 | 0.46 | (-0.79,1.05) |  |
|  | 1943 or earlier; age 80+ | 0.61 | 1.03 | (-1.43,2.65) |  |
| Gender (ref:Male) | Female | 0.31 | 0.11 | (0.09,0.53) | 0.005 |
| Religious affiliation at age 12 (ref: Islam) | Collapsed affiliations with prevalence<3% | -0.65 | 0.27 | (-1.19,-0.11) | 0.019 |
| Race/ethnicity (ref: plurality) | Race/ethnicity minority | 0.12 | 0.51 | (-0.90,1.14) | 0.628 |

***Supplementary Table S5C. E-Values for Estimates and CI for Egypt***

| **Variable** | **Category** | **E-Value for Estimate** | **E-Value for 95% CI** |
| --- | --- | --- | --- |
| Relationship with mother (ref: Very bad/Somewhat bad) | Very good/Somewhat good | 1.63 | 1.00 |
| Relationship with father (ref: Very bad/Somewhat bad) | Very good/Somewhat good | 1.54 | 1.00 |
| Parent marital status (ref: Married) | Divorced | 1.03 | 1.00 |
|  | Single, never married | 2.44 | 1.35 |
|  | One or both parents had died | 1.26 | 1.00 |
| Subjective financial status of family growing up (ref: Got by) | Lived comfortably | 1.26 | 1.00 |
|  | Found it difficult | 1.36 | 1.00 |
|  | Found it very difficult | 1.37 | 1.00 |
| Abuse (ref: No) | Yes | 1.43 | 1.00 |
| Outsider growing up (ref: No) | Yes | 1.19 | 1.00 |
| Self-rated health growing up (ref: Good) | Excellent | 1.28 | 1.00 |
|  | Very good | 1.11 | 1.00 |
|  | Fair | 1.49 | 1.00 |
|  | Poor | 1.49 | 1.00 |
| Immigration status (ref: Born in this country) | Born in another country | 1.94 | 1.00 |
| Age 12 religious service attendance (ref: Never) | At least 1/week | 1.44 | 1.12 |
|  | 1-3/month | 1.40 | 1.00 |
|  | <1/month | 1.16 | 1.00 |
| Birth year (ref: 1998-2005; current age: 18-24) | 1993-1998; age 25-29 | 1.50 | 1.00 |
|  | 1983-1993; age 30-39 | 1.35 | 1.00 |
|  | 1973-1983; age 40-49 | 1.08 | 1.00 |
|  | 1963-1973; age 50-59 | 1.46 | 1.00 |
|  | 1953-1963; age 60-69 | 1.32 | 1.00 |
|  | 1943-1953; age 70-79 | 1.27 | 1.00 |
|  | 1943 or earlier; age 80+ | 1.79 | 1.00 |
| Gender (ref:Male) | Female | 1.47 | 1.22 |
| Religious affiliation at age 12 (ref: Islam) | Collapsed affiliations with prevalence<3% | 1.83 | 1.25 |
| Race/ethnicity (ref: plurality) | Race/ethnicity minority | 1.26 | 1.00 |

***Supplementary Table S6A. Nationally Representative Childhood Descriptive Statistics for Germany***

| **Variable** | **Category** | **N (%)** |
| --- | --- | --- |
| Relationship with mother | Very good | 5497 (58%) |
|  | Somewhat good | 3031 (32%) |
|  | Somewhat bad | 496 (5%) |
|  | Very bad | 187 (2%) |
|  | Does not apply | 241 (3%) |
|  | Missing | 54 (1%) |
| Relationships with father | Very good | 4652 (49%) |
|  | Somewhat good | 3012 (32%) |
|  | Somewhat bad | 846 (9%) |
|  | Very bad | 385 (4%) |
|  | Does not apply | 538 (6%) |
|  | Missing | 73 (1%) |
| Parent marital status | Parents married | 7620 (80%) |
|  | Divorced | 927 (10%) |
|  | Single, never married | 578 (6%) |
|  | One or both parents had died | 245 (3%) |
|  | Missing | 136 (1%) |
| Subjective financial status of family growing up | Lived comfortably | 3177 (33%) |
|  | Got by | 4508 (47%) |
|  | Found it difficult | 1481 (16%) |
|  | Found it very difficult | 314 (3%) |
|  | Missing | 26 (0%) |
| Abuse | Yes | 1086 (11%) |
|  | No | 8321 (88%) |
|  | Missing | 99 (1%) |
| Outsider growing up | Yes | 1105 (12%) |
|  | No | 8262 (87%) |
|  | Missing | 139 (1%) |
| Self-rated health growing up | Excellent | 2633 (28%) |
|  | Very good | 3518 (37%) |
|  | Good | 2582 (27%) |
|  | Fair | 612 (6%) |
|  | Poor | 134 (1%) |
|  | Missing | 26 (0%) |
| Immigration status | Born in this country | 8722 (92%) |
|  | Born in another country | 744 (8%) |
|  | Missing | 40 (0%) |
| Age 12 religious service attendance | At least 1/week | 1943 (20%) |
|  | 1-3/month | 1899 (20%) |
|  | <1/ month | 2887 (30%) |
|  | Never | 2749 (29%) |
|  | Missing | 27 (0%) |
| Year of birth | 1998-2005; current age: 18-24 | 829 (9%) |
|  | 1993-1998; age 25-29 | 774 (8%) |
|  | 1983-1993; age 30-39 | 1438 (15%) |
|  | 1973-1983; age 40-49 | 1494 (16%) |
|  | 1963-1973; age 50-59 | 1729 (18%) |
|  | 1953-1963; age 60-69 | 1915 (20%) |
|  | 1943-1953; age 70-79 | 1137 (12%) |
|  | 1943 or earlier; age 80+ | 190 (2%) |
| Gender | Male | 4641 (49%) |
|  | Female | 4843 (51%) |
|  | Other | 11 (0%) |
|  | Missing | 11 (0%) |
| Religious affiliation at age 12 | Christianity | 5751 (61%) |
|  | Islam | 350 (4%) |
|  | Hinduism | 15 (0%) |
|  | Buddhism | 25 (0%) |
|  | Judaism | 18 (0%) |
|  | Sikhism | 5 (0%) |
|  | Baha’i | 2 (0%) |
|  | Jainism | 1 (0%) |
|  | Confucianism | 4 (0%) |
|  | Primal, Animist, or Folk religion | 19 (0%) |
|  | Some other religion | 67 (1%) |
|  | No religion/Atheist/Agnostic | 3163 (33%) |
|  | Missing | 85 (1%) |

***Supplementary Table S6B. Regression of Optimism on Childhood Predictors for Germany***

| **Variable** | **Category** | **Estimate** | **SE** | **95% CI** | **Global P-value** |
| --- | --- | --- | --- | --- | --- |
| Relationship with mother (ref: Very bad/Somewhat bad) | Very good/Somewhat good | 0.19 | 0.12 | (-0.04,0.43) | 0.096 |
| Relationship with father (ref: Very bad/Somewhat bad) | Very good/Somewhat good | 0.12 | 0.09 | (-0.07,0.30) | 0.208 |
| Parent marital status (ref: Married) | Divorced | -0.20 | 0.10 | (-0.40,0.00) | 0.070 |
|  | Single, never married | -0.20 | 0.13 | (-0.45,0.05) |  |
|  | One or both parents had died | 0.13 | 0.16 | (-0.19,0.44) |  |
| Subjective financial status of family growing up (ref: Got by) | Lived comfortably | 0.18 | 0.06 | (0.05,0.30) | 0.002 |
|  | Found it difficult | -0.16 | 0.09 | (-0.33,0.01) |  |
|  | Found it very difficult | -0.24 | 0.18 | (-0.59,0.12) |  |
| Abuse (ref: No) | Yes | -0.31 | 0.09 | (-0.49,-0.12) | <.001 |
| Outsider growing up (ref: No) | Yes | -0.50 | 0.09 | (-0.67,-0.32) | <.001 |
| Self-rated health growing up (ref: Good) | Excellent | 0.62 | 0.08 | (0.46,0.77) | <.001 |
|  | Very good | 0.18 | 0.07 | (0.05,0.32) |  |
|  | Fair | -0.18 | 0.13 | (-0.44,0.07) |  |
|  | Poor | 0.10 | 0.29 | (-0.47,0.66) |  |
| Immigration status (ref: Born in this country) | Born in another country | 0.30 | 0.12 | (0.06,0.53) | 0.013 |
| Age 12 religious service attendance (ref: Never) | At least 1/week | 0.36 | 0.08 | (0.19,0.52) | <.001 |
|  | 1-3/month | 0.22 | 0.08 | (0.06,0.38) |  |
|  | <1/month | 0.21 | 0.07 | (0.06,0.35) |  |
| Birth year (ref: 1998-2005; current age: 18-24) | 1993-1998; age 25-29 | 0.20 | 0.13 | (-0.07,0.46) | 0.001 |
|  | 1983-1993; age 30-39 | 0.18 | 0.12 | (-0.06,0.42) |  |
|  | 1973-1983; age 40-49 | 0.09 | 0.12 | (-0.15,0.32) |  |
|  | 1963-1973; age 50-59 | 0.11 | 0.12 | (-0.12,0.34) |  |
|  | 1953-1963; age 60-69 | 0.25 | 0.12 | (0.02,0.49) |  |
|  | 1943-1953; age 70-79 | 0.26 | 0.13 | (0.01,0.52) |  |
|  | 1943 or earlier; age 80+ | 0.72 | 0.17 | (0.39,1.05) |  |
| Gender (ref:Male) | Female | 0.15 | 0.05 | (0.05,0.26) | 0.002 |
|  | Other | -2.00 | 1.08 | (-4.12,0.12) |  |
| Religious affiliation at age 12 (ref: No religion/Atheist/Agnostic) | Christianity | 0.02 | 0.06 | (-0.10,0.14) | 0.243 |
|  | Islam | -0.32 | 0.18 | (-0.68,0.03) |  |
|  | Collapsed affiliations with prevalence<3% | -0.13 | 0.33 | (-0.77,0.51) |  |

***Supplementary Table S6C. E-Values for Estimates and CI for Germany***

| **Variable** | **Category** | **E-Value for Estimate** | **E-Value for 95% CI** |
| --- | --- | --- | --- |
| Relationship with mother (ref: Very bad/Somewhat bad) | Very good/Somewhat good | 1.39 | 1.00 |
| Relationship with father (ref: Very bad/Somewhat bad) | Very good/Somewhat good | 1.29 | 1.00 |
| Parent marital status (ref: Married) | Divorced | 1.40 | 1.00 |
|  | Single, never married | 1.40 | 1.00 |
|  | One or both parents had died | 1.30 | 1.00 |
| Subjective financial status of family growing up (ref: Got by) | Lived comfortably | 1.37 | 1.17 |
|  | Found it difficult | 1.35 | 1.00 |
|  | Found it very difficult | 1.45 | 1.00 |
| Abuse (ref: No) | Yes | 1.54 | 1.30 |
| Outsider growing up (ref: No) | Yes | 1.78 | 1.56 |
| Self-rated health growing up (ref: Good) | Excellent | 1.93 | 1.73 |
|  | Very good | 1.38 | 1.17 |
|  | Fair | 1.38 | 1.00 |
|  | Poor | 1.25 | 1.00 |
| Immigration status (ref: Born in this country) | Born in another country | 1.53 | 1.19 |
| Age 12 religious service attendance (ref: Never) | At least 1/week | 1.61 | 1.40 |
|  | 1-3/month | 1.43 | 1.18 |
|  | <1/month | 1.41 | 1.19 |
| Birth year (ref: 1998-2005; current age: 18-24) | 1993-1998; age 25-29 | 1.40 | 1.00 |
|  | 1983-1993; age 30-39 | 1.38 | 1.00 |
|  | 1973-1983; age 40-49 | 1.24 | 1.00 |
|  | 1963-1973; age 50-59 | 1.28 | 1.00 |
|  | 1953-1963; age 60-69 | 1.47 | 1.10 |
|  | 1943-1953; age 70-79 | 1.49 | 1.06 |
|  | 1943 or earlier; age 80+ | 2.07 | 1.65 |
| Gender (ref:Male) | Female | 1.34 | 1.17 |
|  | Other | 4.16 | 1.00 |
| Religious affiliation at age 12 (ref: No religion/Atheist/Agnostic) | Christianity | 1.10 | 1.00 |
|  | Islam | 1.56 | 1.00 |
|  | Collapsed affiliations with prevalence<3% | 1.31 | 1.00 |

***Supplementary Table S7A. Nationally Representative Childhood Descriptive Statistics for Hong Kong***

| **Variable** | **Category** | **N (%)** |
| --- | --- | --- |
| Relationship with mother | Very good | 1077 (36%) |
|  | Somewhat good | 1164 (39%) |
|  | Somewhat bad | 293 (10%) |
|  | Very bad | 49 (2%) |
|  | Does not apply | 426 (14%) |
|  | Missing | 3 (0%) |
| Relationships with father | Very good | 868 (29%) |
|  | Somewhat good | 1089 (36%) |
|  | Somewhat bad | 393 (13%) |
|  | Very bad | 102 (3%) |
|  | Does not apply | 557 (19%) |
|  | Missing | 3 (0%) |
| Parent marital status | Parents married | 2752 (91%) |
|  | Divorced | 114 (4%) |
|  | Single, never married | 40 (1%) |
|  | One or both parents had died | 50 (2%) |
|  | Missing | 56 (2%) |
| Subjective financial status of family growing up | Lived comfortably | 906 (30%) |
|  | Got by | 1527 (51%) |
|  | Found it difficult | 473 (16%) |
|  | Found it very difficult | 84 (3%) |
|  | Missing | 22 (1%) |
| Abuse | Yes | 318 (11%) |
|  | No | 2688 (89%) |
|  | Missing | 5 (0%) |
| Outsider growing up | Yes | 664 (22%) |
|  | No | 2224 (74%) |
|  | Missing | 124 (4%) |
| Self-rated health growing up | Excellent | 545 (18%) |
|  | Very good | 1073 (36%) |
|  | Good | 863 (29%) |
|  | Fair | 426 (14%) |
|  | Poor | 91 (3%) |
|  | Missing | 13 (0%) |
| Immigration status | Born in this country | 2637 (88%) |
|  | Born in another country | 321 (11%) |
|  | Missing | 53 (2%) |
| Age 12 religious service attendance | At least 1/week | 432 (14%) |
|  | 1-3/month | 528 (18%) |
|  | <1/ month | 753 (25%) |
|  | Never | 1295 (43%) |
|  | Missing | 4 (0%) |
| Year of birth | 1998-2005; current age: 18-24 | 217 (7%) |
|  | 1993-1998; age 25-29 | 198 (7%) |
|  | 1983-1993; age 30-39 | 507 (17%) |
|  | 1973-1983; age 40-49 | 580 (19%) |
|  | 1963-1973; age 50-59 | 711 (24%) |
|  | 1953-1963; age 60-69 | 620 (21%) |
|  | 1943-1953; age 70-79 | 164 (5%) |
|  | 1943 or earlier; age 80+ | 15 (0%) |
| Gender | Male | 1390 (46%) |
|  | Female | 1620 (54%) |
|  | Other | 2 (0%) |
| Religious affiliation at age 12 | Christianity | 715 (24%) |
|  | Islam | 86 (3%) |
|  | Hinduism | 27 (1%) |
|  | Buddhism | 323 (11%) |
|  | Judaism | 16 (1%) |
|  | Sikhism | 4 (0%) |
|  | Jainism | 1 (0%) |
|  | Shinto | 18 (1%) |
|  | Taoism | 81 (3%) |
|  | Confucianism | 10 (0%) |
|  | Primal, Animist, or Folk religion | 15 (0%) |
|  | Chinese folk/traditional religion | 108 (4%) |
|  | Some other religion | 5 (0%) |
|  | No religion/Atheist/Agnostic | 1601 (53%) |
|  | Missing | 1 (0%) |
| Race/ethnicity | Chinese (Cantonese) | 1930 (64%) |
|  | Chinese (Chaoshan) | 201 (7%) |
|  | Chinese (Fujianese) | 117 (4%) |
|  | Chinese (Hakka) | 121 (4%) |
|  | Chinese (Shanghainese) | 89 (3%) |
|  | Chinese (Other ethnicity) | 264 (9%) |
|  | East Asian (Korean, Japanese) | 10 (0%) |
|  | Southeast Asian (Filipino, Indonesian, Thailand) | 46 (2%) |
|  | South Asian (Indian, Nepalese, Pakistani) | 17 (1%) |
|  | Taiwanese | 14 (0%) |
|  | White | 15 (0%) |
|  | Other | 4 (0%) |
|  | Missing | 184 (6%) |

***Supplementary Table S7B. Regression of Optimism on Childhood Predictors for Hong Kong***

| **Variable** | **Category** | **Estimate** | **SE** | **95% CI** | **Global P-value** |
| --- | --- | --- | --- | --- | --- |
| Relationship with mother (ref: Very bad/Somewhat bad) | Very good/Somewhat good | 0.22 | 0.14 | (-0.05,0.50) | 0.114 |
| Relationship with father (ref: Very bad/Somewhat bad) | Very good/Somewhat good | 0.02 | 0.14 | (-0.25,0.29) | 0.895 |
| Parent marital status (ref: Married) | Divorced | -0.14 | 0.33 | (-0.79,0.51) | 0.650 |
|  | Single, never married | 0.02 | 0.36 | (-0.70,0.73) |  |
|  | One or both parents had died | -0.60 | 0.50 | (-1.59,0.38) |  |
| Subjective financial status of family growing up (ref: Got by) | Lived comfortably | 0.55 | 0.11 | (0.34,0.76) | <.001 |
|  | Found it difficult | -0.18 | 0.16 | (-0.49,0.13) |  |
|  | Found it very difficult | -0.19 | 0.43 | (-1.02,0.65) |  |
| Abuse (ref: No) | Yes | 0.05 | 0.17 | (-0.29,0.39) | 0.752 |
| Outsider growing up (ref: No) | Yes | -0.26 | 0.13 | (-0.51,-0.01) | 0.042 |
| Self-rated health growing up (ref: Good) | Excellent | 1.37 | 0.15 | (1.07,1.68) | <.001 |
|  | Very good | 0.63 | 0.12 | (0.40,0.86) |  |
|  | Fair | -0.88 | 0.15 | (-1.18,-0.58) |  |
|  | Poor | -1.93 | 0.51 | (-2.93,-0.93) |  |
| Immigration status (ref: Born in this country) | Born in another country | 0.06 | 0.20 | (-0.33,0.45) | 0.749 |
| Age 12 religious service attendance (ref: Never) | At least 1/week | 0.77 | 0.18 | (0.42,1.12) | <.001 |
|  | 1-3/month | 0.42 | 0.16 | (0.11,0.73) |  |
|  | <1/month | 0.14 | 0.12 | (-0.10,0.38) |  |
| Birth year (ref: 1998-2005; current age: 18-24) | 1993-1998; age 25-29 | -0.22 | 0.20 | (-0.61,0.17) | <.001 |
|  | 1983-1993; age 30-39 | -0.10 | 0.16 | (-0.41,0.21) |  |
|  | 1973-1983; age 40-49 | 0.18 | 0.15 | (-0.11,0.47) |  |
|  | 1963-1973; age 50-59 | 0.50 | 0.14 | (0.22,0.77) |  |
|  | 1953-1963; age 60-69 | 0.74 | 0.17 | (0.40,1.07) |  |
|  | 1943-1953; age 70-79 | 0.60 | 0.33 | (-0.06,1.25) |  |
|  | 1943 or earlier; age 80+ | -0.86 | 0.35 | (-1.56,-0.17) |  |
| Gender (ref:Male) | Female | 0.29 | 0.08 | (0.13,0.46) | 0.002 |
|  | Other | -0.24 | 0.73 | (-1.68,1.19) |  |
| Religious affiliation at age 12 (ref: No religion/Atheist/Agnostic) | Christianity | 0.20 | 0.15 | (-0.10,0.49) | 0.310 |
|  | Chinese folk/traditional religion | 0.12 | 0.29 | (-0.45,0.68) |  |
|  | Buddhism | 0.12 | 0.16 | (-0.20,0.45) |  |
|  | Collapsed affiliations with prevalence<3% | 0.41 | 0.19 | (0.03,0.78) |  |
| Race/ethnicity (ref: plurality) | Race/ethnicity minority | 0.04 | 0.10 | (-0.16,0.23) | 0.704 |

***Supplementary Table S7C. E-Values for Estimates and CI for Hong Kong***

| **Variable** | **Category** | **E-Value for Estimate** | **E-Value for 95% CI** |
| --- | --- | --- | --- |
| Relationship with mother (ref: Very bad/Somewhat bad) | Very good/Somewhat good | 1.43 | 1.00 |
| Relationship with father (ref: Very bad/Somewhat bad) | Very good/Somewhat good | 1.09 | 1.00 |
| Parent marital status (ref: Married) | Divorced | 1.31 | 1.00 |
|  | Single, never married | 1.09 | 1.00 |
|  | One or both parents had died | 1.92 | 1.00 |
| Subjective financial status of family growing up (ref: Got by) | Lived comfortably | 1.85 | 1.59 |
|  | Found it difficult | 1.38 | 1.00 |
|  | Found it very difficult | 1.38 | 1.00 |
| Abuse (ref: No) | Yes | 1.17 | 1.00 |
| Outsider growing up (ref: No) | Yes | 1.48 | 1.05 |
| Self-rated health growing up (ref: Good) | Excellent | 3.03 | 2.56 |
|  | Very good | 1.95 | 1.66 |
|  | Fair | 2.29 | 1.89 |
|  | Poor | 4.04 | 2.36 |
| Immigration status (ref: Born in this country) | Born in another country | 1.18 | 1.00 |
| Age 12 religious service attendance (ref: Never) | At least 1/week | 2.14 | 1.69 |
|  | 1-3/month | 1.68 | 1.27 |
|  | <1/month | 1.32 | 1.00 |
| Birth year (ref: 1998-2005; current age: 18-24) | 1993-1998; age 25-29 | 1.44 | 1.00 |
|  | 1983-1993; age 30-39 | 1.26 | 1.00 |
|  | 1973-1983; age 40-49 | 1.37 | 1.00 |
|  | 1963-1973; age 50-59 | 1.79 | 1.43 |
|  | 1953-1963; age 60-69 | 2.09 | 1.66 |
|  | 1943-1953; age 70-79 | 1.91 | 1.00 |
|  | 1943 or earlier; age 80+ | 2.26 | 1.36 |
| Gender (ref:Male) | Female | 1.53 | 1.30 |
|  | Other | 1.46 | 1.00 |
| Religious affiliation at age 12 (ref: No religion/Atheist/Agnostic) | Christianity | 1.40 | 1.00 |
|  | Chinese folk/traditional religion | 1.28 | 1.00 |
|  | Buddhism | 1.30 | 1.00 |
|  | Collapsed affiliations with prevalence<3% | 1.67 | 1.14 |
| Race/ethnicity (ref: plurality) | Race/ethnicity minority | 1.14 | 1.00 |

***Supplementary Table S8A. Nationally Representative Childhood Descriptive Statistics for India***

| **Variable** | **Category** | **N (%)** |
| --- | --- | --- |
| Relationship with mother | Very good | 11465 (90%) |
|  | Somewhat good | 788 (6%) |
|  | Somewhat bad | 88 (1%) |
|  | Very bad | 73 (1%) |
|  | Does not apply | 269 (2%) |
|  | Missing | 82 (1%) |
| Relationships with father | Very good | 10923 (86%) |
|  | Somewhat good | 995 (8%) |
|  | Somewhat bad | 126 (1%) |
|  | Very bad | 100 (1%) |
|  | Does not apply | 481 (4%) |
|  | Missing | 141 (1%) |
| Parent marital status | Parents married | 5578 (44%) |
|  | Divorced | 236 (2%) |
|  | Single, never married | 1055 (8%) |
|  | One or both parents had died | 940 (7%) |
|  | Missing | 4956 (39%) |
| Subjective financial status of family growing up | Lived comfortably | 4946 (39%) |
|  | Got by | 3010 (24%) |
|  | Found it difficult | 2703 (21%) |
|  | Found it very difficult | 2035 (16%) |
|  | Missing | 70 (1%) |
| Abuse | Yes | 1468 (11%) |
|  | No | 10526 (82%) |
|  | Missing | 771 (6%) |
| Outsider growing up | Yes | 1926 (15%) |
|  | No | 10780 (84%) |
|  | Missing | 59 (0%) |
| Self-rated health growing up | Excellent | 2182 (17%) |
|  | Very good | 3882 (30%) |
|  | Good | 4028 (32%) |
|  | Fair | 2202 (17%) |
|  | Poor | 424 (3%) |
|  | Missing | 47 (0%) |
| Immigration status | Born in this country | 12629 (99%) |
|  | Born in another country | 110 (1%) |
|  | Missing | 26 (0%) |
| Age 12 religious service attendance | At least 1/week | 5288 (41%) |
|  | 1-3/month | 2959 (23%) |
|  | <1/ month | 2719 (21%) |
|  | Never | 1478 (12%) |
|  | Missing | 321 (3%) |
| Year of birth | 1998-2005; current age: 18-24 | 2543 (20%) |
|  | 1993-1998; age 25-29 | 1640 (13%) |
|  | 1983-1993; age 30-39 | 3109 (24%) |
|  | 1973-1983; age 40-49 | 2275 (18%) |
|  | 1963-1973; age 50-59 | 1574 (12%) |
|  | 1953-1963; age 60-69 | 1188 (9%) |
|  | 1943-1953; age 70-79 | 370 (3%) |
|  | 1943 or earlier; age 80+ | 67 (1%) |
| Gender | Male | 6473 (51%) |
|  | Female | 6292 (49%) |
| Religious affiliation at age 12 | Christianity | 254 (2%) |
|  | Islam | 1550 (12%) |
|  | Hinduism | 10417 (82%) |
|  | Buddhism | 180 (1%) |
|  | Sikhism | 126 (1%) |
|  | Jainism | 9 (0%) |
|  | Shinto | 4 (0%) |
|  | Primal, Animist, or Folk religion | 27 (0%) |
|  | Some other religion | 59 (0%) |
|  | No religion/Atheist/Agnostic | 7 (0%) |
|  | Missing | 131 (1%) |
| Race/ethnicity | General | 3538 (28%) |
|  | Other backward caste | 4177 (33%) |
|  | Schedule caste | 3599 (28%) |
|  | Schedule tribe | 1185 (9%) |
|  | Missing | 267 (2%) |

***Supplementary Table S8B. Regression of Optimism on Childhood Predictors for India***

| **Variable** | **Category** | **Estimate** | **SE** | **95% CI** | **Global P-value** |
| --- | --- | --- | --- | --- | --- |
| Relationship with mother (ref: Very bad/Somewhat bad) | Very good/Somewhat good | -0.20 | 0.26 | (-0.72,0.31) | 0.441 |
| Relationship with father (ref: Very bad/Somewhat bad) | Very good/Somewhat good | -0.02 | 0.23 | (-0.47,0.43) | 0.918 |
| Parent marital status (ref: Married) | Divorced | -0.50 | 0.19 | (-0.88,-0.12) | <.001 |
|  | Single, never married | -0.23 | 0.11 | (-0.46,0.00) |  |
|  | One or both parents had died | 0.45 | 0.16 | (0.09,0.80) |  |
| Subjective financial status of family growing up (ref: Got by) | Lived comfortably | -0.13 | 0.08 | (-0.29,0.02) | 0.181 |
|  | Found it difficult | -0.14 | 0.09 | (-0.31,0.03) |  |
|  | Found it very difficult | -0.18 | 0.11 | (-0.39,0.03) |  |
| Abuse (ref: No) | Yes | -0.26 | 0.10 | (-0.45,-0.06) | 0.008 |
| Outsider growing up (ref: No) | Yes | -0.21 | 0.10 | (-0.40,-0.02) | 0.032 |
| Self-rated health growing up (ref: Good) | Excellent | 0.04 | 0.12 | (-0.18,0.27) | 0.017 |
|  | Very good | -0.01 | 0.08 | (-0.17,0.14) |  |
|  | Fair | -0.29 | 0.10 | (-0.48,-0.10) |  |
|  | Poor | -0.09 | 0.18 | (-0.44,0.26) |  |
| Immigration status (ref: Born in this country) | Born in another country | -0.41 | 0.36 | (-1.11,0.28) | 0.231 |
| Age 12 religious service attendance (ref: Never) | At least 1/week | 0.09 | 0.10 | (-0.11,0.29) | 0.740 |
|  | 1-3/month | 0.04 | 0.12 | (-0.18,0.27) |  |
|  | <1/month | 0.11 | 0.12 | (-0.13,0.34) |  |
| Birth year (ref: 1998-2005; current age: 18-24) | 1993-1998; age 25-29 | -0.10 | 0.11 | (-0.32,0.13) | <.001 |
|  | 1983-1993; age 30-39 | -0.12 | 0.10 | (-0.32,0.07) |  |
|  | 1973-1983; age 40-49 | -0.12 | 0.11 | (-0.33,0.09) |  |
|  | 1963-1973; age 50-59 | 0.03 | 0.12 | (-0.21,0.28) |  |
|  | 1953-1963; age 60-69 | -0.55 | 0.15 | (-0.85,-0.25) |  |
|  | 1943-1953; age 70-79 | -0.42 | 0.21 | (-0.84,-0.01) |  |
|  | 1943 or earlier; age 80+ | -1.03 | 0.55 | (-2.12,0.05) |  |
| Gender (ref:Male) | Female | 0.17 | 0.06 | (0.04,0.29) | 0.008 |
| Religious affiliation at age 12 (ref: Hinduism) | Islam | -0.24 | 0.13 | (-0.49,0.01) | 0.113 |
|  | Collapsed affiliations with prevalence<3% | 0.10 | 0.14 | (-0.18,0.37) |  |
| Race/ethnicity (ref: plurality) | Race/ethnicity minority | 0.05 | 0.07 | (-0.09,0.20) | 0.465 |

***Supplementary Table S8C. E-Values for Estimates and CI for India***

| **Variable** | **Category** | **E-Value for Estimate** | **E-Value for 95% CI** |
| --- | --- | --- | --- |
| Relationship with mother (ref: Very bad/Somewhat bad) | Very good/Somewhat good | 1.33 | 1.00 |
| Relationship with father (ref: Very bad/Somewhat bad) | Very good/Somewhat good | 1.08 | 1.00 |
| Parent marital status (ref: Married) | Divorced | 1.62 | 1.24 |
|  | Single, never married | 1.35 | 1.00 |
|  | One or both parents had died | 1.57 | 1.25 |
| Subjective financial status of family growing up (ref: Got by) | Lived comfortably | 1.26 | 1.00 |
|  | Found it difficult | 1.26 | 1.00 |
|  | Found it very difficult | 1.31 | 1.00 |
| Abuse (ref: No) | Yes | 1.39 | 1.16 |
| Outsider growing up (ref: No) | Yes | 1.33 | 1.08 |
| Self-rated health growing up (ref: Good) | Excellent | 1.13 | 1.00 |
|  | Very good | 1.07 | 1.00 |
|  | Fair | 1.42 | 1.21 |
|  | Poor | 1.21 | 1.00 |
| Immigration status (ref: Born in this country) | Born in another country | 1.54 | 1.00 |
| Age 12 religious service attendance (ref: Never) | At least 1/week | 1.20 | 1.00 |
|  | 1-3/month | 1.13 | 1.00 |
|  | <1/month | 1.22 | 1.00 |
| Birth year (ref: 1998-2005; current age: 18-24) | 1993-1998; age 25-29 | 1.21 | 1.00 |
|  | 1983-1993; age 30-39 | 1.24 | 1.00 |
|  | 1973-1983; age 40-49 | 1.23 | 1.00 |
|  | 1963-1973; age 50-59 | 1.11 | 1.00 |
|  | 1953-1963; age 60-69 | 1.66 | 1.38 |
|  | 1943-1953; age 70-79 | 1.54 | 1.05 |
|  | 1943 or earlier; age 80+ | 2.11 | 1.00 |
| Gender (ref:Male) | Female | 1.29 | 1.13 |
| Religious affiliation at age 12 (ref: Hinduism) | Islam | 1.37 | 1.00 |
|  | Collapsed affiliations with prevalence<3% | 1.21 | 1.00 |
| Race/ethnicity (ref: plurality) | Race/ethnicity minority | 1.15 | 1.00 |

***Supplementary Table S9A. Nationally Representative Childhood Descriptive Statistics for Indonesia***

| **Variable** | **Category** | **N (%)** |
| --- | --- | --- |
| Relationship with mother | Very good | 6238 (89%) |
|  | Somewhat good | 583 (8%) |
|  | Somewhat bad | 50 (1%) |
|  | Very bad | 26 (0%) |
|  | Does not apply | 68 (1%) |
|  | Missing | 27 (0%) |
| Relationships with father | Very good | 6067 (87%) |
|  | Somewhat good | 628 (9%) |
|  | Somewhat bad | 68 (1%) |
|  | Very bad | 52 (1%) |
|  | Does not apply | 115 (2%) |
|  | Missing | 61 (1%) |
| Parent marital status | Parents married | 5557 (79%) |
|  | Divorced | 448 (6%) |
|  | Single, never married | 47 (1%) |
|  | One or both parents had died | 735 (11%) |
|  | Missing | 205 (3%) |
| Subjective financial status of family growing up | Lived comfortably | 3408 (49%) |
|  | Got by | 2955 (42%) |
|  | Found it difficult | 439 (6%) |
|  | Found it very difficult | 181 (3%) |
|  | Missing | 9 (0%) |
| Abuse | Yes | 486 (7%) |
|  | No | 6427 (92%) |
|  | Missing | 79 (1%) |
| Outsider growing up | Yes | 343 (5%) |
|  | No | 6639 (95%) |
|  | Missing | 10 (0%) |
| Self-rated health growing up | Excellent | 1246 (18%) |
|  | Very good | 1968 (28%) |
|  | Good | 2490 (36%) |
|  | Fair | 1233 (18%) |
|  | Poor | 55 (1%) |
|  | Missing | 1 (0%) |
| Immigration status | Born in this country | 6958 (100%) |
|  | Born in another country | 34 (0%) |
| Age 12 religious service attendance | At least 1/week | 5363 (77%) |
|  | 1-3/month | 973 (14%) |
|  | <1/ month | 329 (5%) |
|  | Never | 275 (4%) |
|  | Missing | 51 (1%) |
| Year of birth | 1998-2005; current age: 18-24 | 1216 (17%) |
|  | 1993-1998; age 25-29 | 849 (12%) |
|  | 1983-1993; age 30-39 | 1591 (23%) |
|  | 1973-1983; age 40-49 | 1576 (23%) |
|  | 1963-1973; age 50-59 | 1169 (17%) |
|  | 1953-1963; age 60-69 | 490 (7%) |
|  | 1943-1953; age 70-79 | 83 (1%) |
|  | 1943 or earlier; age 80+ | 17 (0%) |
| Gender | Male | 3461 (50%) |
|  | Female | 3513 (50%) |
|  | Other | 7 (0%) |
|  | Missing | 11 (0%) |
| Religious affiliation at age 12 | Christianity | 528 (8%) |
|  | Islam | 6373 (91%) |
|  | Hinduism | 75 (1%) |
|  | Buddhism | 5 (0%) |
|  | Jainism | 1 (0%) |
|  | Taoism | 0 (0%) |
|  | Confucianism | 1 (0%) |
|  | Primal, Animist, or Folk religion | 1 (0%) |
|  | No religion/Atheist/Agnostic | 2 (0%) |
|  | Missing | 8 (0%) |
| Race/ethnicity | Banjar/Melayu Banjar | 320 (5%) |
|  | Betawi | 251 (4%) |
|  | Bugis | 243 (3%) |
|  | Jawa | 2846 (41%) |
|  | Madura | 262 (4%) |
|  | Minangkabau | 273 (4%) |
|  | Sunda/Parahyangan | 1172 (17%) |
|  | Bali | 69 (1%) |
|  | Batak | 165 (2%) |
|  | Makasar | 91 (1%) |
|  | Other | 1262 (18%) |
|  | Missing | 38 (1%) |

***Supplementary Table S9B. Regression of Optimism on Childhood Predictors for Indonesia***

| **Variable** | **Category** | **Estimate** | **SE** | **95% CI** | **Global P-value** |
| --- | --- | --- | --- | --- | --- |
| Relationship with mother (ref: Very bad/Somewhat bad) | Very good/Somewhat good | -0.01 | 0.24 | (-0.47,0.45) | 0.895 |
| Relationship with father (ref: Very bad/Somewhat bad) | Very good/Somewhat good | -0.14 | 0.18 | (-0.50,0.21) | 0.421 |
| Parent marital status (ref: Married) | Divorced | -0.04 | 0.13 | (-0.29,0.20) | 0.126 |
|  | Single, never married | 0.12 | 0.32 | (-0.51,0.75) |  |
|  | One or both parents had died | -0.30 | 0.13 | (-0.56,-0.04) |  |
| Subjective financial status of family growing up (ref: Got by) | Lived comfortably | 0.14 | 0.05 | (0.04,0.24) | 0.002 |
|  | Found it difficult | -0.34 | 0.17 | (-0.66,-0.01) |  |
|  | Found it very difficult | -0.26 | 0.24 | (-0.73,0.20) |  |
| Abuse (ref: No) | Yes | -0.27 | 0.14 | (-0.54,0.01) | 0.055 |
| Outsider growing up (ref: No) | Yes | 0.01 | 0.13 | (-0.26,0.27) | 0.887 |
| Self-rated health growing up (ref: Good) | Excellent | 0.03 | 0.07 | (-0.11,0.18) | 0.393 |
|  | Very good | -0.02 | 0.06 | (-0.14,0.11) |  |
|  | Fair | 0.11 | 0.07 | (-0.03,0.26) |  |
|  | Poor | 0.34 | 0.38 | (-0.41,1.09) |  |
| Immigration status (ref: Born in this country) | Born in another country | 0.11 | 0.24 | (-0.36,0.58) | 0.637 |
| Age 12 religious service attendance (ref: Never) | At least 1/week | 0.22 | 0.17 | (-0.12,0.56) | 0.393 |
|  | 1-3/month | 0.15 | 0.19 | (-0.23,0.53) |  |
|  | <1/month | 0.23 | 0.18 | (-0.11,0.58) |  |
| Birth year (ref: 1998-2005; current age: 18-24) | 1993-1998; age 25-29 | 0.13 | 0.09 | (-0.05,0.30) | 0.650 |
|  | 1983-1993; age 30-39 | 0.09 | 0.08 | (-0.06,0.24) |  |
|  | 1973-1983; age 40-49 | 0.13 | 0.08 | (-0.03,0.29) |  |
|  | 1963-1973; age 50-59 | -0.01 | 0.11 | (-0.21,0.20) |  |
|  | 1953-1963; age 60-69 | 0.11 | 0.14 | (-0.16,0.37) |  |
|  | 1943-1953; age 70-79 | -0.03 | 0.23 | (-0.48,0.43) |  |
|  | 1943 or earlier; age 80+ | 0.36 | 0.49 | (-0.61,1.33) |  |
| Gender (ref:Male) | Female | 0.12 | 0.05 | (0.02,0.22) | 0.012 |
|  | Other | -1.88 | 1.13 | (-4.11,0.35) |  |
| Religious affiliation at age 12 (ref: Islam) | Christianity | 0.18 | 0.12 | (-0.05,0.42) | 0.276 |
|  | Collapsed affiliations with prevalence<3% | -0.16 | 0.37 | (-0.89,0.56) |  |
| Race/ethnicity (ref: plurality) | Race/ethnicity minority | -0.02 | 0.05 | (-0.12,0.08) | 0.684 |

***Supplementary Table S9C. E-Values for Estimates and CI for Indonesia***

| **Variable** | **Category** | **E-Value for Estimate** | **E-Value for 95% CI** |
| --- | --- | --- | --- |
| Relationship with mother (ref: Very bad/Somewhat bad) | Very good/Somewhat good | 1.09 | 1.00 |
| Relationship with father (ref: Very bad/Somewhat bad) | Very good/Somewhat good | 1.38 | 1.00 |
| Parent marital status (ref: Married) | Divorced | 1.18 | 1.00 |
|  | Single, never married | 1.33 | 1.00 |
|  | One or both parents had died | 1.62 | 1.17 |
| Subjective financial status of family growing up (ref: Got by) | Lived comfortably | 1.37 | 1.17 |
|  | Found it difficult | 1.68 | 1.07 |
|  | Found it very difficult | 1.57 | 1.00 |
| Abuse (ref: No) | Yes | 1.58 | 1.00 |
| Outsider growing up (ref: No) | Yes | 1.06 | 1.00 |
| Self-rated health growing up (ref: Good) | Excellent | 1.15 | 1.00 |
|  | Very good | 1.11 | 1.00 |
|  | Fair | 1.32 | 1.00 |
|  | Poor | 1.69 | 1.00 |
| Immigration status (ref: Born in this country) | Born in another country | 1.32 | 1.00 |
| Age 12 religious service attendance (ref: Never) | At least 1/week | 1.50 | 1.00 |
|  | 1-3/month | 1.38 | 1.00 |
|  | <1/month | 1.52 | 1.00 |
| Birth year (ref: 1998-2005; current age: 18-24) | 1993-1998; age 25-29 | 1.35 | 1.00 |
|  | 1983-1993; age 30-39 | 1.28 | 1.00 |
|  | 1973-1983; age 40-49 | 1.35 | 1.00 |
|  | 1963-1973; age 50-59 | 1.06 | 1.00 |
|  | 1953-1963; age 60-69 | 1.31 | 1.00 |
|  | 1943-1953; age 70-79 | 1.13 | 1.00 |
|  | 1943 or earlier; age 80+ | 1.72 | 1.00 |
| Gender (ref:Male) | Female | 1.33 | 1.11 |
|  | Other | 4.93 | 1.00 |
| Religious affiliation at age 12 (ref: Islam) | Christianity | 1.44 | 1.00 |
|  | Collapsed affiliations with prevalence<3% | 1.41 | 1.00 |
| Race/ethnicity (ref: plurality) | Race/ethnicity minority | 1.12 | 1.00 |

***Supplementary Table S10A. Nationally Representative Childhood Descriptive Statistics for Israel***

| **Variable** | **Category** | **N (%)** |
| --- | --- | --- |
| Relationship with mother | Very good | 2686 (73%) |
|  | Somewhat good | 793 (22%) |
|  | Somewhat bad | 110 (3%) |
|  | Very bad | 18 (0%) |
|  | Does not apply | 45 (1%) |
|  | Missing | 17 (0%) |
| Relationships with father | Very good | 2290 (62%) |
|  | Somewhat good | 912 (25%) |
|  | Somewhat bad | 234 (6%) |
|  | Very bad | 37 (1%) |
|  | Does not apply | 171 (5%) |
|  | Missing | 25 (1%) |
| Parent marital status | Parents married | 3172 (86%) |
|  | Divorced | 284 (8%) |
|  | Single, never married | 36 (1%) |
|  | One or both parents had died | 130 (4%) |
|  | Missing | 47 (1%) |
| Subjective financial status of family growing up | Lived comfortably | 923 (25%) |
|  | Got by | 1822 (50%) |
|  | Found it difficult | 667 (18%) |
|  | Found it very difficult | 239 (7%) |
|  | Missing | 17 (0%) |
| Outsider growing up | Yes | 371 (10%) |
|  | No | 3228 (88%) |
|  | Missing | 70 (2%) |
| Self-rated health growing up | Excellent | 1785 (49%) |
|  | Very good | 1284 (35%) |
|  | Good | 480 (13%) |
|  | Fair | 105 (3%) |
|  | Poor | 6 (0%) |
|  | Missing | 8 (0%) |
| Immigration status | Born in this country | 2796 (76%) |
|  | Born in another country | 868 (24%) |
|  | Missing | 5 (0%) |
| Age 12 religious service attendance | At least 1/week | 867 (24%) |
|  | 1-3/month | 435 (12%) |
|  | <1/ month | 810 (22%) |
|  | Never | 1539 (42%) |
|  | Missing | 17 (0%) |
| Year of birth | 1998-2005; current age: 18-24 | 553 (15%) |
|  | 1993-1998; age 25-29 | 407 (11%) |
|  | 1983-1993; age 30-39 | 666 (18%) |
|  | 1973-1983; age 40-49 | 616 (17%) |
|  | 1963-1973; age 50-59 | 542 (15%) |
|  | 1953-1963; age 60-69 | 469 (13%) |
|  | 1943-1953; age 70-79 | 336 (9%) |
|  | 1943 or earlier; age 80+ | 79 (2%) |
| Gender | Male | 1791 (49%) |
|  | Female | 1872 (51%) |
|  | Other | 0 (0%) |
|  | Missing | 6 (0%) |
| Religious affiliation at age 12 | Christianity | 60 (2%) |
|  | Islam | 647 (18%) |
|  | Judaism | 2873 (78%) |
|  | Sikhism | 1 (0%) |
|  | Baha’i | 1 (0%) |
|  | Primal, Animist, or Folk religion | 3 (0%) |
|  | Some other religion | 5 (0%) |
|  | No religion/Atheist/Agnostic | 69 (2%) |
|  | Missing | 10 (0%) |
| Race/ethnicity | Jewish | 2926 (80%) |
|  | Arab | 674 (18%) |
|  | Other | 39 (1%) |
|  | Missing | 30 (1%) |

***Supplementary Table S10B. Regression of Optimism on Childhood Predictors for Israel***

| **Variable** | **Category** | **Estimate** | **SE** | **95% CI** | **Global P-value** |
| --- | --- | --- | --- | --- | --- |
| Relationship with mother (ref: Very bad/Somewhat bad) | Very good/Somewhat good | -0.27 | 0.19 | (-0.65,0.11) | 0.135 |
| Relationship with father (ref: Very bad/Somewhat bad) | Very good/Somewhat good | 0.07 | 0.14 | (-0.21,0.35) | 0.598 |
| Parent marital status (ref: Married) | Divorced | -0.47 | 0.15 | (-0.76,-0.18) | 0.001 |
|  | Single, never married | -0.74 | 0.50 | (-1.78,0.31) |  |
|  | One or both parents had died | -0.41 | 0.23 | (-0.86,0.05) |  |
| Subjective financial status of family growing up (ref: Got by) | Lived comfortably | -0.09 | 0.09 | (-0.26,0.08) | 0.111 |
|  | Found it difficult | 0.03 | 0.10 | (-0.16,0.22) |  |
|  | Found it very difficult | -0.35 | 0.18 | (-0.71,0.01) |  |
| Outsider growing up (ref: No) | Yes | -0.05 | 0.11 | (-0.27,0.16) | 0.608 |
| Self-rated health growing up (ref: Good) | Excellent | 0.27 | 0.13 | (0.01,0.53) | 0.022 |
|  | Very good | 0.38 | 0.13 | (0.13,0.63) |  |
|  | Fair | 0.02 | 0.19 | (-0.37,0.40) |  |
|  | Poor | 1.25 | 1.10 | (-0.93,3.43) |  |
| Immigration status (ref: Born in this country) | Born in another country | -0.11 | 0.10 | (-0.31,0.09) | 0.275 |
| Age 12 religious service attendance (ref: Never) | At least 1/week | 0.23 | 0.13 | (-0.03,0.49) | <.001 |
|  | 1-3/month | 0.45 | 0.12 | (0.21,0.70) |  |
|  | <1/month | 0.34 | 0.10 | (0.15,0.54) |  |
| Birth year (ref: 1998-2005; current age: 18-24) | 1993-1998; age 25-29 | -0.08 | 0.17 | (-0.41,0.25) | 0.024 |
|  | 1983-1993; age 30-39 | -0.11 | 0.15 | (-0.40,0.18) |  |
|  | 1973-1983; age 40-49 | -0.07 | 0.15 | (-0.37,0.23) |  |
|  | 1963-1973; age 50-59 | -0.24 | 0.15 | (-0.53,0.05) |  |
|  | 1953-1963; age 60-69 | -0.33 | 0.16 | (-0.65,-0.01) |  |
|  | 1943-1953; age 70-79 | -0.65 | 0.21 | (-1.06,-0.23) |  |
|  | 1943 or earlier; age 80+ | -0.97 | 0.38 | (-1.72,-0.22) |  |
| Gender (ref:Male) | Female | 0.10 | 0.07 | (-0.04,0.25) | 0.165 |
|  | Other | -0.33 | 0.28 | (-0.88,0.22) |  |
| Religious affiliation at age 12 (ref: Judaism) | Islam | -0.90 | 0.34 | (-1.57,-0.23) | 0.027 |
|  | Collapsed affiliations with prevalence<3% | -0.15 | 0.21 | (-0.56,0.26) |  |
| Race/ethnicity (ref: plurality) | Race/ethnicity minority | -0.39 | 0.30 | (-0.98,0.21) | 0.195 |

***Supplementary Table S10C. E-Values for Estimates and CI for Israel***

| **Variable** | **Category** | **E-Value for Estimate** | **E-Value for 95% CI** |
| --- | --- | --- | --- |
| Relationship with mother (ref: Very bad/Somewhat bad) | Very good/Somewhat good | 1.56 | 1.00 |
| Relationship with father (ref: Very bad/Somewhat bad) | Very good/Somewhat good | 1.24 | 1.00 |
| Parent marital status (ref: Married) | Divorced | 1.86 | 1.43 |
|  | Single, never married | 2.28 | 1.00 |
|  | One or both parents had died | 1.76 | 1.00 |
| Subjective financial status of family growing up (ref: Got by) | Lived comfortably | 1.27 | 1.00 |
|  | Found it difficult | 1.13 | 1.00 |
|  | Found it very difficult | 1.68 | 1.00 |
| Outsider growing up (ref: No) | Yes | 1.20 | 1.00 |
| Self-rated health growing up (ref: Good) | Excellent | 1.56 | 1.08 |
|  | Very good | 1.72 | 1.34 |
|  | Fair | 1.11 | 1.00 |
|  | Poor | 3.20 | 1.00 |
| Immigration status (ref: Born in this country) | Born in another country | 1.30 | 1.00 |
| Age 12 religious service attendance (ref: Never) | At least 1/week | 1.51 | 1.00 |
|  | 1-3/month | 1.84 | 1.48 |
|  | <1/month | 1.67 | 1.38 |
| Birth year (ref: 1998-2005; current age: 18-24) | 1993-1998; age 25-29 | 1.25 | 1.00 |
|  | 1983-1993; age 30-39 | 1.30 | 1.00 |
|  | 1973-1983; age 40-49 | 1.24 | 1.00 |
|  | 1963-1973; age 50-59 | 1.52 | 1.00 |
|  | 1953-1963; age 60-69 | 1.65 | 1.08 |
|  | 1943-1953; age 70-79 | 2.13 | 1.51 |
|  | 1943 or earlier; age 80+ | 2.66 | 1.49 |
| Gender (ref:Male) | Female | 1.29 | 1.00 |
|  | Other | 1.65 | 1.00 |
| Religious affiliation at age 12 (ref: Judaism) | Islam | 2.55 | 1.52 |
|  | Collapsed affiliations with prevalence<3% | 1.37 | 1.00 |
| Race/ethnicity (ref: plurality) | Race/ethnicity minority | 1.73 | 1.00 |

***Supplementary Table S11A. Nationally Representative Childhood Descriptive Statistics for Japan***

| **Variable** | **Category** | **N (%)** |
| --- | --- | --- |
| Relationship with mother | Very good | 5630 (27%) |
|  | Somewhat good | 9461 (46%) |
|  | Somewhat bad | 2750 (13%) |
|  | Very bad | 799 (4%) |
|  | Does not apply | 1838 (9%) |
|  | Missing | 66 (0%) |
| Relationships with father | Very good | 4156 (20%) |
|  | Somewhat good | 9081 (44%) |
|  | Somewhat bad | 3446 (17%) |
|  | Very bad | 1223 (6%) |
|  | Does not apply | 2580 (13%) |
|  | Missing | 57 (0%) |
| Parent marital status | Parents married | 17713 (86%) |
|  | Divorced | 1127 (5%) |
|  | Single, never married | 591 (3%) |
|  | One or both parents had died | 754 (4%) |
|  | Missing | 359 (2%) |
| Subjective financial status of family growing up | Lived comfortably | 8320 (41%) |
|  | Got by | 8799 (43%) |
|  | Found it difficult | 2398 (12%) |
|  | Found it very difficult | 973 (5%) |
|  | Missing | 52 (0%) |
| Abuse | Yes | 1482 (7%) |
|  | No | 18964 (92%) |
|  | Missing | 96 (0%) |
| Outsider growing up | Yes | 1963 (10%) |
|  | No | 17136 (83%) |
|  | Missing | 1444 (7%) |
| Self-rated health growing up | Excellent | 2711 (13%) |
|  | Very good | 7106 (35%) |
|  | Good | 6689 (33%) |
|  | Fair | 3199 (16%) |
|  | Poor | 758 (4%) |
|  | Missing | 80 (0%) |
| Immigration status | Born in this country | 19548 (95%) |
|  | Born in another country | 158 (1%) |
|  | Missing | 837 (4%) |
| Age 12 religious service attendance | At least 1/week | 398 (2%) |
|  | 1-3/month | 883 (4%) |
|  | <1/ month | 5023 (24%) |
|  | Never | 14117 (69%) |
|  | Missing | 123 (1%) |
| Year of birth | 1998-2005; current age: 18-24 | 1589 (8%) |
|  | 1993-1998; age 25-29 | 806 (4%) |
|  | 1983-1993; age 30-39 | 2851 (14%) |
|  | 1973-1983; age 40-49 | 3363 (16%) |
|  | 1963-1973; age 50-59 | 3770 (18%) |
|  | 1953-1963; age 60-69 | 4118 (20%) |
|  | 1943-1953; age 70-79 | 3554 (17%) |
|  | 1943 or earlier; age 80+ | 493 (2%) |
| Gender | Male | 9847 (48%) |
|  | Female | 10602 (52%) |
|  | Other | 28 (0%) |
|  | Missing | 66 (0%) |
| Religious affiliation at age 12 | Christianity | 343 (2%) |
|  | Islam | 7 (0%) |
|  | Hinduism | 4 (0%) |
|  | Buddhism | 6536 (32%) |
|  | Baha’i | 7 (0%) |
|  | Jainism | 1 (0%) |
|  | Shinto | 382 (2%) |
|  | Taoism | 14 (0%) |
|  | Confucianism | 25 (0%) |
|  | Primal, Animist, or Folk religion | 13 (0%) |
|  | Some other religion | 46 (0%) |
|  | No religion/Atheist/Agnostic | 12950 (63%) |
|  | Missing | 215 (1%) |

***Supplementary Table S11B. Regression of Optimism on Childhood Predictors for Japan***

| **Variable** | **Category** | **Estimate** | **SE** | **95% CI** | **Global P-value** |
| --- | --- | --- | --- | --- | --- |
| Relationship with mother (ref: Very bad/Somewhat bad) | Very good/Somewhat good | 0.32 | 0.05 | (0.22,0.42) | <.001 |
| Relationship with father (ref: Very bad/Somewhat bad) | Very good/Somewhat good | 0.22 | 0.05 | (0.13,0.31) | <.001 |
| Parent marital status (ref: Married) | Divorced | 0.20 | 0.09 | (0.04,0.37) | 0.007 |
|  | Single, never married | -0.19 | 0.10 | (-0.39,0.01) |  |
|  | One or both parents had died | 0.13 | 0.10 | (-0.05,0.32) |  |
| Subjective financial status of family growing up (ref: Got by) | Lived comfortably | 0.38 | 0.04 | (0.31,0.46) | <.001 |
|  | Found it difficult | -0.06 | 0.06 | (-0.18,0.06) |  |
|  | Found it very difficult | -0.14 | 0.11 | (-0.36,0.07) |  |
| Abuse (ref: No) | Yes | 0.01 | 0.08 | (-0.15,0.17) | 0.896 |
| Outsider growing up (ref: No) | Yes | -0.18 | 0.07 | (-0.31,-0.04) | 0.011 |
| Self-rated health growing up (ref: Good) | Excellent | 1.19 | 0.06 | (1.07,1.31) | <.001 |
|  | Very good | 0.55 | 0.04 | (0.47,0.63) |  |
|  | Fair | -0.32 | 0.05 | (-0.43,-0.22) |  |
|  | Poor | -0.47 | 0.12 | (-0.71,-0.23) |  |
| Immigration status (ref: Born in this country) | Born in another country | 0.11 | 0.18 | (-0.25,0.47) | 0.482 |
| Age 12 religious service attendance (ref: Never) | At least 1/week | 0.50 | 0.13 | (0.24,0.76) | <.001 |
|  | 1-3/month | 0.53 | 0.08 | (0.36,0.69) |  |
|  | <1/month | 0.21 | 0.04 | (0.13,0.29) |  |
| Birth year (ref: 1998-2005; current age: 18-24) | 1993-1998; age 25-29 | -0.08 | 0.11 | (-0.30,0.13) | <.001 |
|  | 1983-1993; age 30-39 | 0.05 | 0.08 | (-0.12,0.21) |  |
|  | 1973-1983; age 40-49 | 0.13 | 0.08 | (-0.03,0.29) |  |
|  | 1963-1973; age 50-59 | 0.36 | 0.08 | (0.21,0.52) |  |
|  | 1953-1963; age 60-69 | 0.75 | 0.08 | (0.60,0.91) |  |
|  | 1943-1953; age 70-79 | 1.13 | 0.08 | (0.98,1.29) |  |
|  | 1943 or earlier; age 80+ | 1.35 | 0.12 | (1.11,1.58) |  |
| Gender (ref:Male) | Female | 0.32 | 0.03 | (0.26,0.39) | <.001 |
|  | Other | 0.48 | 0.43 | (-0.36,1.32) |  |
| Religious affiliation at age 12 (ref: No religion/Atheist/Agnostic) | Buddhism | 0.17 | 0.04 | (0.09,0.24) | <.001 |
|  | Collapsed affiliations with prevalence<3% | 0.19 | 0.10 | (-0.01,0.39) |  |

***Supplementary Table S11C. E-Values for Estimates and CI for Japan***

| **Variable** | **Category** | **E-Value for Estimate** | **E-Value for 95% CI** |
| --- | --- | --- | --- |
| Relationship with mother (ref: Very bad/Somewhat bad) | Very good/Somewhat good | 1.53 | 1.40 |
| Relationship with father (ref: Very bad/Somewhat bad) | Very good/Somewhat good | 1.41 | 1.29 |
| Parent marital status (ref: Married) | Divorced | 1.39 | 1.14 |
|  | Single, never married | 1.37 | 1.00 |
|  | One or both parents had died | 1.30 | 1.00 |
| Subjective financial status of family growing up (ref: Got by) | Lived comfortably | 1.60 | 1.52 |
|  | Found it difficult | 1.18 | 1.00 |
|  | Found it very difficult | 1.31 | 1.00 |
| Abuse (ref: No) | Yes | 1.06 | 1.00 |
| Outsider growing up (ref: No) | Yes | 1.35 | 1.14 |
| Self-rated health growing up (ref: Good) | Excellent | 2.60 | 2.45 |
|  | Very good | 1.80 | 1.71 |
|  | Fair | 1.54 | 1.41 |
|  | Poor | 1.71 | 1.42 |
| Immigration status (ref: Born in this country) | Born in another country | 1.27 | 1.00 |
| Age 12 religious service attendance (ref: Never) | At least 1/week | 1.74 | 1.43 |
|  | 1-3/month | 1.77 | 1.58 |
|  | <1/month | 1.40 | 1.29 |
| Birth year (ref: 1998-2005; current age: 18-24) | 1993-1998; age 25-29 | 1.22 | 1.00 |
|  | 1983-1993; age 30-39 | 1.16 | 1.00 |
|  | 1973-1983; age 40-49 | 1.29 | 1.00 |
|  | 1963-1973; age 50-59 | 1.58 | 1.40 |
|  | 1953-1963; age 60-69 | 2.04 | 1.86 |
|  | 1943-1953; age 70-79 | 2.53 | 2.33 |
|  | 1943 or earlier; age 80+ | 2.83 | 2.50 |
| Gender (ref:Male) | Female | 1.54 | 1.46 |
|  | Other | 1.72 | 1.00 |
| Religious affiliation at age 12 (ref: No religion/Atheist/Agnostic) | Buddhism | 1.34 | 1.24 |
|  | Collapsed affiliations with prevalence<3% | 1.37 | 1.00 |

***Supplementary Table S12A. Nationally Representative Childhood Descriptive Statistics for Kenya***

| **Variable** | **Category** | **N (%)** |
| --- | --- | --- |
| Relationship with mother | Very good | 9418 (83%) |
|  | Somewhat good | 1435 (13%) |
|  | Somewhat bad | 130 (1%) |
|  | Very bad | 100 (1%) |
|  | Does not apply | 240 (2%) |
|  | Missing | 66 (1%) |
| Relationships with father | Very good | 7958 (70%) |
|  | Somewhat good | 1896 (17%) |
|  | Somewhat bad | 216 (2%) |
|  | Very bad | 220 (2%) |
|  | Does not apply | 967 (8%) |
|  | Missing | 132 (1%) |
| Parent marital status | Parents married | 9238 (81%) |
|  | Divorced | 697 (6%) |
|  | Single, never married | 681 (6%) |
|  | One or both parents had died | 471 (4%) |
|  | Missing | 301 (3%) |
| Subjective financial status of family growing up | Lived comfortably | 3026 (27%) |
|  | Got by | 3279 (29%) |
|  | Found it difficult | 4071 (36%) |
|  | Found it very difficult | 994 (9%) |
|  | Missing | 19 (0%) |
| Abuse | Yes | 1300 (11%) |
|  | No | 10039 (88%) |
|  | Missing | 49 (0%) |
| Outsider growing up | Yes | 1223 (11%) |
|  | No | 10114 (89%) |
|  | Missing | 52 (0%) |
| Self-rated health growing up | Excellent | 4449 (39%) |
|  | Very good | 2598 (23%) |
|  | Good | 2582 (23%) |
|  | Fair | 1384 (12%) |
|  | Poor | 349 (3%) |
|  | Missing | 26 (0%) |
| Immigration status | Born in this country | 11270 (99%) |
|  | Born in another country | 117 (1%) |
|  | Missing | 2 (0%) |
| Age 12 religious service attendance | At least 1/week | 9189 (81%) |
|  | 1-3/month | 1687 (15%) |
|  | <1/ month | 236 (2%) |
|  | Never | 198 (2%) |
|  | Missing | 79 (1%) |
| Year of birth | 1998-2005; current age: 18-24 | 2868 (25%) |
|  | 1993-1998; age 25-29 | 2035 (18%) |
|  | 1983-1993; age 30-39 | 2564 (23%) |
|  | 1973-1983; age 40-49 | 1708 (15%) |
|  | 1963-1973; age 50-59 | 1072 (9%) |
|  | 1953-1963; age 60-69 | 710 (6%) |
|  | 1943-1953; age 70-79 | 360 (3%) |
|  | 1943 or earlier; age 80+ | 67 (1%) |
|  | Missing | 5 (0%) |
| Gender | Male | 5567 (49%) |
|  | Female | 5813 (51%) |
|  | Other | 2 (0%) |
|  | Missing | 7 (0%) |
| Religious affiliation at age 12 | Christianity | 10369 (91%) |
|  | Islam | 916 (8%) |
|  | Buddhism | 5 (0%) |
|  | Judaism | 6 (0%) |
|  | Sikhism | 0 (0%) |
|  | Baha’i | 3 (0%) |
|  | Jainism | 1 (0%) |
|  | Primal, Animist, or Folk religion | 13 (0%) |
|  | Some other religion | 0 (0%) |
|  | No religion/Atheist/Agnostic | 67 (1%) |
|  | Missing | 9 (0%) |
| Race/ethnicity | Luhya | 1943 (17%) |
|  | Luo | 1120 (10%) |
|  | Kalenjin | 1377 (12%) |
|  | Kamba | 1299 (11%) |
|  | Kikuyu | 2119 (19%) |
|  | Kisii | 789 (7%) |
|  | Maasai | 237 (2%) |
|  | Meru | 630 (6%) |
|  | Kenyan Somali/Somali | 396 (3%) |
|  | Miji Kenda tribes | 708 (6%) |
|  | Embu | 197 (2%) |
|  | Other | 548 (5%) |
|  | Missing | 27 (0%) |

***Supplementary Table S12B. Regression of Optimism on Childhood Predictors for Kenya***

| **Variable** | **Category** | **Estimate** | **SE** | **95% CI** | **Global P-value** |
| --- | --- | --- | --- | --- | --- |
| Relationship with mother (ref: Very bad/Somewhat bad) | Very good/Somewhat good | -0.05 | 0.15 | (-0.34,0.24) | 0.739 |
| Relationship with father (ref: Very bad/Somewhat bad) | Very good/Somewhat good | 0.05 | 0.12 | (-0.19,0.30) | 0.668 |
| Parent marital status (ref: Married) | Divorced | -0.23 | 0.15 | (-0.52,0.06) | 0.075 |
|  | Single, never married | -0.19 | 0.15 | (-0.49,0.10) |  |
|  | One or both parents had died | 0.22 | 0.12 | (-0.01,0.46) |  |
| Subjective financial status of family growing up (ref: Got by) | Lived comfortably | 0.03 | 0.07 | (-0.10,0.16) | 0.205 |
|  | Found it difficult | 0.07 | 0.06 | (-0.05,0.19) |  |
|  | Found it very difficult | -0.15 | 0.12 | (-0.38,0.08) |  |
| Abuse (ref: No) | Yes | -0.60 | 0.10 | (-0.80,-0.41) | <.001 |
| Outsider growing up (ref: No) | Yes | -0.17 | 0.09 | (-0.35,0.01) | 0.058 |
| Self-rated health growing up (ref: Good) | Excellent | 0.10 | 0.07 | (-0.03,0.23) | 0.398 |
|  | Very good | -0.01 | 0.08 | (-0.16,0.15) |  |
|  | Fair | -0.01 | 0.09 | (-0.19,0.17) |  |
|  | Poor | -0.07 | 0.21 | (-0.48,0.34) |  |
| Immigration status (ref: Born in this country) | Born in another country | -0.24 | 0.26 | (-0.75,0.28) | 0.364 |
| Age 12 religious service attendance (ref: Never) | At least 1/week | -0.09 | 0.18 | (-0.43,0.26) | 0.937 |
|  | 1-3/month | -0.06 | 0.20 | (-0.45,0.32) |  |
|  | <1/month | -0.05 | 0.25 | (-0.55,0.45) |  |
| Birth year (ref: 1998-2005; current age: 18-24) | 1993-1998; age 25-29 | -0.06 | 0.06 | (-0.18,0.06) | 0.389 |
|  | 1983-1993; age 30-39 | -0.05 | 0.06 | (-0.18,0.07) |  |
|  | 1973-1983; age 40-49 | -0.23 | 0.10 | (-0.43,-0.02) |  |
|  | 1963-1973; age 50-59 | -0.12 | 0.11 | (-0.33,0.10) |  |
|  | 1953-1963; age 60-69 | -0.21 | 0.15 | (-0.50,0.08) |  |
|  | 1943-1953; age 70-79 | -0.25 | 0.21 | (-0.66,0.17) |  |
|  | 1943 or earlier; age 80+ | -0.25 | 0.43 | (-1.09,0.58) |  |
| Gender (ref:Male) | Female | 0.08 | 0.05 | (-0.03,0.18) | <.001 |
|  | Other | 1.13 | 0.12 | (0.90,1.37) |  |
| Religious affiliation at age 12 (ref: Christianity) | Islam | -0.54 | 0.19 | (-0.91,-0.17) | 0.013 |
|  | Collapsed affiliations with prevalence<3% | 0.10 | 0.27 | (-0.43,0.64) |  |
| Race/ethnicity (ref: plurality) | Race/ethnicity minority | -0.05 | 0.09 | (-0.22,0.11) | 0.528 |

***Supplementary Table S12C. E-Values for Estimates and CI for Kenya***

| **Variable** | **Category** | **E-Value for Estimate** | **E-Value for 95% CI** |
| --- | --- | --- | --- |
| Relationship with mother (ref: Very bad/Somewhat bad) | Very good/Somewhat good | 1.16 | 1.00 |
| Relationship with father (ref: Very bad/Somewhat bad) | Very good/Somewhat good | 1.17 | 1.00 |
| Parent marital status (ref: Married) | Divorced | 1.43 | 1.00 |
|  | Single, never married | 1.39 | 1.00 |
|  | One or both parents had died | 1.42 | 1.00 |
| Subjective financial status of family growing up (ref: Got by) | Lived comfortably | 1.12 | 1.00 |
|  | Found it difficult | 1.20 | 1.00 |
|  | Found it very difficult | 1.32 | 1.00 |
| Abuse (ref: No) | Yes | 1.89 | 1.65 |
| Outsider growing up (ref: No) | Yes | 1.36 | 1.00 |
| Self-rated health growing up (ref: Good) | Excellent | 1.26 | 1.00 |
|  | Very good | 1.05 | 1.00 |
|  | Fair | 1.06 | 1.00 |
|  | Poor | 1.20 | 1.00 |
| Immigration status (ref: Born in this country) | Born in another country | 1.44 | 1.00 |
| Age 12 religious service attendance (ref: Never) | At least 1/week | 1.23 | 1.00 |
|  | 1-3/month | 1.19 | 1.00 |
|  | <1/month | 1.17 | 1.00 |
| Birth year (ref: 1998-2005; current age: 18-24) | 1993-1998; age 25-29 | 1.18 | 1.00 |
|  | 1983-1993; age 30-39 | 1.17 | 1.00 |
|  | 1973-1983; age 40-49 | 1.43 | 1.12 |
|  | 1963-1973; age 50-59 | 1.28 | 1.00 |
|  | 1953-1963; age 60-69 | 1.40 | 1.00 |
|  | 1943-1953; age 70-79 | 1.45 | 1.00 |
|  | 1943 or earlier; age 80+ | 1.46 | 1.00 |
| Gender (ref:Male) | Female | 1.21 | 1.00 |
|  | Other | 2.58 | 2.27 |
| Religious affiliation at age 12 (ref: Christianity) | Islam | 1.81 | 1.36 |
|  | Collapsed affiliations with prevalence<3% | 1.26 | 1.00 |
| Race/ethnicity (ref: plurality) | Race/ethnicity minority | 1.17 | 1.00 |

***Supplementary Table S13A. Nationally Representative Childhood Descriptive Statistics for Mexico***

| **Variable** | **Category** | **N (%)** |
| --- | --- | --- |
| Relationship with mother | Very good | 3912 (68%) |
|  | Somewhat good | 1340 (23%) |
|  | Somewhat bad | 177 (3%) |
|  | Very bad | 90 (2%) |
|  | Does not apply | 177 (3%) |
|  | Missing | 80 (1%) |
| Relationships with father | Very good | 3089 (53%) |
|  | Somewhat good | 1556 (27%) |
|  | Somewhat bad | 335 (6%) |
|  | Very bad | 267 (5%) |
|  | Does not apply | 470 (8%) |
|  | Missing | 60 (1%) |
| Parent marital status | Parents married | 3999 (69%) |
|  | Divorced | 341 (6%) |
|  | Single, never married | 827 (14%) |
|  | One or both parents had died | 176 (3%) |
|  | Missing | 432 (7%) |
| Subjective financial status of family growing up | Lived comfortably | 1775 (31%) |
|  | Got by | 1872 (32%) |
|  | Found it difficult | 1712 (30%) |
|  | Found it very difficult | 369 (6%) |
|  | Missing | 48 (1%) |
| Abuse | Yes | 905 (16%) |
|  | No | 4604 (80%) |
|  | Missing | 267 (5%) |
| Outsider growing up | Yes | 772 (13%) |
|  | No | 4897 (85%) |
|  | Missing | 107 (2%) |
| Self-rated health growing up | Excellent | 1860 (32%) |
|  | Very good | 1350 (23%) |
|  | Good | 1677 (29%) |
|  | Fair | 743 (13%) |
|  | Poor | 133 (2%) |
|  | Missing | 14 (0%) |
| Immigration status | Born in this country | 5517 (96%) |
|  | Born in another country | 108 (2%) |
|  | Missing | 151 (3%) |
| Age 12 religious service attendance | At least 1/week | 2514 (44%) |
|  | 1-3/month | 1162 (20%) |
|  | <1/ month | 1087 (19%) |
|  | Never | 944 (16%) |
|  | Missing | 69 (1%) |
| Year of birth | 1998-2005; current age: 18-24 | 986 (17%) |
|  | 1993-1998; age 25-29 | 623 (11%) |
|  | 1983-1993; age 30-39 | 1312 (23%) |
|  | 1973-1983; age 40-49 | 1027 (18%) |
|  | 1963-1973; age 50-59 | 873 (15%) |
|  | 1953-1963; age 60-69 | 611 (11%) |
|  | 1943-1953; age 70-79 | 277 (5%) |
|  | 1943 or earlier; age 80+ | 68 (1%) |
| Gender | Male | 2755 (48%) |
|  | Female | 2997 (52%) |
|  | Other | 3 (0%) |
|  | Missing | 21 (0%) |
| Religious affiliation at age 12 | Christianity | 5337 (92%) |
|  | Islam | 6 (0%) |
|  | Hinduism | 1 (0%) |
|  | Buddhism | 1 (0%) |
|  | Judaism | 8 (0%) |
|  | Sikhism | 4 (0%) |
|  | Baha’i | 1 (0%) |
|  | Shinto | 2 (0%) |
|  | Taoism | 5 (0%) |
|  | Primal, Animist, or Folk religion | 2 (0%) |
|  | Some other religion | 7 (0%) |
|  | No religion/Atheist/Agnostic | 328 (6%) |
|  | Missing | 74 (1%) |
| Race/ethnicity | White | 1116 (19%) |
|  | Mestizo | 2762 (48%) |
|  | Indigenous | 594 (10%) |
|  | Black | 108 (2%) |
|  | Mulatto | 63 (1%) |
|  | Other | 339 (6%) |
|  | Missing | 794 (14%) |

***Supplementary Table S13B. Regression of Optimism on Childhood Predictors for Mexico***

| **Variable** | **Category** | **Estimate** | **SE** | **95% CI** | **Global P-value** |
| --- | --- | --- | --- | --- | --- |
| Relationship with mother (ref: Very bad/Somewhat bad) | Very good/Somewhat good | 0.11 | 0.14 | (-0.16,0.37) | 0.431 |
| Relationship with father (ref: Very bad/Somewhat bad) | Very good/Somewhat good | 0.07 | 0.10 | (-0.11,0.26) | 0.436 |
| Parent marital status (ref: Married) | Divorced | 0.02 | 0.11 | (-0.20,0.24) | 0.099 |
|  | Single, never married | 0.15 | 0.08 | (-0.01,0.30) |  |
|  | One or both parents had died | 0.29 | 0.15 | (-0.01,0.59) |  |
| Subjective financial status of family growing up (ref: Got by) | Lived comfortably | -0.00 | 0.08 | (-0.15,0.15) | 0.036 |
|  | Found it difficult | 0.18 | 0.07 | (0.05,0.31) |  |
|  | Found it very difficult | 0.10 | 0.14 | (-0.17,0.38) |  |
| Abuse (ref: No) | Yes | -0.09 | 0.08 | (-0.25,0.06) | 0.216 |
| Outsider growing up (ref: No) | Yes | -0.14 | 0.09 | (-0.32,0.04) | 0.127 |
| Self-rated health growing up (ref: Good) | Excellent | 0.15 | 0.08 | (0.00,0.30) | <.001 |
|  | Very good | 0.13 | 0.07 | (-0.01,0.28) |  |
|  | Fair | -0.25 | 0.11 | (-0.46,-0.03) |  |
|  | Poor | -0.45 | 0.27 | (-0.98,0.08) |  |
| Immigration status (ref: Born in this country) | Born in another country | -0.11 | 0.21 | (-0.52,0.30) | 0.570 |
| Age 12 religious service attendance (ref: Never) | At least 1/week | 0.08 | 0.08 | (-0.08,0.25) | 0.226 |
|  | 1-3/month | -0.06 | 0.09 | (-0.23,0.11) |  |
|  | <1/month | -0.03 | 0.10 | (-0.22,0.15) |  |
| Birth year (ref: 1998-2005; current age: 18-24) | 1993-1998; age 25-29 | 0.05 | 0.10 | (-0.15,0.25) | 0.002 |
|  | 1983-1993; age 30-39 | 0.08 | 0.09 | (-0.10,0.27) |  |
|  | 1973-1983; age 40-49 | 0.25 | 0.10 | (0.05,0.46) |  |
|  | 1963-1973; age 50-59 | 0.37 | 0.09 | (0.19,0.56) |  |
|  | 1953-1963; age 60-69 | 0.20 | 0.11 | (-0.02,0.41) |  |
|  | 1943-1953; age 70-79 | 0.15 | 0.14 | (-0.12,0.41) |  |
|  | 1943 or earlier; age 80+ | -0.01 | 0.23 | (-0.46,0.43) |  |
| Gender (ref:Male) | Female | 0.20 | 0.06 | (0.08,0.31) | 0.001 |
|  | Other | -0.71 | 0.63 | (-1.95,0.52) |  |
| Religious affiliation at age 12 (ref: No religion/Atheist/Agnostic) | Christianity | 0.25 | 0.13 | (0.00,0.50) | 0.020 |
|  | Collapsed affiliations with prevalence<3% | 0.61 | 0.24 | (0.15,1.07) |  |
| Race/ethnicity (ref: plurality) | Race/ethnicity minority | 0.04 | 0.06 | (-0.08,0.15) | 0.515 |

***Supplementary Table S13C. E-Values for Estimates and CI for Mexico***

| **Variable** | **Category** | **E-Value for Estimate** | **E-Value for 95% CI** |
| --- | --- | --- | --- |
| Relationship with mother (ref: Very bad/Somewhat bad) | Very good/Somewhat good | 1.31 | 1.00 |
| Relationship with father (ref: Very bad/Somewhat bad) | Very good/Somewhat good | 1.25 | 1.00 |
| Parent marital status (ref: Married) | Divorced | 1.12 | 1.00 |
|  | Single, never married | 1.39 | 1.00 |
|  | One or both parents had died | 1.62 | 1.00 |
| Subjective financial status of family growing up (ref: Got by) | Lived comfortably | 1.02 | 1.00 |
|  | Found it difficult | 1.44 | 1.19 |
|  | Found it very difficult | 1.30 | 1.00 |
| Abuse (ref: No) | Yes | 1.29 | 1.00 |
| Outsider growing up (ref: No) | Yes | 1.37 | 1.00 |
| Self-rated health growing up (ref: Good) | Excellent | 1.40 | 1.05 |
|  | Very good | 1.36 | 1.00 |
|  | Fair | 1.55 | 1.15 |
|  | Poor | 1.88 | 1.00 |
| Immigration status (ref: Born in this country) | Born in another country | 1.32 | 1.00 |
| Age 12 religious service attendance (ref: Never) | At least 1/week | 1.27 | 1.00 |
|  | 1-3/month | 1.21 | 1.00 |
|  | <1/month | 1.16 | 1.00 |
| Birth year (ref: 1998-2005; current age: 18-24) | 1993-1998; age 25-29 | 1.20 | 1.00 |
|  | 1983-1993; age 30-39 | 1.27 | 1.00 |
|  | 1973-1983; age 40-49 | 1.56 | 1.19 |
|  | 1963-1973; age 50-59 | 1.75 | 1.46 |
|  | 1953-1963; age 60-69 | 1.47 | 1.00 |
|  | 1943-1953; age 70-79 | 1.38 | 1.00 |
|  | 1943 or earlier; age 80+ | 1.10 | 1.00 |
| Gender (ref:Male) | Female | 1.47 | 1.27 |
|  | Other | 2.32 | 1.00 |
| Religious affiliation at age 12 (ref: No religion/Atheist/Agnostic) | Christianity | 1.56 | 1.05 |
|  | Collapsed affiliations with prevalence<3% | 2.14 | 1.39 |
| Race/ethnicity (ref: plurality) | Race/ethnicity minority | 1.16 | 1.00 |

***Supplementary Table S14A. Nationally Representative Childhood Descriptive Statistics for Nigeria***

| **Variable** | **Category** | **N (%)** |
| --- | --- | --- |
| Relationship with mother | Very good | 5986 (88%) |
|  | Somewhat good | 648 (9%) |
|  | Somewhat bad | 62 (1%) |
|  | Very bad | 18 (0%) |
|  | Does not apply | 104 (2%) |
|  | Missing | 9 (0%) |
| Relationships with father | Very good | 5578 (82%) |
|  | Somewhat good | 924 (14%) |
|  | Somewhat bad | 76 (1%) |
|  | Very bad | 43 (1%) |
|  | Does not apply | 177 (3%) |
|  | Missing | 29 (0%) |
| Parent marital status | Parents married | 5568 (82%) |
|  | Divorced | 307 (5%) |
|  | Single, never married | 335 (5%) |
|  | One or both parents had died | 462 (7%) |
|  | Missing | 154 (2%) |
| Subjective financial status of family growing up | Lived comfortably | 2192 (32%) |
|  | Got by | 2381 (35%) |
|  | Found it difficult | 1661 (24%) |
|  | Found it very difficult | 563 (8%) |
|  | Missing | 29 (0%) |
| Abuse | Yes | 880 (13%) |
|  | No | 5851 (86%) |
|  | Missing | 96 (1%) |
| Outsider growing up | Yes | 669 (10%) |
|  | No | 6059 (89%) |
|  | Missing | 99 (1%) |
| Self-rated health growing up | Excellent | 2644 (39%) |
|  | Very good | 2613 (38%) |
|  | Good | 1152 (17%) |
|  | Fair | 306 (4%) |
|  | Poor | 98 (1%) |
|  | Missing | 14 (0%) |
| Immigration status | Born in this country | 6779 (99%) |
|  | Born in another country | 47 (1%) |
|  | Missing | 1 (0%) |
| Age 12 religious service attendance | At least 1/week | 5907 (87%) |
|  | 1-3/month | 600 (9%) |
|  | <1/ month | 136 (2%) |
|  | Never | 138 (2%) |
|  | Missing | 45 (1%) |
| Year of birth | 1998-2005; current age: 18-24 | 1533 (22%) |
|  | 1993-1998; age 25-29 | 1193 (17%) |
|  | 1983-1993; age 30-39 | 1943 (28%) |
|  | 1973-1983; age 40-49 | 1059 (16%) |
|  | 1963-1973; age 50-59 | 619 (9%) |
|  | 1953-1963; age 60-69 | 296 (4%) |
|  | 1943-1953; age 70-79 | 133 (2%) |
|  | 1943 or earlier; age 80+ | 50 (1%) |
| Gender | Male | 3371 (49%) |
|  | Female | 3456 (51%) |
|  | Other | 0 (0%) |
| Religious affiliation at age 12 | Christianity | 3463 (51%) |
|  | Islam | 3314 (49%) |
|  | Buddhism | 0 (0%) |
|  | Confucianism | 0 (0%) |
|  | Primal, Animist, or Folk religion | 17 (0%) |
|  | No religion/Atheist/Agnostic | 19 (0%) |
|  | Missing | 14 (0%) |
| Race/ethnicity | Hausa | 2342 (34%) |
|  | Yoruba | 1230 (18%) |
|  | Igbo (Ibo) | 1111 (16%) |
|  | Edo | 116 (2%) |
|  | Urhobo | 38 (1%) |
|  | Fulani | 266 (4%) |
|  | Kanuri | 31 (0%) |
|  | Tiv | 198 (3%) |
|  | Efik | 48 (1%) |
|  | Ijaw | 110 (2%) |
|  | Igala | 77 (1%) |
|  | Ibibio | 180 (3%) |
|  | Idoma | 61 (1%) |
|  | Other | 1014 (15%) |
|  | Missing | 4 (0%) |

***Supplementary Table S14B. Regression of Optimism on Childhood Predictors for Nigeria***

| **Variable** | **Category** | **Estimate** | **SE** | **95% CI** | **Global P-value** |
| --- | --- | --- | --- | --- | --- |
| Relationship with mother (ref: Very bad/Somewhat bad) | Very good/Somewhat good | -0.25 | 0.27 | (-0.79,0.28) | 0.355 |
| Relationship with father (ref: Very bad/Somewhat bad) | Very good/Somewhat good | -0.19 | 0.17 | (-0.52,0.14) | 0.246 |
| Parent marital status (ref: Married) | Divorced | -0.51 | 0.17 | (-0.84,-0.17) | <.001 |
|  | Single, never married | -0.32 | 0.13 | (-0.58,-0.06) |  |
|  | One or both parents had died | -0.58 | 0.16 | (-0.90,-0.26) |  |
| Subjective financial status of family growing up (ref: Got by) | Lived comfortably | -0.13 | 0.08 | (-0.29,0.04) | 0.221 |
|  | Found it difficult | -0.07 | 0.08 | (-0.23,0.10) |  |
|  | Found it very difficult | -0.24 | 0.13 | (-0.50,0.02) |  |
| Abuse (ref: No) | Yes | -0.26 | 0.12 | (-0.51,-0.02) | 0.033 |
| Outsider growing up (ref: No) | Yes | -0.11 | 0.11 | (-0.32,0.10) | 0.297 |
| Self-rated health growing up (ref: Good) | Excellent | -0.06 | 0.10 | (-0.25,0.13) | 0.858 |
|  | Very good | -0.06 | 0.09 | (-0.25,0.13) |  |
|  | Fair | -0.11 | 0.16 | (-0.43,0.21) |  |
|  | Poor | -0.19 | 0.22 | (-0.62,0.24) |  |
| Immigration status (ref: Born in this country) | Born in another country | 0.11 | 0.30 | (-0.49,0.71) | 0.717 |
| Age 12 religious service attendance (ref: Never) | At least 1/week | -0.19 | 0.17 | (-0.51,0.14) | 0.088 |
|  | 1-3/month | -0.17 | 0.19 | (-0.54,0.20) |  |
|  | <1/month | -0.56 | 0.23 | (-1.00,-0.11) |  |
| Birth year (ref: 1998-2005; current age: 18-24) | 1993-1998; age 25-29 | 0.00 | 0.09 | (-0.17,0.18) | 0.908 |
|  | 1983-1993; age 30-39 | 0.02 | 0.08 | (-0.14,0.18) |  |
|  | 1973-1983; age 40-49 | -0.02 | 0.11 | (-0.24,0.20) |  |
|  | 1963-1973; age 50-59 | 0.17 | 0.13 | (-0.09,0.44) |  |
|  | 1953-1963; age 60-69 | 0.14 | 0.19 | (-0.22,0.51) |  |
|  | 1943-1953; age 70-79 | -0.28 | 0.59 | (-1.44,0.88) |  |
|  | 1943 or earlier; age 80+ | -0.10 | 0.24 | (-0.58,0.38) |  |
| Gender (ref:Male) | Female | 0.10 | 0.06 | (-0.01,0.22) | 0.187 |
|  | Other | 0.14 | 0.16 | (-0.18,0.45) |  |
| Religious affiliation at age 12 (ref: Christianity) | Islam | -0.21 | 0.10 | (-0.41,-0.00) | 0.023 |
|  | Collapsed affiliations with prevalence<3% | 0.48 | 0.32 | (-0.15,1.12) |  |
| Race/ethnicity (ref: plurality) | Race/ethnicity minority | -0.07 | 0.11 | (-0.28,0.14) | 0.498 |

***Supplementary Table S14C. E-Values for Estimates and CI for Nigeria***

| **Variable** | **Category** | **E-Value for Estimate** | **E-Value for 95% CI** |
| --- | --- | --- | --- |
| Relationship with mother (ref: Very bad/Somewhat bad) | Very good/Somewhat good | 1.53 | 1.00 |
| Relationship with father (ref: Very bad/Somewhat bad) | Very good/Somewhat good | 1.45 | 1.00 |
| Parent marital status (ref: Married) | Divorced | 1.92 | 1.42 |
|  | Single, never married | 1.64 | 1.22 |
|  | One or both parents had died | 2.03 | 1.54 |
| Subjective financial status of family growing up (ref: Got by) | Lived comfortably | 1.34 | 1.00 |
|  | Found it difficult | 1.22 | 1.00 |
|  | Found it very difficult | 1.52 | 1.00 |
| Abuse (ref: No) | Yes | 1.55 | 1.11 |
| Outsider growing up (ref: No) | Yes | 1.30 | 1.00 |
| Self-rated health growing up (ref: Good) | Excellent | 1.21 | 1.00 |
|  | Very good | 1.21 | 1.00 |
|  | Fair | 1.30 | 1.00 |
|  | Poor | 1.44 | 1.00 |
| Immigration status (ref: Born in this country) | Born in another country | 1.31 | 1.00 |
| Age 12 religious service attendance (ref: Never) | At least 1/week | 1.43 | 1.00 |
|  | 1-3/month | 1.41 | 1.00 |
|  | <1/month | 1.99 | 1.31 |
| Birth year (ref: 1998-2005; current age: 18-24) | 1993-1998; age 25-29 | 1.05 | 1.00 |
|  | 1983-1993; age 30-39 | 1.12 | 1.00 |
|  | 1973-1983; age 40-49 | 1.11 | 1.00 |
|  | 1963-1973; age 50-59 | 1.41 | 1.00 |
|  | 1953-1963; age 60-69 | 1.36 | 1.00 |
|  | 1943-1953; age 70-79 | 1.58 | 1.00 |
|  | 1943 or earlier; age 80+ | 1.28 | 1.00 |
| Gender (ref:Male) | Female | 1.29 | 1.00 |
|  | Other | 1.35 | 1.00 |
| Religious affiliation at age 12 (ref: Christianity) | Islam | 1.46 | 1.05 |
|  | Collapsed affiliations with prevalence<3% | 1.89 | 1.00 |
| Race/ethnicity (ref: plurality) | Race/ethnicity minority | 1.24 | 1.00 |

***Supplementary Table S15A. Nationally Representative Childhood Descriptive Statistics for Philippines***

| **Variable** | **Category** | **N (%)** |
| --- | --- | --- |
| Relationship with mother | Very good | 3333 (63%) |
|  | Somewhat good | 1703 (32%) |
|  | Somewhat bad | 124 (2%) |
|  | Very bad | 39 (1%) |
|  | Does not apply | 59 (1%) |
|  | Missing | 35 (1%) |
| Relationships with father | Very good | 3443 (65%) |
|  | Somewhat good | 1429 (27%) |
|  | Somewhat bad | 159 (3%) |
|  | Very bad | 58 (1%) |
|  | Does not apply | 108 (2%) |
|  | Missing | 95 (2%) |
| Parent marital status | Parents married | 4575 (86%) |
|  | Divorced | 64 (1%) |
|  | Single, never married | 517 (10%) |
|  | One or both parents had died | 51 (1%) |
|  | Missing | 85 (2%) |
| Subjective financial status of family growing up | Lived comfortably | 937 (18%) |
|  | Got by | 3006 (57%) |
|  | Found it difficult | 1055 (20%) |
|  | Found it very difficult | 291 (6%) |
|  | Missing | 3 (0%) |
| Abuse | Yes | 420 (8%) |
|  | No | 4837 (91%) |
|  | Missing | 35 (1%) |
| Outsider growing up | Yes | 395 (7%) |
|  | No | 4884 (92%) |
|  | Missing | 13 (0%) |
| Self-rated health growing up | Excellent | 1041 (20%) |
|  | Very good | 559 (11%) |
|  | Good | 2174 (41%) |
|  | Fair | 1246 (24%) |
|  | Poor | 272 (5%) |
|  | Missing | 0 (0%) |
| Immigration status | Born in this country | 5284 (100%) |
|  | Born in another country | 8 (0%) |
| Age 12 religious service attendance | At least 1/week | 2453 (46%) |
|  | 1-3/month | 1699 (32%) |
|  | <1/ month | 892 (17%) |
|  | Never | 201 (4%) |
|  | Missing | 47 (1%) |
| Year of birth | 1998-2005; current age: 18-24 | 1073 (20%) |
|  | 1993-1998; age 25-29 | 695 (13%) |
|  | 1983-1993; age 30-39 | 1160 (22%) |
|  | 1973-1983; age 40-49 | 972 (18%) |
|  | 1963-1973; age 50-59 | 732 (14%) |
|  | 1953-1963; age 60-69 | 495 (9%) |
|  | 1943-1953; age 70-79 | 143 (3%) |
|  | 1943 or earlier; age 80+ | 23 (0%) |
| Gender | Male | 2625 (50%) |
|  | Female | 2643 (50%) |
|  | Other | 13 (0%) |
|  | Missing | 11 (0%) |
| Religious affiliation at age 12 | Christianity | 4968 (94%) |
|  | Islam | 276 (5%) |
|  | Buddhism | 1 (0%) |
|  | Sikhism | 4 (0%) |
|  | Baha’i | 1 (0%) |
|  | Primal, Animist, or Folk religion | 14 (0%) |
|  | Some other religion | 9 (0%) |
|  | No religion/Atheist/Agnostic | 9 (0%) |
|  | Missing | 11 (0%) |
| Race/ethnicity | Tagalog | 1691 (32%) |
|  | Cebuano | 656 (12%) |
|  | Ilocano/Ilokano | 429 (8%) |
|  | Visayan/Bisaya | 739 (14%) |
|  | Ilonggo/Hiligaynon | 428 (8%) |
|  | Bicolano/Bikolano | 300 (6%) |
|  | Waray | 216 (4%) |
|  | Tausug | 94 (2%) |
|  | Maranao | 39 (1%) |
|  | Maguindanaoan | 84 (2%) |
|  | Chinese-Filipino | 3 (0%) |
|  | Kapampangan | 107 (2%) |
|  | Pangasinense | 107 (2%) |
|  | Zamboangueno | 51 (1%) |
|  | Masbateno | 54 (1%) |
|  | Aeta | 1 (0%) |
|  | Igorot | 42 (1%) |
|  | Mangyan | 2 (0%) |
|  | Badjao | 2 (0%) |
|  | Other | 244 (5%) |
|  | Missing | 3 (0%) |

***Supplementary Table S15B. Regression of Optimism on Childhood Predictors for Philippines***

| **Variable** | **Category** | **Estimate** | **SE** | **95% CI** | **Global P-value** |
| --- | --- | --- | --- | --- | --- |
| Relationship with mother (ref: Very bad/Somewhat bad) | Very good/Somewhat good | 0.43 | 0.26 | (-0.08,0.93) | 0.094 |
| Relationship with father (ref: Very bad/Somewhat bad) | Very good/Somewhat good | -0.17 | 0.15 | (-0.47,0.13) | 0.259 |
| Parent marital status (ref: Married) | Divorced | -0.40 | 0.40 | (-1.18,0.39) | 0.060 |
|  | Single, never married | -0.09 | 0.11 | (-0.31,0.12) |  |
|  | One or both parents had died | 0.54 | 0.23 | (0.08,0.99) |  |
| Subjective financial status of family growing up (ref: Got by) | Lived comfortably | 0.17 | 0.08 | (0.02,0.33) | 0.007 |
|  | Found it difficult | -0.19 | 0.10 | (-0.39,0.00) |  |
|  | Found it very difficult | 0.00 | 0.22 | (-0.42,0.43) |  |
| Abuse (ref: No) | Yes | -0.45 | 0.18 | (-0.80,-0.10) | 0.008 |
| Outsider growing up (ref: No) | Yes | -0.17 | 0.15 | (-0.46,0.12) | 0.243 |
| Self-rated health growing up (ref: Good) | Excellent | 0.10 | 0.10 | (-0.09,0.29) | 0.081 |
|  | Very good | 0.00 | 0.11 | (-0.21,0.21) |  |
|  | Fair | -0.11 | 0.09 | (-0.28,0.07) |  |
|  | Poor | -0.37 | 0.19 | (-0.73,-0.01) |  |
| Immigration status (ref: Born in this country) | Born in another country | -0.31 | 0.58 | (-1.46,0.83) | 0.590 |
| Age 12 religious service attendance (ref: Never) | At least 1/week | 0.48 | 0.25 | (0.00,0.97) | 0.015 |
|  | 1-3/month | 0.33 | 0.25 | (-0.17,0.82) |  |
|  | <1/month | 0.29 | 0.27 | (-0.23,0.81) |  |
| Birth year (ref: 1998-2005; current age: 18-24) | 1993-1998; age 25-29 | 0.14 | 0.11 | (-0.07,0.36) | 0.024 |
|  | 1983-1993; age 30-39 | 0.11 | 0.10 | (-0.08,0.31) |  |
|  | 1973-1983; age 40-49 | -0.15 | 0.11 | (-0.36,0.06) |  |
|  | 1963-1973; age 50-59 | -0.15 | 0.14 | (-0.42,0.13) |  |
|  | 1953-1963; age 60-69 | -0.12 | 0.16 | (-0.43,0.19) |  |
|  | 1943-1953; age 70-79 | -0.47 | 0.30 | (-1.07,0.12) |  |
|  | 1943 or earlier; age 80+ | 0.12 | 0.39 | (-0.65,0.90) |  |
| Gender (ref:Male) | Female | 0.14 | 0.07 | (0.01,0.28) | 0.103 |
|  | Other | 0.25 | 0.37 | (-0.48,0.99) |  |
| Religious affiliation at age 12 (ref: Christianity) | Islam | -0.19 | 0.17 | (-0.52,0.14) | 0.374 |
|  | Collapsed affiliations with prevalence<3% | -0.59 | 0.66 | (-1.89,0.70) |  |
| Race/ethnicity (ref: plurality) | Race/ethnicity minority | -0.30 | 0.07 | (-0.44,-0.17) | <.001 |

***Supplementary Table S15C. E-Values for Estimates and CI for Philippines***

| **Variable** | **Category** | **E-Value for Estimate** | **E-Value for 95% CI** |
| --- | --- | --- | --- |
| Relationship with mother (ref: Very bad/Somewhat bad) | Very good/Somewhat good | 1.72 | 1.00 |
| Relationship with father (ref: Very bad/Somewhat bad) | Very good/Somewhat good | 1.37 | 1.00 |
| Parent marital status (ref: Married) | Divorced | 1.68 | 1.00 |
|  | Single, never married | 1.26 | 1.00 |
|  | One or both parents had died | 1.87 | 1.24 |
| Subjective financial status of family growing up (ref: Got by) | Lived comfortably | 1.38 | 1.09 |
|  | Found it difficult | 1.40 | 1.00 |
|  | Found it very difficult | 1.04 | 1.00 |
| Abuse (ref: No) | Yes | 1.75 | 1.27 |
| Outsider growing up (ref: No) | Yes | 1.38 | 1.00 |
| Self-rated health growing up (ref: Good) | Excellent | 1.26 | 1.00 |
|  | Very good | 1.03 | 1.00 |
|  | Fair | 1.28 | 1.00 |
|  | Poor | 1.65 | 1.06 |
| Immigration status (ref: Born in this country) | Born in another country | 1.57 | 1.00 |
| Age 12 religious service attendance (ref: Never) | At least 1/week | 1.80 | 1.03 |
|  | 1-3/month | 1.59 | 1.00 |
|  | <1/month | 1.54 | 1.00 |
| Birth year (ref: 1998-2005; current age: 18-24) | 1993-1998; age 25-29 | 1.34 | 1.00 |
|  | 1983-1993; age 30-39 | 1.29 | 1.00 |
|  | 1973-1983; age 40-49 | 1.34 | 1.00 |
|  | 1963-1973; age 50-59 | 1.34 | 1.00 |
|  | 1953-1963; age 60-69 | 1.29 | 1.00 |
|  | 1943-1953; age 70-79 | 1.79 | 1.00 |
|  | 1943 or earlier; age 80+ | 1.30 | 1.00 |
| Gender (ref:Male) | Female | 1.33 | 1.07 |
|  | Other | 1.49 | 1.00 |
| Religious affiliation at age 12 (ref: Christianity) | Islam | 1.40 | 1.00 |
|  | Collapsed affiliations with prevalence<3% | 1.95 | 1.00 |
| Race/ethnicity (ref: plurality) | Race/ethnicity minority | 1.56 | 1.37 |

***Supplementary Table S16A. Nationally Representative Childhood Descriptive Statistics for Poland***

| **Variable** | **Category** | **N (%)** |
| --- | --- | --- |
| Relationship with mother | Very good | 4879 (47%) |
|  | Somewhat good | 4973 (48%) |
|  | Somewhat bad | 285 (3%) |
|  | Very bad | 58 (1%) |
|  | Does not apply | 80 (1%) |
|  | Missing | 112 (1%) |
| Relationships with father | Very good | 4231 (41%) |
|  | Somewhat good | 4984 (48%) |
|  | Somewhat bad | 516 (5%) |
|  | Very bad | 78 (1%) |
|  | Does not apply | 407 (4%) |
|  | Missing | 173 (2%) |
| Parent marital status | Parents married | 8972 (86%) |
|  | Divorced | 587 (6%) |
|  | Single, never married | 193 (2%) |
|  | One or both parents had died | 313 (3%) |
|  | Missing | 324 (3%) |
| Subjective financial status of family growing up | Lived comfortably | 1384 (13%) |
|  | Got by | 6257 (60%) |
|  | Found it difficult | 2133 (21%) |
|  | Found it very difficult | 509 (5%) |
|  | Missing | 106 (1%) |
| Abuse | Yes | 325 (3%) |
|  | No | 10009 (96%) |
|  | Missing | 55 (1%) |
| Outsider growing up | Yes | 490 (5%) |
|  | No | 9615 (93%) |
|  | Missing | 284 (3%) |
| Self-rated health growing up | Excellent | 2676 (26%) |
|  | Very good | 5371 (52%) |
|  | Good | 1779 (17%) |
|  | Fair | 406 (4%) |
|  | Poor | 123 (1%) |
|  | Missing | 34 (0%) |
| Immigration status | Born in this country | 10258 (99%) |
|  | Born in another country | 108 (1%) |
|  | Missing | 23 (0%) |
| Age 12 religious service attendance | At least 1/week | 4751 (46%) |
|  | 1-3/month | 2689 (26%) |
|  | <1/ month | 2161 (21%) |
|  | Never | 354 (3%) |
|  | Missing | 434 (4%) |
| Year of birth | 1998-2005; current age: 18-24 | 955 (9%) |
|  | 1993-1998; age 25-29 | 761 (7%) |
|  | 1983-1993; age 30-39 | 2159 (21%) |
|  | 1973-1983; age 40-49 | 1956 (19%) |
|  | 1963-1973; age 50-59 | 1670 (16%) |
|  | 1953-1963; age 60-69 | 1909 (18%) |
|  | 1943-1953; age 70-79 | 833 (8%) |
|  | 1943 or earlier; age 80+ | 145 (1%) |
|  | Missing | 1 (0%) |
| Gender | Male | 4974 (48%) |
|  | Female | 5387 (52%) |
|  | Other | 3 (0%) |
|  | Missing | 26 (0%) |
| Religious affiliation at age 12 | Christianity | 9861 (95%) |
|  | Islam | 3 (0%) |
|  | Buddhism | 2 (0%) |
|  | Sikhism | 1 (0%) |
|  | Primal, Animist, or Folk religion | 5 (0%) |
|  | No religion/Atheist/Agnostic | 482 (5%) |
|  | Missing | 35 (0%) |
| Race/ethnicity | Polish | 10309 (99%) |
|  | German | 4 (0%) |
|  | Belarussian | 2 (0%) |
|  | Ukrainian | 38 (0%) |
|  | Silesia | 14 (0%) |
|  | Kashubians | 3 (0%) |
|  | Other | 4 (0%) |
|  | Missing | 14 (0%) |

***Supplementary Table S16B. Regression of Optimism on Childhood Predictors for Poland***

| **Variable** | **Category** | **Estimate** | **SE** | **95% CI** | **Global P-value** |
| --- | --- | --- | --- | --- | --- |
| Relationship with mother (ref: Very bad/Somewhat bad) | Very good/Somewhat good | 0.41 | 0.21 | (0.00,0.83) | 0.045 |
| Relationship with father (ref: Very bad/Somewhat bad) | Very good/Somewhat good | 0.13 | 0.15 | (-0.17,0.43) | 0.393 |
| Parent marital status (ref: Married) | Divorced | -0.52 | 0.11 | (-0.74,-0.31) | <.001 |
|  | Single, never married | -0.69 | 0.28 | (-1.24,-0.14) |  |
|  | One or both parents had died | -0.15 | 0.25 | (-0.64,0.35) |  |
| Subjective financial status of family growing up (ref: Got by) | Lived comfortably | -0.03 | 0.09 | (-0.21,0.16) | 0.863 |
|  | Found it difficult | -0.06 | 0.08 | (-0.23,0.10) |  |
|  | Found it very difficult | 0.06 | 0.21 | (-0.35,0.47) |  |
| Abuse (ref: No) | Yes | -0.30 | 0.24 | (-0.77,0.16) | 0.199 |
| Outsider growing up (ref: No) | Yes | -0.20 | 0.15 | (-0.49,0.10) | 0.164 |
| Self-rated health growing up (ref: Good) | Excellent | 0.35 | 0.12 | (0.12,0.58) | 0.025 |
|  | Very good | 0.14 | 0.09 | (-0.04,0.32) |  |
|  | Fair | -0.06 | 0.21 | (-0.48,0.35) |  |
|  | Poor | 0.35 | 0.36 | (-0.36,1.05) |  |
| Immigration status (ref: Born in this country) | Born in another country | -0.69 | 0.43 | (-1.53,0.15) | 0.107 |
| Age 12 religious service attendance (ref: Never) | At least 1/week | 0.78 | 0.17 | (0.43,1.12) | <.001 |
|  | 1-3/month | 0.47 | 0.18 | (0.12,0.81) |  |
|  | <1/month | 0.13 | 0.18 | (-0.24,0.49) |  |
| Birth year (ref: 1998-2005; current age: 18-24) | 1993-1998; age 25-29 | -0.18 | 0.11 | (-0.40,0.04) | <.001 |
|  | 1983-1993; age 30-39 | -0.23 | 0.11 | (-0.44,-0.01) |  |
|  | 1973-1983; age 40-49 | -0.35 | 0.11 | (-0.57,-0.12) |  |
|  | 1963-1973; age 50-59 | -0.48 | 0.12 | (-0.72,-0.25) |  |
|  | 1953-1963; age 60-69 | -0.47 | 0.13 | (-0.73,-0.21) |  |
|  | 1943-1953; age 70-79 | -0.52 | 0.18 | (-0.87,-0.16) |  |
|  | 1943 or earlier; age 80+ | -0.62 | 0.32 | (-1.25,0.02) |  |
| Gender (ref:Male) | Female | 0.07 | 0.06 | (-0.04,0.18) | 0.149 |
|  | Other | -1.87 | 1.33 | (-4.48,0.75) |  |
| Religious affiliation at age 12 (ref: No religion/Atheist/Agnostic) | Christianity | -0.42 | 0.17 | (-0.75,-0.09) | 0.007 |
|  | Collapsed affiliations with prevalence<3% | -1.38 | 0.59 | (-2.54,-0.22) |  |
| Race/ethnicity (ref: plurality) | Race/ethnicity minority | 0.63 | 0.36 | (-0.07,1.34) | 0.075 |

***Supplementary Table S16C. E-Values for Estimates and CI for Poland***

| **Variable** | **Category** | **E-Value for Estimate** | **E-Value for 95% CI** |
| --- | --- | --- | --- |
| Relationship with mother (ref: Very bad/Somewhat bad) | Very good/Somewhat good | 1.76 | 1.04 |
| Relationship with father (ref: Very bad/Somewhat bad) | Very good/Somewhat good | 1.33 | 1.00 |
| Parent marital status (ref: Married) | Divorced | 1.92 | 1.60 |
|  | Single, never married | 2.17 | 1.35 |
|  | One or both parents had died | 1.36 | 1.00 |
| Subjective financial status of family growing up (ref: Got by) | Lived comfortably | 1.13 | 1.00 |
|  | Found it difficult | 1.21 | 1.00 |
|  | Found it very difficult | 1.20 | 1.00 |
| Abuse (ref: No) | Yes | 1.60 | 1.00 |
| Outsider growing up (ref: No) | Yes | 1.44 | 1.00 |
| Self-rated health growing up (ref: Good) | Excellent | 1.67 | 1.33 |
|  | Very good | 1.35 | 1.00 |
|  | Fair | 1.21 | 1.00 |
|  | Poor | 1.66 | 1.00 |
| Immigration status (ref: Born in this country) | Born in another country | 2.17 | 1.00 |
| Age 12 religious service attendance (ref: Never) | At least 1/week | 2.31 | 1.79 |
|  | 1-3/month | 1.84 | 1.33 |
|  | <1/month | 1.33 | 1.00 |
| Birth year (ref: 1998-2005; current age: 18-24) | 1993-1998; age 25-29 | 1.41 | 1.00 |
|  | 1983-1993; age 30-39 | 1.48 | 1.09 |
|  | 1973-1983; age 40-49 | 1.66 | 1.33 |
|  | 1963-1973; age 50-59 | 1.86 | 1.52 |
|  | 1953-1963; age 60-69 | 1.84 | 1.46 |
|  | 1943-1953; age 70-79 | 1.91 | 1.39 |
|  | 1943 or earlier; age 80+ | 2.06 | 1.00 |
| Gender (ref:Male) | Female | 1.23 | 1.00 |
|  | Other | 4.50 | 1.00 |
| Religious affiliation at age 12 (ref: No religion/Atheist/Agnostic) | Christianity | 1.77 | 1.27 |
|  | Collapsed affiliations with prevalence<3% | 3.38 | 1.48 |
| Race/ethnicity (ref: plurality) | Race/ethnicity minority | 2.09 | 1.00 |

***Supplementary Table S17A. Nationally Representative Childhood Descriptive Statistics for South Africa***

| **Variable** | **Category** | **N (%)** |
| --- | --- | --- |
| Relationship with mother | Very good | 2186 (82%) |
|  | Somewhat good | 263 (10%) |
|  | Somewhat bad | 51 (2%) |
|  | Very bad | 39 (1%) |
|  | Does not apply | 90 (3%) |
|  | Missing | 21 (1%) |
| Relationships with father | Very good | 1656 (62%) |
|  | Somewhat good | 333 (13%) |
|  | Somewhat bad | 86 (3%) |
|  | Very bad | 159 (6%) |
|  | Does not apply | 331 (12%) |
|  | Missing | 85 (3%) |
| Parent marital status | Parents married | 1321 (50%) |
|  | Divorced | 131 (5%) |
|  | Single, never married | 904 (34%) |
|  | One or both parents had died | 140 (5%) |
|  | Missing | 155 (6%) |
| Subjective financial status of family growing up | Lived comfortably | 1050 (40%) |
|  | Got by | 875 (33%) |
|  | Found it difficult | 432 (16%) |
|  | Found it very difficult | 289 (11%) |
|  | Missing | 5 (0%) |
| Abuse | Yes | 450 (17%) |
|  | No | 2149 (81%) |
|  | Missing | 52 (2%) |
| Outsider growing up | Yes | 434 (16%) |
|  | No | 2211 (83%) |
|  | Missing | 6 (0%) |
| Self-rated health growing up | Excellent | 1225 (46%) |
|  | Very good | 590 (22%) |
|  | Good | 370 (14%) |
|  | Fair | 266 (10%) |
|  | Poor | 183 (7%) |
|  | Missing | 17 (1%) |
| Immigration status | Born in this country | 2511 (95%) |
|  | Born in another country | 139 (5%) |
|  | Missing | 1 (0%) |
| Age 12 religious service attendance | At least 1/week | 1681 (63%) |
|  | 1-3/month | 552 (21%) |
|  | <1/ month | 175 (7%) |
|  | Never | 217 (8%) |
|  | Missing | 26 (1%) |
| Year of birth | 1998-2005; current age: 18-24 | 461 (17%) |
|  | 1993-1998; age 25-29 | 364 (14%) |
|  | 1983-1993; age 30-39 | 655 (25%) |
|  | 1973-1983; age 40-49 | 522 (20%) |
|  | 1963-1973; age 50-59 | 309 (12%) |
|  | 1953-1963; age 60-69 | 195 (7%) |
|  | 1943-1953; age 70-79 | 120 (5%) |
|  | 1943 or earlier; age 80+ | 17 (1%) |
|  | Missing | 9 (0%) |
| Gender | Male | 1288 (49%) |
|  | Female | 1356 (51%) |
|  | Other | 2 (0%) |
|  | Missing | 4 (0%) |
| Religious affiliation at age 12 | Christianity | 2323 (88%) |
|  | Islam | 52 (2%) |
|  | Hinduism | 2 (0%) |
|  | Buddhism | 11 (0%) |
|  | Shinto | 2 (0%) |
|  | Taoism | 1 (0%) |
|  | Primal, Animist, or Folk religion | 117 (4%) |
|  | Some other religion | 7 (0%) |
|  | No religion/Atheist/Agnostic | 107 (4%) |
|  | Missing | 27 (1%) |
| Race/ethnicity | Black | 2381 (90%) |
|  | Asian/Indian | 6 (0%) |
|  | Colored | 252 (10%) |
|  | White | 8 (0%) |
|  | Other | 1 (0%) |
|  | Missing | 3 (0%) |

***Supplementary Table S17B. Regression of Optimism on Childhood Predictors for South Africa***

| **Variable** | **Category** | **Estimate** | **SE** | **95% CI** | **Global P-value** |
| --- | --- | --- | --- | --- | --- |
| Relationship with mother (ref: Very bad/Somewhat bad) | Very good/Somewhat good | 0.71 | 0.38 | (-0.04,1.47) | 0.063 |
| Relationship with father (ref: Very bad/Somewhat bad) | Very good/Somewhat good | -0.14 | 0.19 | (-0.51,0.24) | 0.464 |
| Parent marital status (ref: Married) | Divorced | 0.16 | 0.36 | (-0.55,0.88) | 0.032 |
|  | Single, never married | -0.22 | 0.13 | (-0.47,0.04) |  |
|  | One or both parents had died | 0.40 | 0.25 | (-0.10,0.90) |  |
| Subjective financial status of family growing up (ref: Got by) | Lived comfortably | 0.08 | 0.15 | (-0.22,0.38) | 0.785 |
|  | Found it difficult | -0.02 | 0.20 | (-0.42,0.37) |  |
|  | Found it very difficult | 0.16 | 0.21 | (-0.26,0.58) |  |
| Abuse (ref: No) | Yes | -0.31 | 0.17 | (-0.64,0.02) | 0.056 |
| Outsider growing up (ref: No) | Yes | -0.18 | 0.17 | (-0.51,0.14) | 0.271 |
| Self-rated health growing up (ref: Good) | Excellent | 0.12 | 0.17 | (-0.22,0.46) | 0.385 |
|  | Very good | 0.20 | 0.19 | (-0.18,0.57) |  |
|  | Fair | 0.17 | 0.29 | (-0.39,0.73) |  |
|  | Poor | -0.30 | 0.27 | (-0.83,0.23) |  |
| Immigration status (ref: Born in this country) | Born in another country | -0.32 | 0.29 | (-0.89,0.24) | 0.263 |
| Age 12 religious service attendance (ref: Never) | At least 1/week | 0.19 | 0.27 | (-0.35,0.73) | 0.366 |
|  | 1-3/month | 0.35 | 0.28 | (-0.19,0.89) |  |
|  | <1/month | 0.37 | 0.31 | (-0.25,0.99) |  |
| Birth year (ref: 1998-2005; current age: 18-24) | 1993-1998; age 25-29 | -0.18 | 0.17 | (-0.52,0.16) | 0.753 |
|  | 1983-1993; age 30-39 | 0.10 | 0.15 | (-0.20,0.40) |  |
|  | 1973-1983; age 40-49 | -0.09 | 0.19 | (-0.47,0.28) |  |
|  | 1963-1973; age 50-59 | -0.07 | 0.27 | (-0.61,0.48) |  |
|  | 1953-1963; age 60-69 | -0.01 | 0.28 | (-0.56,0.54) |  |
|  | 1943-1953; age 70-79 | 0.17 | 0.40 | (-0.62,0.96) |  |
|  | 1943 or earlier; age 80+ | -0.99 | 1.16 | (-3.29,1.30) |  |
| Gender (ref:Male) | Female | -0.16 | 0.12 | (-0.39,0.07) | 0.003 |
|  | Other | -1.13 | 0.34 | (-1.80,-0.46) |  |
| Religious affiliation at age 12 (ref: No religion/Atheist/Agnostic) | Christianity | -0.62 | 0.32 | (-1.26,0.01) | 0.214 |
|  | Primal, Animist, or Folk religion | -0.38 | 0.35 | (-1.07,0.31) |  |
|  | Collapsed affiliations with prevalence<3% | -0.98 | 0.55 | (-2.06,0.11) |  |
| Race/ethnicity (ref: plurality) | Race/ethnicity minority | -0.11 | 0.25 | (-0.61,0.38) | 0.625 |

***Supplementary Table S17C. E-Values for Estimates and CI for South Africa***

| **Variable** | **Category** | **E-Value for Estimate** | **E-Value for 95% CI** |
| --- | --- | --- | --- |
| Relationship with mother (ref: Very bad/Somewhat bad) | Very good/Somewhat good | 2.01 | 1.00 |
| Relationship with father (ref: Very bad/Somewhat bad) | Very good/Somewhat good | 1.30 | 1.00 |
| Parent marital status (ref: Married) | Divorced | 1.34 | 1.00 |
|  | Single, never married | 1.41 | 1.00 |
|  | One or both parents had died | 1.63 | 1.00 |
| Subjective financial status of family growing up (ref: Got by) | Lived comfortably | 1.22 | 1.00 |
|  | Found it difficult | 1.11 | 1.00 |
|  | Found it very difficult | 1.34 | 1.00 |
| Abuse (ref: No) | Yes | 1.53 | 1.00 |
| Outsider growing up (ref: No) | Yes | 1.37 | 1.00 |
| Self-rated health growing up (ref: Good) | Excellent | 1.27 | 1.00 |
|  | Very good | 1.39 | 1.00 |
|  | Fair | 1.35 | 1.00 |
|  | Poor | 1.51 | 1.00 |
| Immigration status (ref: Born in this country) | Born in another country | 1.54 | 1.00 |
| Age 12 religious service attendance (ref: Never) | At least 1/week | 1.38 | 1.00 |
|  | 1-3/month | 1.58 | 1.00 |
|  | <1/month | 1.60 | 1.00 |
| Birth year (ref: 1998-2005; current age: 18-24) | 1993-1998; age 25-29 | 1.36 | 1.00 |
|  | 1983-1993; age 30-39 | 1.25 | 1.00 |
|  | 1973-1983; age 40-49 | 1.24 | 1.00 |
|  | 1963-1973; age 50-59 | 1.20 | 1.00 |
|  | 1953-1963; age 60-69 | 1.06 | 1.00 |
|  | 1943-1953; age 70-79 | 1.35 | 1.00 |
|  | 1943 or earlier; age 80+ | 2.37 | 1.00 |
| Gender (ref:Male) | Female | 1.33 | 1.00 |
|  | Other | 2.56 | 1.72 |
| Religious affiliation at age 12 (ref: No religion/Atheist/Agnostic) | Christianity | 1.90 | 1.00 |
|  | Primal, Animist, or Folk religion | 1.62 | 1.00 |
|  | Collapsed affiliations with prevalence<3% | 2.35 | 1.00 |
| Race/ethnicity (ref: plurality) | Race/ethnicity minority | 1.27 | 1.00 |

***Supplementary Table S18A. Nationally Representative Childhood Descriptive Statistics for Spain***

| **Variable** | **Category** | **N (%)** |
| --- | --- | --- |
| Relationship with mother | Very good | 4557 (72%) |
|  | Somewhat good | 1258 (20%) |
|  | Somewhat bad | 248 (4%) |
|  | Very bad | 92 (1%) |
|  | Does not apply | 107 (2%) |
|  | Missing | 28 (0%) |
| Relationships with father | Very good | 4131 (66%) |
|  | Somewhat good | 1397 (22%) |
|  | Somewhat bad | 309 (5%) |
|  | Very bad | 178 (3%) |
|  | Does not apply | 243 (4%) |
|  | Missing | 33 (1%) |
| Parent marital status | Parents married | 5285 (84%) |
|  | Divorced | 378 (6%) |
|  | Single, never married | 312 (5%) |
|  | One or both parents had died | 126 (2%) |
|  | Missing | 188 (3%) |
| Subjective financial status of family growing up | Lived comfortably | 2041 (32%) |
|  | Got by | 2956 (47%) |
|  | Found it difficult | 1154 (18%) |
|  | Found it very difficult | 110 (2%) |
|  | Missing | 29 (0%) |
| Abuse | Yes | 659 (10%) |
|  | No | 5510 (88%) |
|  | Missing | 122 (2%) |
| Outsider growing up | Yes | 579 (9%) |
|  | No | 5637 (90%) |
|  | Missing | 75 (1%) |
| Self-rated health growing up | Excellent | 2450 (39%) |
|  | Very good | 2286 (36%) |
|  | Good | 1235 (20%) |
|  | Fair | 164 (3%) |
|  | Poor | 135 (2%) |
|  | Missing | 20 (0%) |
| Immigration status | Born in this country | 5479 (87%) |
|  | Born in another country | 788 (13%) |
|  | Missing | 23 (0%) |
| Age 12 religious service attendance | At least 1/week | 2391 (38%) |
|  | 1-3/month | 1132 (18%) |
|  | <1/ month | 1287 (20%) |
|  | Never | 1445 (23%) |
|  | Missing | 36 (1%) |
| Year of birth | 1998-2005; current age: 18-24 | 594 (9%) |
|  | 1993-1998; age 25-29 | 450 (7%) |
|  | 1983-1993; age 30-39 | 1111 (18%) |
|  | 1973-1983; age 40-49 | 1396 (22%) |
|  | 1963-1973; age 50-59 | 1252 (20%) |
|  | 1953-1963; age 60-69 | 977 (16%) |
|  | 1943-1953; age 70-79 | 467 (7%) |
|  | 1943 or earlier; age 80+ | 43 (1%) |
| Gender | Male | 3142 (50%) |
|  | Female | 3119 (50%) |
|  | Other | 6 (0%) |
|  | Missing | 22 (0%) |
| Religious affiliation at age 12 | Christianity | 5119 (81%) |
|  | Islam | 132 (2%) |
|  | Hinduism | 5 (0%) |
|  | Buddhism | 8 (0%) |
|  | Judaism | 5 (0%) |
|  | Sikhism | 2 (0%) |
|  | Confucianism | 1 (0%) |
|  | Primal, Animist, or Folk religion | 4 (0%) |
|  | Some other religion | 13 (0%) |
|  | No religion/Atheist/Agnostic | 972 (15%) |
|  | Missing | 29 (0%) |

***Supplementary Table S18B. Regression of Optimism on Childhood Predictors for Spain***

| **Variable** | **Category** | **Estimate** | **SE** | **95% CI** | **Global P-value** |
| --- | --- | --- | --- | --- | --- |
| Relationship with mother (ref: Very bad/Somewhat bad) | Very good/Somewhat good | 0.56 | 0.17 | (0.23,0.88) | <.001 |
| Relationship with father (ref: Very bad/Somewhat bad) | Very good/Somewhat good | 0.07 | 0.12 | (-0.17,0.31) | 0.542 |
| Parent marital status (ref: Married) | Divorced | 0.17 | 0.13 | (-0.09,0.43) | 0.232 |
|  | Single, never married | 0.08 | 0.13 | (-0.17,0.33) |  |
|  | One or both parents had died | -0.38 | 0.27 | (-0.91,0.15) |  |
| Subjective financial status of family growing up (ref: Got by) | Lived comfortably | 0.11 | 0.07 | (-0.03,0.25) | 0.304 |
|  | Found it difficult | -0.04 | 0.10 | (-0.24,0.16) |  |
|  | Found it very difficult | -0.26 | 0.35 | (-0.95,0.43) |  |
| Abuse (ref: No) | Yes | -0.14 | 0.11 | (-0.36,0.09) | 0.222 |
| Outsider growing up (ref: No) | Yes | -0.19 | 0.12 | (-0.43,0.05) | 0.113 |
| Self-rated health growing up (ref: Good) | Excellent | 0.63 | 0.10 | (0.43,0.82) | <.001 |
|  | Very good | 0.34 | 0.09 | (0.15,0.52) |  |
|  | Fair | 0.24 | 0.25 | (-0.26,0.74) |  |
|  | Poor | 0.27 | 0.30 | (-0.32,0.87) |  |
| Immigration status (ref: Born in this country) | Born in another country | 0.87 | 0.08 | (0.72,1.03) | <.001 |
| Age 12 religious service attendance (ref: Never) | At least 1/week | 0.30 | 0.09 | (0.12,0.48) | <.001 |
|  | 1-3/month | 0.03 | 0.10 | (-0.16,0.22) |  |
|  | <1/month | -0.08 | 0.10 | (-0.28,0.12) |  |
| Birth year (ref: 1998-2005; current age: 18-24) | 1993-1998; age 25-29 | 0.07 | 0.15 | (-0.23,0.37) | 0.397 |
|  | 1983-1993; age 30-39 | 0.11 | 0.13 | (-0.15,0.37) |  |
|  | 1973-1983; age 40-49 | 0.16 | 0.13 | (-0.10,0.41) |  |
|  | 1963-1973; age 50-59 | 0.20 | 0.13 | (-0.06,0.47) |  |
|  | 1953-1963; age 60-69 | 0.16 | 0.15 | (-0.13,0.46) |  |
|  | 1943-1953; age 70-79 | -0.02 | 0.20 | (-0.41,0.37) |  |
|  | 1943 or earlier; age 80+ | 0.70 | 0.33 | (0.05,1.35) |  |
| Gender (ref:Male) | Female | 0.05 | 0.06 | (-0.08,0.17) | 0.464 |
|  | Other | 0.28 | 0.26 | (-0.22,0.78) |  |
| Religious affiliation at age 12 (ref: No religion/Atheist/Agnostic) | Christianity | 0.12 | 0.10 | (-0.09,0.32) | 0.196 |
|  | Collapsed affiliations with prevalence<3% | -0.24 | 0.26 | (-0.75,0.26) |  |

***Supplementary Table S18C. E-Values for Estimates and CI for Spain***

| **Variable** | **Category** | **E-Value for Estimate** | **E-Value for 95% CI** |
| --- | --- | --- | --- |
| Relationship with mother (ref: Very bad/Somewhat bad) | Very good/Somewhat good | 1.91 | 1.47 |
| Relationship with father (ref: Very bad/Somewhat bad) | Very good/Somewhat good | 1.22 | 1.00 |
| Parent marital status (ref: Married) | Divorced | 1.37 | 1.00 |
|  | Single, never married | 1.24 | 1.00 |
|  | One or both parents had died | 1.67 | 1.00 |
| Subjective financial status of family growing up (ref: Got by) | Lived comfortably | 1.28 | 1.00 |
|  | Found it difficult | 1.15 | 1.00 |
|  | Found it very difficult | 1.51 | 1.00 |
| Abuse (ref: No) | Yes | 1.33 | 1.00 |
| Outsider growing up (ref: No) | Yes | 1.41 | 1.00 |
| Self-rated health growing up (ref: Good) | Excellent | 2.01 | 1.74 |
|  | Very good | 1.61 | 1.35 |
|  | Fair | 1.48 | 1.00 |
|  | Poor | 1.52 | 1.00 |
| Immigration status (ref: Born in this country) | Born in another country | 2.36 | 2.14 |
| Age 12 religious service attendance (ref: Never) | At least 1/week | 1.56 | 1.30 |
|  | 1-3/month | 1.14 | 1.00 |
|  | <1/month | 1.24 | 1.00 |
| Birth year (ref: 1998-2005; current age: 18-24) | 1993-1998; age 25-29 | 1.22 | 1.00 |
|  | 1983-1993; age 30-39 | 1.29 | 1.00 |
|  | 1973-1983; age 40-49 | 1.36 | 1.00 |
|  | 1963-1973; age 50-59 | 1.43 | 1.00 |
|  | 1953-1963; age 60-69 | 1.37 | 1.00 |
|  | 1943-1953; age 70-79 | 1.11 | 1.00 |
|  | 1943 or earlier; age 80+ | 2.11 | 1.17 |
| Gender (ref:Male) | Female | 1.17 | 1.00 |
|  | Other | 1.53 | 1.00 |
| Religious affiliation at age 12 (ref: No religion/Atheist/Agnostic) | Christianity | 1.30 | 1.00 |
|  | Collapsed affiliations with prevalence<3% | 1.48 | 1.00 |

***Supplementary Table S19A. Nationally Representative Childhood Descriptive Statistics for Sweden***

| **Variable** | **Category** | **N (%)** |
| --- | --- | --- |
| Relationship with mother | Very good | 8743 (58%) |
|  | Somewhat good | 4513 (30%) |
|  | Somewhat bad | 1194 (8%) |
|  | Very bad | 371 (2%) |
|  | Does not apply | 216 (1%) |
|  | Missing | 30 (0%) |
| Relationships with father | Very good | 7134 (47%) |
|  | Somewhat good | 4885 (32%) |
|  | Somewhat bad | 1588 (11%) |
|  | Very bad | 725 (5%) |
|  | Does not apply | 720 (5%) |
|  | Missing | 16 (0%) |
| Parent marital status | Parents married | 10887 (72%) |
|  | Divorced | 1927 (13%) |
|  | Single, never married | 1747 (12%) |
|  | One or both parents had died | 362 (2%) |
|  | Missing | 145 (1%) |
| Subjective financial status of family growing up | Lived comfortably | 5951 (39%) |
|  | Got by | 7717 (51%) |
|  | Found it difficult | 1238 (8%) |
|  | Found it very difficult | 140 (1%) |
|  | Missing | 22 (0%) |
| Abuse | Yes | 2288 (15%) |
|  | No | 12735 (85%) |
|  | Missing | 45 (0%) |
| Outsider growing up | Yes | 1867 (12%) |
|  | No | 13034 (86%) |
|  | Missing | 168 (1%) |
| Self-rated health growing up | Excellent | 5733 (38%) |
|  | Very good | 5124 (34%) |
|  | Good | 2669 (18%) |
|  | Fair | 1108 (7%) |
|  | Poor | 397 (3%) |
|  | Missing | 38 (0%) |
| Immigration status | Born in this country | 13922 (92%) |
|  | Born in another country | 1052 (7%) |
|  | Missing | 94 (1%) |
| Age 12 religious service attendance | At least 1/week | 955 (6%) |
|  | 1-3/month | 1362 (9%) |
|  | <1/ month | 6224 (41%) |
|  | Never | 6472 (43%) |
|  | Missing | 54 (0%) |
| Year of birth | 1998-2005; current age: 18-24 | 1515 (10%) |
|  | 1993-1998; age 25-29 | 1399 (9%) |
|  | 1983-1993; age 30-39 | 2398 (16%) |
|  | 1973-1983; age 40-49 | 2221 (15%) |
|  | 1963-1973; age 50-59 | 2493 (17%) |
|  | 1953-1963; age 60-69 | 2168 (14%) |
|  | 1943-1953; age 70-79 | 2253 (15%) |
|  | 1943 or earlier; age 80+ | 621 (4%) |
| Gender | Male | 7536 (50%) |
|  | Female | 7493 (50%) |
|  | Other | 27 (0%) |
|  | Missing | 12 (0%) |
| Religious affiliation at age 12 | Christianity | 10617 (70%) |
|  | Islam | 462 (3%) |
|  | Hinduism | 16 (0%) |
|  | Buddhism | 41 (0%) |
|  | Judaism | 51 (0%) |
|  | Sikhism | 9 (0%) |
|  | Baha’i | 3 (0%) |
|  | Shinto | 1 (0%) |
|  | Confucianism | 4 (0%) |
|  | Primal, Animist, or Folk religion | 31 (0%) |
|  | Some other religion | 69 (0%) |
|  | No religion/Atheist/Agnostic | 3738 (25%) |
|  | Missing | 26 (0%) |

***Supplementary Table S19B. Regression of Optimism on Childhood Predictors for Sweden***

| **Variable** | **Category** | **Estimate** | **SE** | **95% CI** | **Global P-value** |
| --- | --- | --- | --- | --- | --- |
| Relationship with mother (ref: Very bad/Somewhat bad) | Very good/Somewhat good | -0.04 | 0.08 | (-0.20,0.13) | 0.674 |
| Relationship with father (ref: Very bad/Somewhat bad) | Very good/Somewhat good | 0.13 | 0.07 | (-0.01,0.27) | 0.077 |
| Parent marital status (ref: Married) | Divorced | 0.21 | 0.08 | (0.06,0.36) | 0.002 |
|  | Single, never married | -0.06 | 0.08 | (-0.22,0.10) |  |
|  | One or both parents had died | 0.37 | 0.15 | (0.08,0.65) |  |
| Subjective financial status of family growing up (ref: Got by) | Lived comfortably | 0.31 | 0.05 | (0.22,0.40) | <.001 |
|  | Found it difficult | -0.12 | 0.09 | (-0.31,0.06) |  |
|  | Found it very difficult | 0.18 | 0.28 | (-0.36,0.72) |  |
| Abuse (ref: No) | Yes | -0.13 | 0.07 | (-0.27,0.01) | 0.071 |
| Outsider growing up (ref: No) | Yes | -0.50 | 0.09 | (-0.67,-0.32) | <.001 |
| Self-rated health growing up (ref: Good) | Excellent | 1.18 | 0.07 | (1.05,1.31) | <.001 |
|  | Very good | 0.58 | 0.07 | (0.45,0.71) |  |
|  | Fair | -0.15 | 0.11 | (-0.37,0.06) |  |
|  | Poor | -0.46 | 0.20 | (-0.84,-0.07) |  |
| Immigration status (ref: Born in this country) | Born in another country | 0.32 | 0.09 | (0.15,0.50) | <.001 |
| Age 12 religious service attendance (ref: Never) | At least 1/week | 0.20 | 0.11 | (-0.01,0.41) | 0.082 |
|  | 1-3/month | 0.01 | 0.08 | (-0.15,0.17) |  |
|  | <1/month | 0.10 | 0.05 | (0.00,0.19) |  |
| Birth year (ref: 1998-2005; current age: 18-24) | 1993-1998; age 25-29 | 0.07 | 0.10 | (-0.13,0.28) | <.001 |
|  | 1983-1993; age 30-39 | 0.05 | 0.09 | (-0.13,0.23) |  |
|  | 1973-1983; age 40-49 | 0.11 | 0.10 | (-0.08,0.31) |  |
|  | 1963-1973; age 50-59 | 0.33 | 0.09 | (0.14,0.51) |  |
|  | 1953-1963; age 60-69 | 0.35 | 0.09 | (0.16,0.53) |  |
|  | 1943-1953; age 70-79 | 0.39 | 0.10 | (0.20,0.58) |  |
|  | 1943 or earlier; age 80+ | 0.18 | 0.14 | (-0.09,0.46) |  |
| Gender (ref:Male) | Female | 0.14 | 0.04 | (0.06,0.23) | <.001 |
|  | Other | -1.21 | 0.40 | (-1.99,-0.43) |  |
| Religious affiliation at age 12 (ref: No religion/Atheist/Agnostic) | Christianity | 0.25 | 0.06 | (0.13,0.36) | <.001 |
|  | Islam | 0.50 | 0.16 | (0.19,0.81) |  |
|  | Collapsed affiliations with prevalence<3% | 0.35 | 0.23 | (-0.09,0.79) |  |

***Supplementary Table S19C. E-Values for Estimates and CI for Sweden***

| **Variable** | **Category** | **E-Value for Estimate** | **E-Value for 95% CI** |
| --- | --- | --- | --- |
| Relationship with mother (ref: Very bad/Somewhat bad) | Very good/Somewhat good | 1.13 | 1.00 |
| Relationship with father (ref: Very bad/Somewhat bad) | Very good/Somewhat good | 1.28 | 1.00 |
| Parent marital status (ref: Married) | Divorced | 1.39 | 1.19 |
|  | Single, never married | 1.17 | 1.00 |
|  | One or both parents had died | 1.58 | 1.21 |
| Subjective financial status of family growing up (ref: Got by) | Lived comfortably | 1.51 | 1.40 |
|  | Found it difficult | 1.28 | 1.00 |
|  | Found it very difficult | 1.35 | 1.00 |
| Abuse (ref: No) | Yes | 1.29 | 1.00 |
| Outsider growing up (ref: No) | Yes | 1.73 | 1.53 |
| Self-rated health growing up (ref: Good) | Excellent | 2.55 | 2.38 |
|  | Very good | 1.83 | 1.68 |
|  | Fair | 1.32 | 1.00 |
|  | Poor | 1.68 | 1.20 |
| Immigration status (ref: Born in this country) | Born in another country | 1.53 | 1.31 |
| Age 12 religious service attendance (ref: Never) | At least 1/week | 1.38 | 1.00 |
|  | 1-3/month | 1.06 | 1.00 |
|  | <1/month | 1.24 | 1.04 |
| Birth year (ref: 1998-2005; current age: 18-24) | 1993-1998; age 25-29 | 1.20 | 1.00 |
|  | 1983-1993; age 30-39 | 1.16 | 1.00 |
|  | 1973-1983; age 40-49 | 1.27 | 1.00 |
|  | 1963-1973; age 50-59 | 1.53 | 1.31 |
|  | 1953-1963; age 60-69 | 1.56 | 1.33 |
|  | 1943-1953; age 70-79 | 1.60 | 1.38 |
|  | 1943 or earlier; age 80+ | 1.36 | 1.00 |
| Gender (ref:Male) | Female | 1.31 | 1.17 |
|  | Other | 2.59 | 1.65 |
| Religious affiliation at age 12 (ref: No religion/Atheist/Agnostic) | Christianity | 1.43 | 1.29 |
|  | Islam | 1.73 | 1.37 |
|  | Collapsed affiliations with prevalence<3% | 1.56 | 1.00 |

***Supplementary Table S20A. Nationally Representative Childhood Descriptive Statistics for Tanzania***

| **Variable** | **Category** | **N (%)** |
| --- | --- | --- |
| Relationship with mother | Very good | 7739 (85%) |
|  | Somewhat good | 796 (9%) |
|  | Somewhat bad | 84 (1%) |
|  | Very bad | 84 (1%) |
|  | Does not apply | 303 (3%) |
|  | Missing | 70 (1%) |
| Relationships with father | Very good | 6831 (75%) |
|  | Somewhat good | 1101 (12%) |
|  | Somewhat bad | 203 (2%) |
|  | Very bad | 247 (3%) |
|  | Does not apply | 550 (6%) |
|  | Missing | 142 (2%) |
| Parent marital status | Parents married | 6929 (76%) |
|  | Divorced | 678 (7%) |
|  | Single, never married | 751 (8%) |
|  | One or both parents had died | 313 (3%) |
|  | Missing | 404 (4%) |
| Subjective financial status of family growing up | Lived comfortably | 2611 (29%) |
|  | Got by | 2909 (32%) |
|  | Found it difficult | 2679 (30%) |
|  | Found it very difficult | 814 (9%) |
|  | Missing | 61 (1%) |
| Abuse | Yes | 716 (8%) |
|  | No | 8328 (92%) |
|  | Missing | 32 (0%) |
| Outsider growing up | Yes | 734 (8%) |
|  | No | 8320 (92%) |
|  | Missing | 22 (0%) |
| Self-rated health growing up | Excellent | 2406 (27%) |
|  | Very good | 2036 (22%) |
|  | Good | 2946 (32%) |
|  | Fair | 1177 (13%) |
|  | Poor | 456 (5%) |
|  | Missing | 54 (1%) |
| Immigration status | Born in this country | 9048 (100%) |
|  | Born in another country | 25 (0%) |
|  | Missing | 1 (0%) |
| Age 12 religious service attendance | At least 1/week | 5580 (61%) |
|  | 1-3/month | 2383 (26%) |
|  | <1/ month | 333 (4%) |
|  | Never | 595 (7%) |
|  | Missing | 184 (2%) |
| Year of birth | 1998-2005; current age: 18-24 | 2284 (25%) |
|  | 1993-1998; age 25-29 | 1349 (15%) |
|  | 1983-1993; age 30-39 | 2060 (23%) |
|  | 1973-1983; age 40-49 | 1503 (17%) |
|  | 1963-1973; age 50-59 | 912 (10%) |
|  | 1953-1963; age 60-69 | 575 (6%) |
|  | 1943-1953; age 70-79 | 297 (3%) |
|  | 1943 or earlier; age 80+ | 93 (1%) |
|  | Missing | 2 (0%) |
| Gender | Male | 4299 (47%) |
|  | Female | 4776 (53%) |
| Religious affiliation at age 12 | Christianity | 5651 (62%) |
|  | Islam | 3060 (34%) |
|  | Baha’i | 1 (0%) |
|  | Primal, Animist, or Folk religion | 11 (0%) |
|  | No religion/Atheist/Agnostic | 345 (4%) |
|  | Missing | 7 (0%) |
| Race/ethnicity | African | 9060 (100%) |
|  | Indian | 3 (0%) |
|  | Arab | 11 (0%) |
|  | Missing | 2 (0%) |

***Supplementary Table S20B. Regression of Optimism on Childhood Predictors for Tanzania***

| **Variable** | **Category** | **Estimate** | **SE** | **95% CI** | **Global P-value** |
| --- | --- | --- | --- | --- | --- |
| Relationship with mother (ref: Very bad/Somewhat bad) | Very good/Somewhat good | 0.33 | 0.21 | (-0.08,0.75) | 0.115 |
| Relationship with father (ref: Very bad/Somewhat bad) | Very good/Somewhat good | -0.23 | 0.13 | (-0.49,0.04) | 0.095 |
| Parent marital status (ref: Married) | Divorced | -0.40 | 0.12 | (-0.64,-0.15) | 0.007 |
|  | Single, never married | 0.01 | 0.13 | (-0.24,0.27) |  |
|  | One or both parents had died | -0.26 | 0.20 | (-0.65,0.13) |  |
| Subjective financial status of family growing up (ref: Got by) | Lived comfortably | -0.04 | 0.08 | (-0.20,0.13) | 0.455 |
|  | Found it difficult | -0.06 | 0.07 | (-0.21,0.08) |  |
|  | Found it very difficult | -0.18 | 0.12 | (-0.42,0.05) |  |
| Abuse (ref: No) | Yes | -0.11 | 0.12 | (-0.34,0.13) | 0.366 |
| Outsider growing up (ref: No) | Yes | -0.13 | 0.12 | (-0.37,0.10) | 0.261 |
| Self-rated health growing up (ref: Good) | Excellent | 0.04 | 0.08 | (-0.13,0.21) | 0.378 |
|  | Very good | 0.08 | 0.09 | (-0.09,0.26) |  |
|  | Fair | -0.08 | 0.11 | (-0.29,0.13) |  |
|  | Poor | -0.15 | 0.15 | (-0.44,0.14) |  |
| Immigration status (ref: Born in this country) | Born in another country | 0.14 | 0.53 | (-0.90,1.19) | 0.787 |
| Age 12 religious service attendance (ref: Never) | At least 1/week | 0.27 | 0.19 | (-0.10,0.64) | 0.052 |
|  | 1-3/month | 0.04 | 0.20 | (-0.36,0.44) |  |
|  | <1/month | 0.18 | 0.25 | (-0.32,0.67) |  |
| Birth year (ref: 1998-2005; current age: 18-24) | 1993-1998; age 25-29 | 0.02 | 0.09 | (-0.15,0.20) | 0.002 |
|  | 1983-1993; age 30-39 | -0.06 | 0.09 | (-0.23,0.11) |  |
|  | 1973-1983; age 40-49 | -0.31 | 0.10 | (-0.51,-0.10) |  |
|  | 1963-1973; age 50-59 | -0.31 | 0.12 | (-0.55,-0.08) |  |
|  | 1953-1963; age 60-69 | -0.25 | 0.15 | (-0.55,0.05) |  |
|  | 1943-1953; age 70-79 | -0.44 | 0.30 | (-1.03,0.15) |  |
|  | 1943 or earlier; age 80+ | -1.18 | 0.50 | (-2.15,-0.20) |  |
| Gender (ref:Male) | Female | -0.04 | 0.06 | (-0.16,0.07) | 0.459 |
| Religious affiliation at age 12 (ref: No religion/Atheist/Agnostic) | Christianity | 0.11 | 0.24 | (-0.37,0.58) | 0.229 |
|  | Islam | 0.22 | 0.25 | (-0.28,0.72) |  |
|  | Collapsed affiliations with prevalence<3% | 0.80 | 0.53 | (-0.25,1.85) |  |
| Race/ethnicity (ref: plurality) | Race/ethnicity minority | -0.99 | 0.47 | (-1.94,-0.05) | 0.012 |

***Supplementary Table S20C. E-Values for Estimates and CI for Tanzania***

| **Variable** | **Category** | **E-Value for Estimate** | **E-Value for 95% CI** |
| --- | --- | --- | --- |
| Relationship with mother (ref: Very bad/Somewhat bad) | Very good/Somewhat good | 1.53 | 1.00 |
| Relationship with father (ref: Very bad/Somewhat bad) | Very good/Somewhat good | 1.41 | 1.00 |
| Parent marital status (ref: Married) | Divorced | 1.61 | 1.32 |
|  | Single, never married | 1.08 | 1.00 |
|  | One or both parents had died | 1.44 | 1.00 |
| Subjective financial status of family growing up (ref: Got by) | Lived comfortably | 1.14 | 1.00 |
|  | Found it difficult | 1.19 | 1.00 |
|  | Found it very difficult | 1.36 | 1.00 |
| Abuse (ref: No) | Yes | 1.25 | 1.00 |
| Outsider growing up (ref: No) | Yes | 1.29 | 1.00 |
| Self-rated health growing up (ref: Good) | Excellent | 1.14 | 1.00 |
|  | Very good | 1.22 | 1.00 |
|  | Fair | 1.22 | 1.00 |
|  | Poor | 1.32 | 1.00 |
| Immigration status (ref: Born in this country) | Born in another country | 1.30 | 1.00 |
| Age 12 religious service attendance (ref: Never) | At least 1/week | 1.46 | 1.00 |
|  | 1-3/month | 1.14 | 1.00 |
|  | <1/month | 1.34 | 1.00 |
| Birth year (ref: 1998-2005; current age: 18-24) | 1993-1998; age 25-29 | 1.10 | 1.00 |
|  | 1983-1993; age 30-39 | 1.18 | 1.00 |
|  | 1973-1983; age 40-49 | 1.50 | 1.25 |
|  | 1963-1973; age 50-59 | 1.51 | 1.21 |
|  | 1953-1963; age 60-69 | 1.44 | 1.00 |
|  | 1943-1953; age 70-79 | 1.65 | 1.00 |
|  | 1943 or earlier; age 80+ | 2.53 | 1.38 |
| Gender (ref:Male) | Female | 1.15 | 1.00 |
| Religious affiliation at age 12 (ref: No religion/Atheist/Agnostic) | Christianity | 1.25 | 1.00 |
|  | Islam | 1.40 | 1.00 |
|  | Collapsed affiliations with prevalence<3% | 2.06 | 1.00 |
| Race/ethnicity (ref: plurality) | Race/ethnicity minority | 2.30 | 1.21 |

***Supplementary Table S21A. Nationally Representative Childhood Descriptive Statistics for Türkiye***

| **Variable** | **Category** | **N (%)** |
| --- | --- | --- |
| Relationship with mother | Very good | 970 (66%) |
|  | Somewhat good | 401 (27%) |
|  | Somewhat bad | 48 (3%) |
|  | Very bad | 26 (2%) |
|  | Does not apply | 21 (1%) |
|  | Missing | 7 (0%) |
| Relationships with father | Very good | 795 (54%) |
|  | Somewhat good | 425 (29%) |
|  | Somewhat bad | 73 (5%) |
|  | Very bad | 95 (6%) |
|  | Does not apply | 60 (4%) |
|  | Missing | 25 (2%) |
| Parent marital status | Parents married | 1325 (90%) |
|  | Divorced | 57 (4%) |
|  | Single, never married | 7 (0%) |
|  | One or both parents had died | 61 (4%) |
|  | Missing | 23 (2%) |
| Subjective financial status of family growing up | Lived comfortably | 498 (34%) |
|  | Got by | 647 (44%) |
|  | Found it difficult | 218 (15%) |
|  | Found it very difficult | 108 (7%) |
|  | Missing | 2 (0%) |
| Abuse | Yes | 158 (11%) |
|  | No | 1290 (88%) |
|  | Missing | 25 (2%) |
| Outsider growing up | Yes | 157 (11%) |
|  | No | 1306 (89%) |
|  | Missing | 9 (1%) |
| Self-rated health growing up | Excellent | 377 (26%) |
|  | Very good | 410 (28%) |
|  | Good | 419 (28%) |
|  | Fair | 220 (15%) |
|  | Poor | 47 (3%) |
|  | Missing | 0 (0%) |
| Immigration status | Born in this country | 1415 (96%) |
|  | Born in another country | 58 (4%) |
| Age 12 religious service attendance | At least 1/week | 609 (41%) |
|  | 1-3/month | 238 (16%) |
|  | <1/ month | 225 (15%) |
|  | Never | 383 (26%) |
|  | Missing | 18 (1%) |
| Year of birth | 1998-2005; current age: 18-24 | 222 (15%) |
|  | 1993-1998; age 25-29 | 152 (10%) |
|  | 1983-1993; age 30-39 | 315 (21%) |
|  | 1973-1983; age 40-49 | 312 (21%) |
|  | 1963-1973; age 50-59 | 225 (15%) |
|  | 1953-1963; age 60-69 | 164 (11%) |
|  | 1943-1953; age 70-79 | 65 (4%) |
|  | 1943 or earlier; age 80+ | 18 (1%) |
| Gender | Male | 754 (51%) |
|  | Female | 719 (49%) |
| Religious affiliation at age 12 | Christianity | 1 (0%) |
|  | Islam | 1439 (98%) |
|  | Judaism | 1 (0%) |
|  | No religion/Atheist/Agnostic | 13 (1%) |
|  | Missing | 19 (1%) |
| Race/ethnicity | Turkish | 1030 (70%) |
|  | Kurdish/Zaza | 252 (17%) |
|  | Arab | 51 (3%) |
|  | Laz | 25 (2%) |
|  | Circassian | 19 (1%) |
|  | Bosnian | 5 (0%) |
|  | Armenian | 1 (0%) |
|  | Georgian | 4 (0%) |
|  | Uyghur | 1 (0%) |
|  | Albanian | 8 (1%) |
|  | Greek | 1 (0%) |
|  | Azeri | 9 (1%) |
|  | Other | 58 (4%) |
|  | Missing | 9 (1%) |

***Supplementary Table S21B. Regression of Optimism on Childhood Predictors for Türkiye***

| **Variable** | **Category** | **Estimate** | **SE** | **95% CI** | **Global P-value** |
| --- | --- | --- | --- | --- | --- |
| Relationship with mother (ref: Very bad/Somewhat bad) | Very good/Somewhat good | 0.23 | 0.39 | (-0.53,0.99) | 0.545 |
| Relationship with father (ref: Very bad/Somewhat bad) | Very good/Somewhat good | -0.03 | 0.25 | (-0.52,0.47) | 0.858 |
| Parent marital status (ref: Married) | Divorced | 0.19 | 0.40 | (-0.59,0.97) | 0.339 |
|  | Single, never married | -0.43 | 1.11 | (-2.63,1.77) |  |
|  | One or both parents had died | -0.67 | 0.41 | (-1.47,0.13) |  |
| Subjective financial status of family growing up (ref: Got by) | Lived comfortably | 0.10 | 0.17 | (-0.23,0.43) | 0.292 |
|  | Found it difficult | -0.43 | 0.28 | (-0.97,0.12) |  |
|  | Found it very difficult | 0.19 | 0.34 | (-0.47,0.85) |  |
| Abuse (ref: No) | Yes | -0.33 | 0.29 | (-0.90,0.24) | 0.243 |
| Outsider growing up (ref: No) | Yes | -0.35 | 0.28 | (-0.89,0.19) | 0.202 |
| Self-rated health growing up (ref: Good) | Excellent | 0.15 | 0.19 | (-0.23,0.53) | 0.408 |
|  | Very good | -0.01 | 0.20 | (-0.41,0.39) |  |
|  | Fair | 0.05 | 0.25 | (-0.45,0.55) |  |
|  | Poor | -0.96 | 0.57 | (-2.07,0.15) |  |
| Immigration status (ref: Born in this country) | Born in another country | -0.01 | 0.44 | (-0.86,0.85) | 0.980 |
| Age 12 religious service attendance (ref: Never) | At least 1/week | 0.36 | 0.21 | (-0.05,0.76) | 0.057 |
|  | 1-3/month | 0.22 | 0.25 | (-0.27,0.71) |  |
|  | <1/month | -0.18 | 0.25 | (-0.67,0.31) |  |
| Birth year (ref: 1998-2005; current age: 18-24) | 1993-1998; age 25-29 | 0.60 | 0.29 | (0.02,1.17) | <.001 |
|  | 1983-1993; age 30-39 | 0.33 | 0.25 | (-0.15,0.81) |  |
|  | 1973-1983; age 40-49 | 0.54 | 0.24 | (0.08,1.01) |  |
|  | 1963-1973; age 50-59 | 0.67 | 0.28 | (0.13,1.21) |  |
|  | 1953-1963; age 60-69 | 0.97 | 0.32 | (0.34,1.60) |  |
|  | 1943-1953; age 70-79 | 0.70 | 0.59 | (-0.45,1.85) |  |
|  | 1943 or earlier; age 80+ | 2.02 | 0.32 | (1.38,2.65) |  |
| Gender (ref:Male) | Female | 0.71 | 0.16 | (0.40,1.03) | <.001 |
| Religious affiliation at age 12 (ref: Islam) | Collapsed affiliations with prevalence<3% | 0.85 | 0.43 | (0.00,1.69) | 0.022 |
| Race/ethnicity (ref: plurality) | Race/ethnicity minority | -0.10 | 0.19 | (-0.47,0.27) | 0.596 |

***Supplementary Table S21C. E-Values for Estimates and CI for Türkiye***

| **Variable** | **Category** | **E-Value for Estimate** | **E-Value for 95% CI** |
| --- | --- | --- | --- |
| Relationship with mother (ref: Very bad/Somewhat bad) | Very good/Somewhat good | 1.40 | 1.00 |
| Relationship with father (ref: Very bad/Somewhat bad) | Very good/Somewhat good | 1.12 | 1.00 |
| Parent marital status (ref: Married) | Divorced | 1.35 | 1.00 |
|  | Single, never married | 1.62 | 1.00 |
|  | One or both parents had died | 1.87 | 1.00 |
| Subjective financial status of family growing up (ref: Got by) | Lived comfortably | 1.24 | 1.00 |
|  | Found it difficult | 1.61 | 1.00 |
|  | Found it very difficult | 1.35 | 1.00 |
| Abuse (ref: No) | Yes | 1.51 | 1.00 |
| Outsider growing up (ref: No) | Yes | 1.53 | 1.00 |
| Self-rated health growing up (ref: Good) | Excellent | 1.30 | 1.00 |
|  | Very good | 1.05 | 1.00 |
|  | Fair | 1.15 | 1.00 |
|  | Poor | 2.19 | 1.00 |
| Immigration status (ref: Born in this country) | Born in another country | 1.05 | 1.00 |
| Age 12 religious service attendance (ref: Never) | At least 1/week | 1.54 | 1.00 |
|  | 1-3/month | 1.38 | 1.00 |
|  | <1/month | 1.34 | 1.00 |
| Birth year (ref: 1998-2005; current age: 18-24) | 1993-1998; age 25-29 | 1.79 | 1.10 |
|  | 1983-1993; age 30-39 | 1.51 | 1.00 |
|  | 1973-1983; age 40-49 | 1.74 | 1.21 |
|  | 1963-1973; age 50-59 | 1.87 | 1.27 |
|  | 1953-1963; age 60-69 | 2.20 | 1.52 |
|  | 1943-1953; age 70-79 | 1.91 | 1.00 |
|  | 1943 or earlier; age 80+ | 3.60 | 2.70 |
| Gender (ref:Male) | Female | 1.92 | 1.58 |
| Religious affiliation at age 12 (ref: Islam) | Collapsed affiliations with prevalence<3% | 2.06 | 1.08 |
| Race/ethnicity (ref: plurality) | Race/ethnicity minority | 1.23 | 1.00 |

***Supplementary Table S22A. Nationally Representative Childhood Descriptive Statistics for United Kingdom***

| **Variable** | **Category** | **N (%)** |
| --- | --- | --- |
| Relationship with mother | Very good | 3435 (64%) |
|  | Somewhat good | 1338 (25%) |
|  | Somewhat bad | 325 (6%) |
|  | Very bad | 150 (3%) |
|  | Does not apply | 92 (2%) |
|  | Missing | 27 (1%) |
| Relationships with father | Very good | 2907 (54%) |
|  | Somewhat good | 1383 (26%) |
|  | Somewhat bad | 407 (8%) |
|  | Very bad | 321 (6%) |
|  | Does not apply | 321 (6%) |
|  | Missing | 29 (1%) |
| Parent marital status | Parents married | 4343 (81%) |
|  | Divorced | 481 (9%) |
|  | Single, never married | 315 (6%) |
|  | One or both parents had died | 154 (3%) |
|  | Missing | 75 (1%) |
| Subjective financial status of family growing up | Lived comfortably | 2552 (48%) |
|  | Got by | 1933 (36%) |
|  | Found it difficult | 632 (12%) |
|  | Found it very difficult | 230 (4%) |
|  | Missing | 22 (0%) |
| Abuse | Yes | 864 (16%) |
|  | No | 4455 (83%) |
|  | Missing | 49 (1%) |
| Outsider growing up | Yes | 1017 (19%) |
|  | No | 4308 (80%) |
|  | Missing | 43 (1%) |
| Self-rated health growing up | Excellent | 2154 (40%) |
|  | Very good | 1736 (32%) |
|  | Good | 995 (19%) |
|  | Fair | 332 (6%) |
|  | Poor | 130 (2%) |
|  | Missing | 20 (0%) |
| Immigration status | Born in this country | 4659 (87%) |
|  | Born in another country | 682 (13%) |
|  | Missing | 27 (0%) |
| Age 12 religious service attendance | At least 1/week | 1732 (32%) |
|  | 1-3/month | 733 (14%) |
|  | <1/ month | 903 (17%) |
|  | Never | 1972 (37%) |
|  | Missing | 28 (1%) |
| Year of birth | 1998-2005; current age: 18-24 | 490 (9%) |
|  | 1993-1998; age 25-29 | 391 (7%) |
|  | 1983-1993; age 30-39 | 946 (18%) |
|  | 1973-1983; age 40-49 | 827 (15%) |
|  | 1963-1973; age 50-59 | 949 (18%) |
|  | 1953-1963; age 60-69 | 889 (17%) |
|  | 1943-1953; age 70-79 | 711 (13%) |
|  | 1943 or earlier; age 80+ | 163 (3%) |
|  | Missing | 1 (0%) |
| Gender | Male | 2557 (48%) |
|  | Female | 2789 (52%) |
|  | Other | 14 (0%) |
|  | Missing | 9 (0%) |
| Religious affiliation at age 12 | Christianity | 3461 (64%) |
|  | Islam | 230 (4%) |
|  | Hinduism | 88 (2%) |
|  | Buddhism | 15 (0%) |
|  | Judaism | 59 (1%) |
|  | Sikhism | 30 (1%) |
|  | Baha’i | 5 (0%) |
|  | Jainism | 0 (0%) |
|  | Taoism | 2 (0%) |
|  | Confucianism | 3 (0%) |
|  | Primal, Animist, or Folk religion | 22 (0%) |
|  | Some other religion | 24 (0%) |
|  | No religion/Atheist/Agnostic | 1409 (26%) |
|  | Missing | 21 (0%) |
| Race/ethnicity | Asian | 426 (8%) |
|  | Black | 152 (3%) |
|  | White | 4647 (87%) |
|  | Other | 96 (2%) |
|  | Missing | 47 (1%) |

***Supplementary Table S22B. Regression of Optimism on Childhood Predictors for United Kingdom***

| **Variable** | **Category** | **Estimate** | **SE** | **95% CI** | **Global P-value** |
| --- | --- | --- | --- | --- | --- |
| Relationship with mother (ref: Very bad/Somewhat bad) | Very good/Somewhat good | 0.40 | 0.18 | (0.05,0.76) | 0.026 |
| Relationship with father (ref: Very bad/Somewhat bad) | Very good/Somewhat good | 0.26 | 0.15 | (-0.03,0.56) | 0.078 |
| Parent marital status (ref: Married) | Divorced | -0.02 | 0.17 | (-0.36,0.31) | 0.449 |
|  | Single, never married | -0.36 | 0.23 | (-0.81,0.09) |  |
|  | One or both parents had died | -0.19 | 0.36 | (-0.91,0.52) |  |
| Subjective financial status of family growing up (ref: Got by) | Lived comfortably | 0.11 | 0.10 | (-0.08,0.30) | 0.192 |
|  | Found it difficult | -0.04 | 0.15 | (-0.34,0.26) |  |
|  | Found it very difficult | -0.46 | 0.29 | (-1.03,0.11) |  |
| Abuse (ref: No) | Yes | -0.66 | 0.14 | (-0.93,-0.38) | <.001 |
| Outsider growing up (ref: No) | Yes | -0.28 | 0.13 | (-0.54,-0.02) | 0.030 |
| Self-rated health growing up (ref: Good) | Excellent | 0.72 | 0.13 | (0.47,0.97) | <.001 |
|  | Very good | 0.61 | 0.13 | (0.36,0.86) |  |
|  | Fair | -0.42 | 0.22 | (-0.86,0.02) |  |
|  | Poor | -0.77 | 0.37 | (-1.49,-0.05) |  |
| Immigration status (ref: Born in this country) | Born in another country | 0.40 | 0.14 | (0.12,0.67) | 0.004 |
| Age 12 religious service attendance (ref: Never) | At least 1/week | 0.48 | 0.12 | (0.24,0.72) | <.001 |
|  | 1-3/month | 0.57 | 0.14 | (0.29,0.85) |  |
|  | <1/month | 0.24 | 0.14 | (-0.03,0.51) |  |
| Birth year (ref: 1998-2005; current age: 18-24) | 1993-1998; age 25-29 | 0.44 | 0.24 | (-0.03,0.91) | 0.316 |
|  | 1983-1993; age 30-39 | 0.17 | 0.20 | (-0.23,0.57) |  |
|  | 1973-1983; age 40-49 | 0.09 | 0.21 | (-0.32,0.49) |  |
|  | 1963-1973; age 50-59 | 0.27 | 0.20 | (-0.13,0.67) |  |
|  | 1953-1963; age 60-69 | 0.25 | 0.21 | (-0.17,0.67) |  |
|  | 1943-1953; age 70-79 | 0.43 | 0.22 | (-0.00,0.86) |  |
|  | 1943 or earlier; age 80+ | 0.43 | 0.33 | (-0.21,1.07) |  |
| Gender (ref:Male) | Female | -0.07 | 0.09 | (-0.24,0.10) | 0.089 |
|  | Other | -1.00 | 0.50 | (-1.98,-0.03) |  |
| Religious affiliation at age 12 (ref: No religion/Atheist/Agnostic) | Christianity | 0.41 | 0.12 | (0.17,0.65) | 0.005 |
|  | Islam | 0.42 | 0.25 | (-0.06,0.91) |  |
|  | Collapsed affiliations with prevalence<3% | 0.12 | 0.22 | (-0.32,0.56) |  |
| Race/ethnicity (ref: plurality) | Race/ethnicity minority | 0.53 | 0.16 | (0.21,0.84) | <.001 |

***Supplementary Table S22C. E-Values for Estimates and CI for United Kingdom***

| **Variable** | **Category** | **E-Value for Estimate** | **E-Value for 95% CI** |
| --- | --- | --- | --- |
| Relationship with mother (ref: Very bad/Somewhat bad) | Very good/Somewhat good | 1.59 | 1.15 |
| Relationship with father (ref: Very bad/Somewhat bad) | Very good/Somewhat good | 1.44 | 1.00 |
| Parent marital status (ref: Married) | Divorced | 1.11 | 1.00 |
|  | Single, never married | 1.55 | 1.00 |
|  | One or both parents had died | 1.36 | 1.00 |
| Subjective financial status of family growing up (ref: Got by) | Lived comfortably | 1.25 | 1.00 |
|  | Found it difficult | 1.14 | 1.00 |
|  | Found it very difficult | 1.66 | 1.00 |
| Abuse (ref: No) | Yes | 1.87 | 1.57 |
| Outsider growing up (ref: No) | Yes | 1.46 | 1.11 |
| Self-rated health growing up (ref: Good) | Excellent | 1.94 | 1.67 |
|  | Very good | 1.82 | 1.55 |
|  | Fair | 1.61 | 1.00 |
|  | Poor | 1.99 | 1.16 |
| Immigration status (ref: Born in this country) | Born in another country | 1.59 | 1.27 |
| Age 12 religious service attendance (ref: Never) | At least 1/week | 1.68 | 1.41 |
|  | 1-3/month | 1.78 | 1.48 |
|  | <1/month | 1.41 | 1.00 |
| Birth year (ref: 1998-2005; current age: 18-24) | 1993-1998; age 25-29 | 1.64 | 1.00 |
|  | 1983-1993; age 30-39 | 1.33 | 1.00 |
|  | 1973-1983; age 40-49 | 1.22 | 1.00 |
|  | 1963-1973; age 50-59 | 1.45 | 1.00 |
|  | 1953-1963; age 60-69 | 1.43 | 1.00 |
|  | 1943-1953; age 70-79 | 1.62 | 1.00 |
|  | 1943 or earlier; age 80+ | 1.63 | 1.00 |
| Gender (ref:Male) | Female | 1.19 | 1.00 |
|  | Other | 2.26 | 1.12 |
| Religious affiliation at age 12 (ref: No religion/Atheist/Agnostic) | Christianity | 1.61 | 1.34 |
|  | Islam | 1.62 | 1.00 |
|  | Collapsed affiliations with prevalence<3% | 1.26 | 1.00 |
| Race/ethnicity (ref: plurality) | Race/ethnicity minority | 1.73 | 1.38 |

***Supplementary Table S23A. Nationally Representative Childhood Descriptive Statistics for United States***

| **Variable** | **Category** | **N (%)** |
| --- | --- | --- |
| Relationship with mother | Very good | 20590 (54%) |
|  | Somewhat good | 11525 (30%) |
|  | Somewhat bad | 3523 (9%) |
|  | Very bad | 1874 (5%) |
|  | Does not apply | 694 (2%) |
|  | Missing | 106 (0%) |
| Relationships with father | Very good | 15313 (40%) |
|  | Somewhat good | 12665 (33%) |
|  | Somewhat bad | 4879 (13%) |
|  | Very bad | 2604 (7%) |
|  | Does not apply | 2811 (7%) |
|  | Missing | 38 (0%) |
| Parent marital status | Parents married | 27415 (72%) |
|  | Divorced | 6325 (17%) |
|  | Single, never married | 3048 (8%) |
|  | One or both parents had died | 1024 (3%) |
|  | Missing | 500 (1%) |
| Subjective financial status of family growing up | Lived comfortably | 15116 (39%) |
|  | Got by | 15682 (41%) |
|  | Found it difficult | 5152 (13%) |
|  | Found it very difficult | 2342 (6%) |
|  | Missing | 19 (0%) |
| Abuse | Yes | 10026 (26%) |
|  | No | 28045 (73%) |
|  | Missing | 242 (1%) |
| Outsider growing up | Yes | 10185 (27%) |
|  | No | 27714 (72%) |
|  | Missing | 413 (1%) |
| Self-rated health growing up | Excellent | 16866 (44%) |
|  | Very good | 12108 (32%) |
|  | Good | 6444 (17%) |
|  | Fair | 2303 (6%) |
|  | Poor | 520 (1%) |
|  | Missing | 71 (0%) |
| Immigration status | Born in this country | 34865 (91%) |
|  | Born in another country | 3020 (8%) |
|  | Missing | 427 (1%) |
| Age 12 religious service attendance | At least 1/week | 18609 (49%) |
|  | 1-3/month | 6644 (17%) |
|  | <1/ month | 5829 (15%) |
|  | Never | 7085 (18%) |
|  | Missing | 145 (0%) |
| Year of birth | 1998-2005; current age: 18-24 | 2682 (7%) |
|  | 1993-1998; age 25-29 | 3540 (9%) |
|  | 1983-1993; age 30-39 | 7284 (19%) |
|  | 1973-1983; age 40-49 | 5649 (15%) |
|  | 1963-1973; age 50-59 | 6745 (18%) |
|  | 1953-1963; age 60-69 | 6832 (18%) |
|  | 1943-1953; age 70-79 | 4054 (11%) |
|  | 1943 or earlier; age 80+ | 1525 (4%) |
| Gender | Male | 18222 (48%) |
|  | Female | 19562 (51%) |
|  | Other | 392 (1%) |
|  | Missing | 136 (0%) |
| Religious affiliation at age 12 | Christianity | 30444 (79%) |
|  | Islam | 220 (1%) |
|  | Hinduism | 203 (1%) |
|  | Buddhism | 172 (0%) |
|  | Judaism | 787 (2%) |
|  | Sikhism | 47 (0%) |
|  | Baha’i | 4 (0%) |
|  | Jainism | 18 (0%) |
|  | Shinto | 6 (0%) |
|  | Taoism | 17 (0%) |
|  | Confucianism | 8 (0%) |
|  | Primal, Animist, or Folk religion | 67 (0%) |
|  | Some other religion | 359 (1%) |
|  | No religion/Atheist/Agnostic | 5845 (15%) |
|  | Missing | 115 (0%) |
| Race/ethnicity | White | 23605 (62%) |
|  | Other | 997 (3%) |
|  | Black | 4501 (12%) |
|  | Asian | 2466 (6%) |
|  | Hispanic | 6724 (18%) |
|  | Missing | 20 (0%) |

***Supplementary Table S23B. Regression of Optimism on Childhood Predictors for United States***

| **Variable** | **Category** | **Estimate** | **SE** | **95% CI** | **Global P-value** |
| --- | --- | --- | --- | --- | --- |
| Relationship with mother (ref: Very bad/Somewhat bad) | Very good/Somewhat good | 0.31 | 0.10 | (0.11,0.50) | 0.003 |
| Relationship with father (ref: Very bad/Somewhat bad) | Very good/Somewhat good | 0.25 | 0.08 | (0.09,0.42) | 0.003 |
| Parent marital status (ref: Married) | Divorced | 0.04 | 0.09 | (-0.13,0.21) | 0.301 |
|  | Single, never married | 0.14 | 0.18 | (-0.21,0.49) |  |
|  | One or both parents had died | -0.33 | 0.22 | (-0.77,0.11) |  |
| Subjective financial status of family growing up (ref: Got by) | Lived comfortably | 0.22 | 0.06 | (0.11,0.32) | <.001 |
|  | Found it difficult | 0.03 | 0.09 | (-0.15,0.21) |  |
|  | Found it very difficult | 0.02 | 0.16 | (-0.29,0.32) |  |
| Abuse (ref: No) | Yes | -0.23 | 0.08 | (-0.38,-0.08) | 0.003 |
| Outsider growing up (ref: No) | Yes | -0.61 | 0.08 | (-0.78,-0.45) | <.001 |
| Self-rated health growing up (ref: Good) | Excellent | 0.91 | 0.09 | (0.74,1.08) | <.001 |
|  | Very good | 0.35 | 0.09 | (0.17,0.53) |  |
|  | Fair | -0.01 | 0.20 | (-0.41,0.38) |  |
|  | Poor | -0.38 | 0.45 | (-1.26,0.51) |  |
| Immigration status (ref: Born in this country) | Born in another country | 0.16 | 0.12 | (-0.08,0.40) | 0.180 |
| Age 12 religious service attendance (ref: Never) | At least 1/week | 0.18 | 0.11 | (-0.03,0.40) | 0.162 |
|  | 1-3/month | 0.17 | 0.11 | (-0.05,0.39) |  |
|  | <1/month | 0.03 | 0.11 | (-0.19,0.24) |  |
| Birth year (ref: 1998-2005; current age: 18-24) | 1993-1998; age 25-29 | -0.18 | 0.22 | (-0.62,0.25) | <.001 |
|  | 1983-1993; age 30-39 | 0.21 | 0.19 | (-0.17,0.59) |  |
|  | 1973-1983; age 40-49 | 0.33 | 0.19 | (-0.04,0.70) |  |
|  | 1963-1973; age 50-59 | 0.58 | 0.18 | (0.23,0.94) |  |
|  | 1953-1963; age 60-69 | 0.77 | 0.18 | (0.42,1.12) |  |
|  | 1943-1953; age 70-79 | 0.87 | 0.18 | (0.52,1.23) |  |
|  | 1943 or earlier; age 80+ | 0.86 | 0.21 | (0.45,1.28) |  |
| Gender (ref:Male) | Female | 0.36 | 0.06 | (0.25,0.47) | <.001 |
|  | Other | -0.72 | 0.44 | (-1.59,0.14) |  |
| Religious affiliation at age 12 (ref: No religion/Atheist/Agnostic) | Christianity | 0.46 | 0.12 | (0.22,0.71) | <.001 |
|  | Collapsed affiliations with prevalence<3% | 0.34 | 0.14 | (0.06,0.62) |  |
| Race/ethnicity (ref: plurality) | Race/ethnicity minority | 0.43 | 0.06 | (0.31,0.55) | <.001 |

***Supplementary Table S23C. E-Values for Estimates and CI for United States***

| **Variable** | **Category** | **E-Value for Estimate** | **E-Value for 95% CI** |
| --- | --- | --- | --- |
| Relationship with mother (ref: Very bad/Somewhat bad) | Very good/Somewhat good | 1.51 | 1.26 |
| Relationship with father (ref: Very bad/Somewhat bad) | Very good/Somewhat good | 1.45 | 1.23 |
| Parent marital status (ref: Married) | Divorced | 1.14 | 1.00 |
|  | Single, never married | 1.30 | 1.00 |
|  | One or both parents had died | 1.54 | 1.00 |
| Subjective financial status of family growing up (ref: Got by) | Lived comfortably | 1.40 | 1.26 |
|  | Found it difficult | 1.13 | 1.00 |
|  | Found it very difficult | 1.09 | 1.00 |
| Abuse (ref: No) | Yes | 1.42 | 1.22 |
| Outsider growing up (ref: No) | Yes | 1.87 | 1.68 |
| Self-rated health growing up (ref: Good) | Excellent | 2.23 | 2.02 |
|  | Very good | 1.56 | 1.34 |
|  | Fair | 1.08 | 1.00 |
|  | Poor | 1.60 | 1.00 |
| Immigration status (ref: Born in this country) | Born in another country | 1.33 | 1.00 |
| Age 12 religious service attendance (ref: Never) | At least 1/week | 1.36 | 1.00 |
|  | 1-3/month | 1.34 | 1.00 |
|  | <1/month | 1.12 | 1.00 |
| Birth year (ref: 1998-2005; current age: 18-24) | 1993-1998; age 25-29 | 1.36 | 1.00 |
|  | 1983-1993; age 30-39 | 1.39 | 1.00 |
|  | 1973-1983; age 40-49 | 1.54 | 1.00 |
|  | 1963-1973; age 50-59 | 1.83 | 1.42 |
|  | 1953-1963; age 60-69 | 2.06 | 1.65 |
|  | 1943-1953; age 70-79 | 2.18 | 1.76 |
|  | 1943 or earlier; age 80+ | 2.17 | 1.69 |
| Gender (ref:Male) | Female | 1.57 | 1.44 |
|  | Other | 2.00 | 1.00 |
| Religious affiliation at age 12 (ref: No religion/Atheist/Agnostic) | Christianity | 1.70 | 1.41 |
|  | Collapsed affiliations with prevalence<3% | 1.55 | 1.18 |
| Race/ethnicity (ref: plurality) | Race/ethnicity minority | 1.65 | 1.51 |

***Supplementary Table S24. Population Weighted Meta-Analysis of Regression Results.***

| Variable | Category | Est | 95% CI | SE |
| --- | --- | --- | --- | --- |
| Relationship with mother | (Ref: Very bad/somewhat bad) |  |  |  |
|  | Very good/somewhat good | 0.03 | (-0.18,0.25) | 0.109 |
| Relationship with father | (Ref: Very bad/somewhat bad) |  |  |  |
|  | Very good/somewhat good | 0.02 | (-0.16,0.21) | 0.094 |
| Parent marital status | (Ref: Parents married) |  |  |  |
|  | Divorced | -0.24 | (-0.40,-0.08) | 0.080 |
|  | Single, never married | -0.09 | (-0.22,0.04) | 0.064 |
|  | One or both parents had died | 0.10 | (-0.05,0.24) | 0.072 |
| Subjective financial status of family growing up | (Ref: Got by) |  |  |  |
|  | Lived comfortably | 0.02 | (-0.05,0.08) | 0.033 |
|  | Found it difficult | -0.12 | (-0.19,-0.04) | 0.040 |
|  | Found it very difficult | -0.12 | (-0.23,-0.02) | 0.053 |
| Abuse | (Ref: No) |  |  |  |
|  | Yes | -0.24 | (-0.33,-0.16) | 0.044 |
| Outsider growing up | (Ref: No) |  |  |  |
|  | Yes | -0.25 | (-0.33,-0.16) | 0.043 |
| Self-rated health growing up | (Ref: Good) |  |  |  |
|  | Excellent | 0.28 | (0.18,0.37) | 0.048 |
|  | Very good | 0.10 | (0.03,0.17) | 0.036 |
|  | Fair | -0.16 | (-0.25,-0.07) | 0.048 |
|  | Poor | -0.15 | (-0.33,0.04) | 0.095 |
| Immigration status | (Ref: Born in this country) |  |  |  |
|  | Born in another country | -0.12 | (-0.41,0.16) | 0.146 |
| Age 12 religious service attendance | (Ref: Never) |  |  |  |
|  | At least 1/week | 0.17 | (0.08,0.26) | 0.047 |
|  | 1-3/month | 0.11 | (0.01,0.22) | 0.053 |
|  | <1/month | 0.07 | (-0.03,0.18) | 0.053 |
| Year of birth | (Ref: 1998-2005; age 18-24) |  |  |  |
|  | 1993-1998; age 25-29 | -0.01 | (-0.11,0.10) | 0.053 |
|  | 1983-1993; age 30-39 | 0.02 | (-0.07,0.12) | 0.047 |
|  | 1973-1983; age 40-49 | 0.05 | (-0.05,0.14) | 0.049 |
|  | 1963-1973; age 50-59 | 0.17 | (0.06,0.28) | 0.056 |
|  | 1953-1963; age 60-69 | -0.01 | (-0.14,0.12) | 0.067 |
|  | 1943-1953; age 70-79 | 0.01 | (-0.18,0.20) | 0.096 |
|  | 1943 or earlier; age 80+ | -0.14 | (-0.58,0.31) | 0.227 |
| Gender | (Ref: Male) |  |  |  |
|  | Female | 0.20 | (0.15,0.25) | 0.027 |
|  | Other | -0.67 | (-1.10,-0.23) | 0.222 |

***Supplementary Table S25. Population Weighted Meta-Analysis of E-Values.***

| Variable | Category | E-value of the  Point Estimate | E-value of the Confidence Limit |
| --- | --- | --- | --- |
| Relationship with mother | (Ref: Very bad/somewhat bad) |  |  |
|  | Very good/somewhat good | 1.14 | 1.00 |
| Relationship with father | (Ref: Very bad/somewhat bad) |  |  |
|  | Very good/somewhat good | 1.11 | 1.00 |
| Parent marital status | (Ref: Parents married) |  |  |
|  | Divorced | 1.45 | 1.23 |
|  | Single, never married | 1.24 | 1.00 |
|  | One or both parents had died | 1.25 | 1.00 |
| Subjective financial status of family growing up | (Ref: Got by) |  |  |
|  | Lived comfortably | 1.09 | 1.00 |
|  | Found it difficult | 1.28 | 1.14 |
|  | Found it very difficult | 1.30 | 1.10 |
| Abuse | (Ref: No) |  |  |
|  | Yes | 1.46 | 1.35 |
| Outsider growing up | (Ref: No) |  |  |
|  | Yes | 1.47 | 1.35 |
| Self-rated health growing up | (Ref: Good) |  |  |
|  | Excellent | 1.50 | 1.38 |
|  | Very good | 1.26 | 1.13 |
|  | Fair | 1.35 | 1.20 |
|  | Poor | 1.33 | 1.00 |
| Immigration status | (Ref: Born in this country) |  |  |
|  | Born in another country | 1.29 | 1.00 |
| Age 12 religious service attendance | (Ref: Never) |  |  |
|  | At least 1/week | 1.37 | 1.23 |
|  | 1-3/month | 1.28 | 1.07 |
|  | <1/month | 1.21 | 1.00 |
| Year of birth | (Ref: 1998-2005; age 18-24) |  |  |
|  | 1993-1998; age 25-29 | 1.06 | 1.00 |
|  | 1983-1993; age 30-39 | 1.11 | 1.00 |
|  | 1973-1983; age 40-49 | 1.16 | 1.00 |
|  | 1963-1973; age 50-59 | 1.36 | 1.19 |
|  | 1953-1963; age 60-69 | 1.08 | 1.00 |
|  | 1943-1953; age 70-79 | 1.07 | 1.00 |
|  | 1943 or earlier; age 80+ | 1.31 | 1.00 |
| Gender | (Ref: Male) |  |  |
|  | Female | 1.40 | 1.33 |
|  | Other | 2.00 | 1.45 |

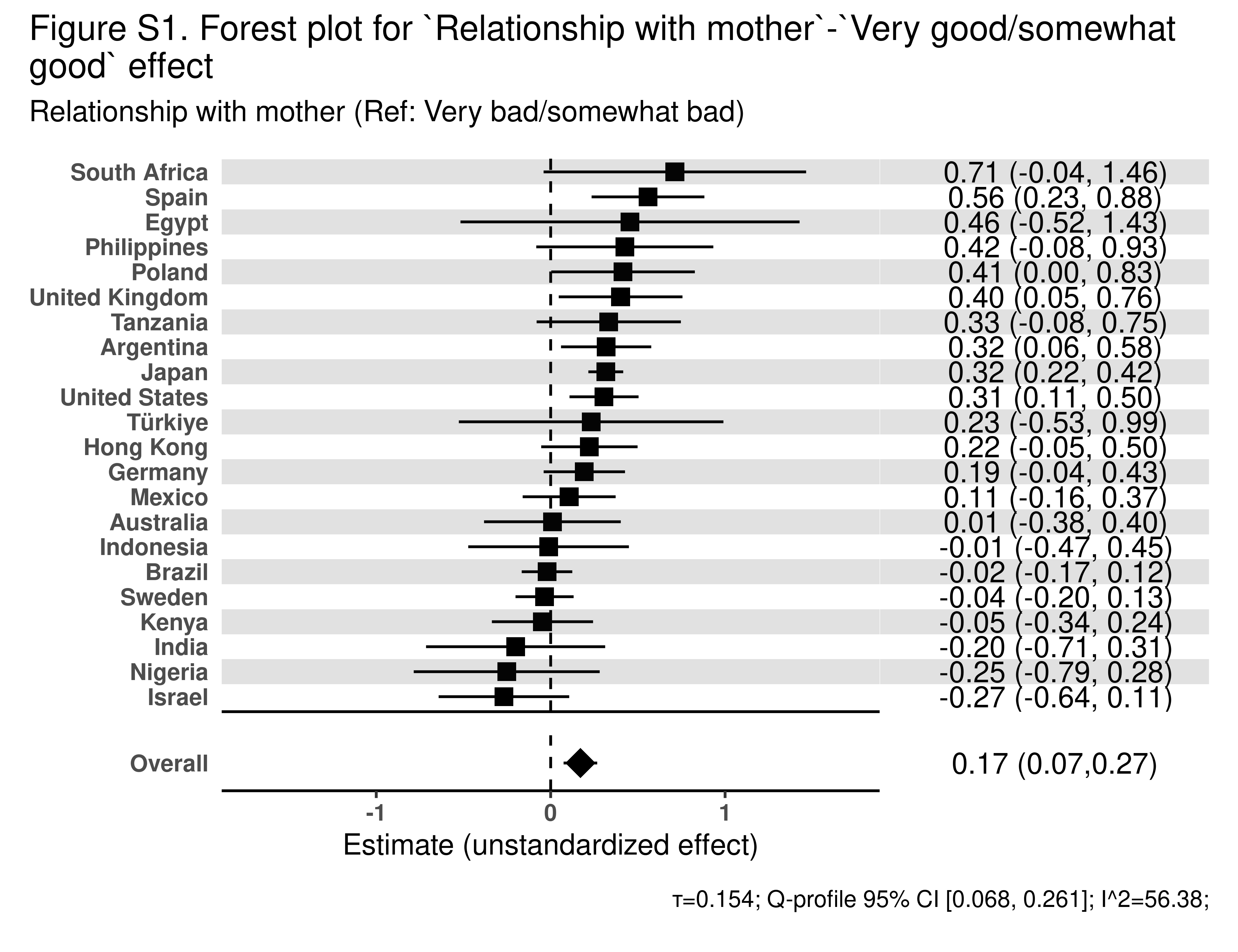

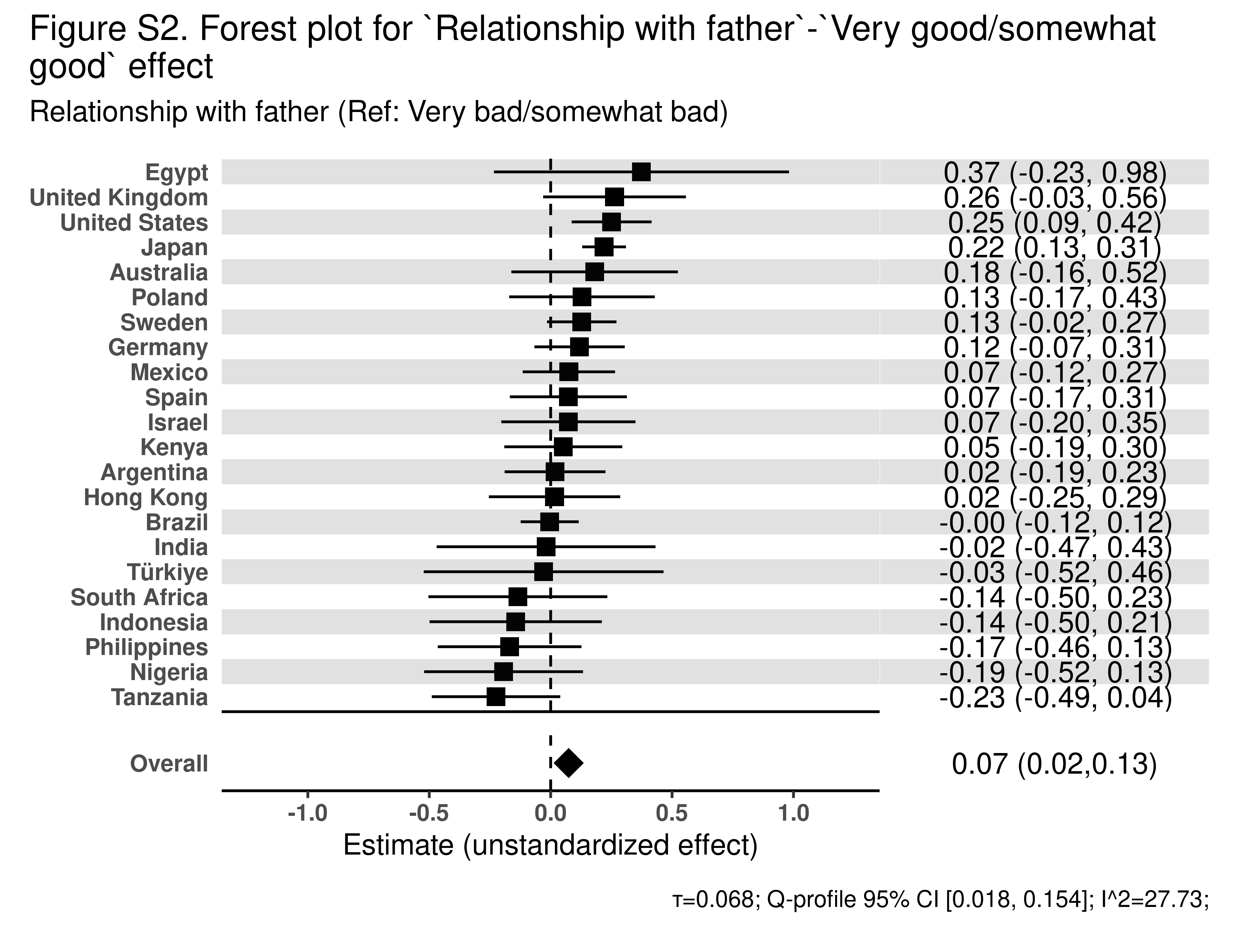

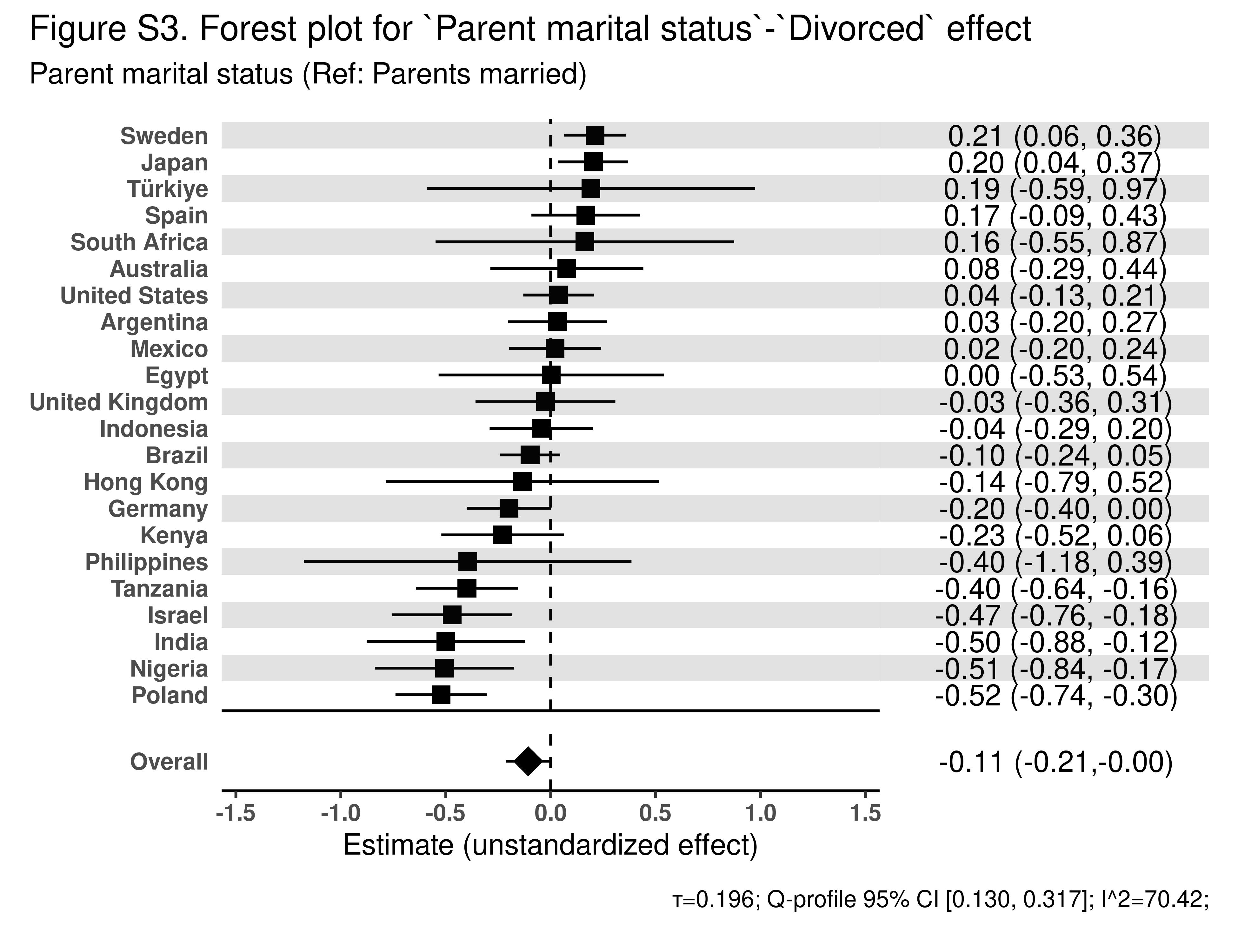

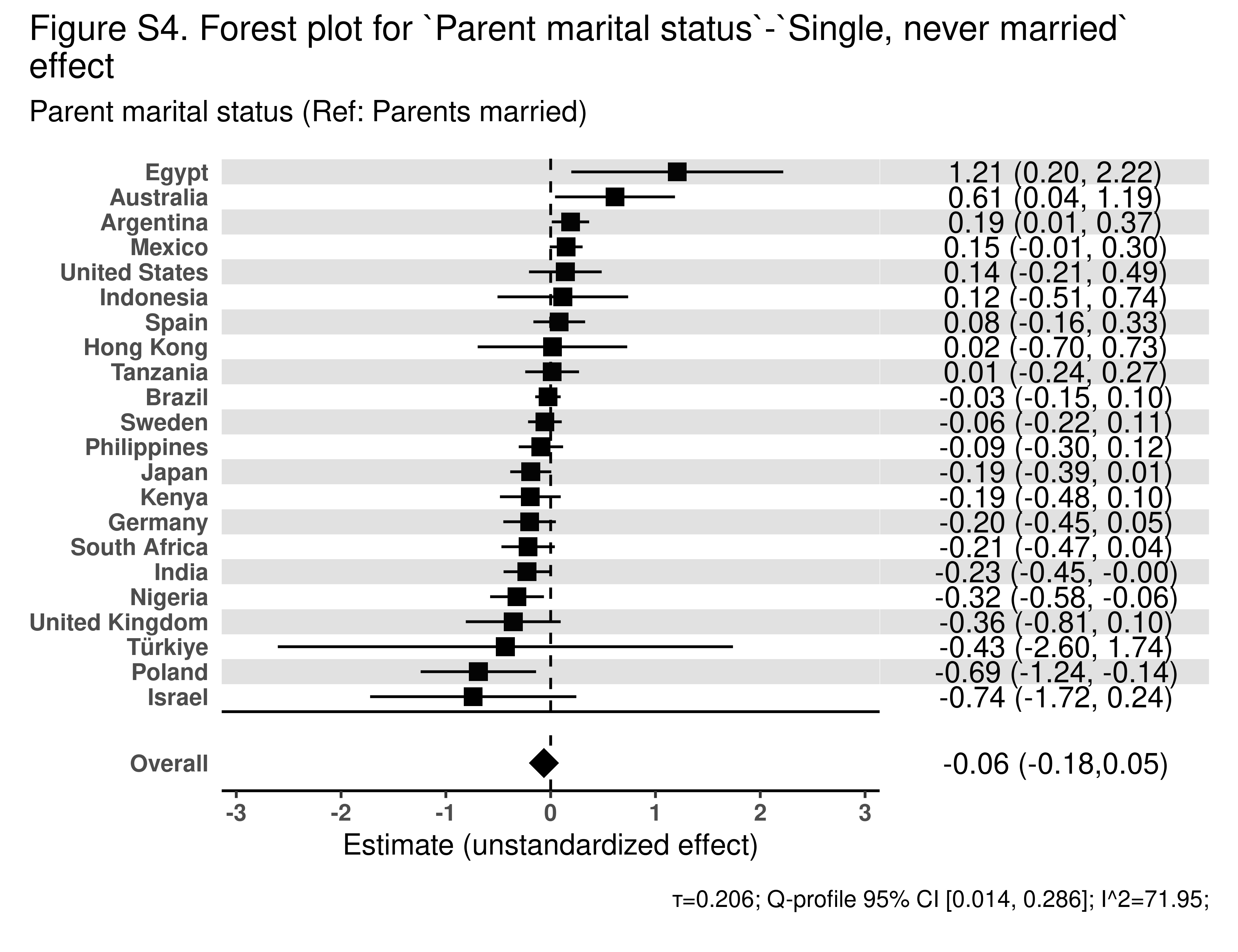

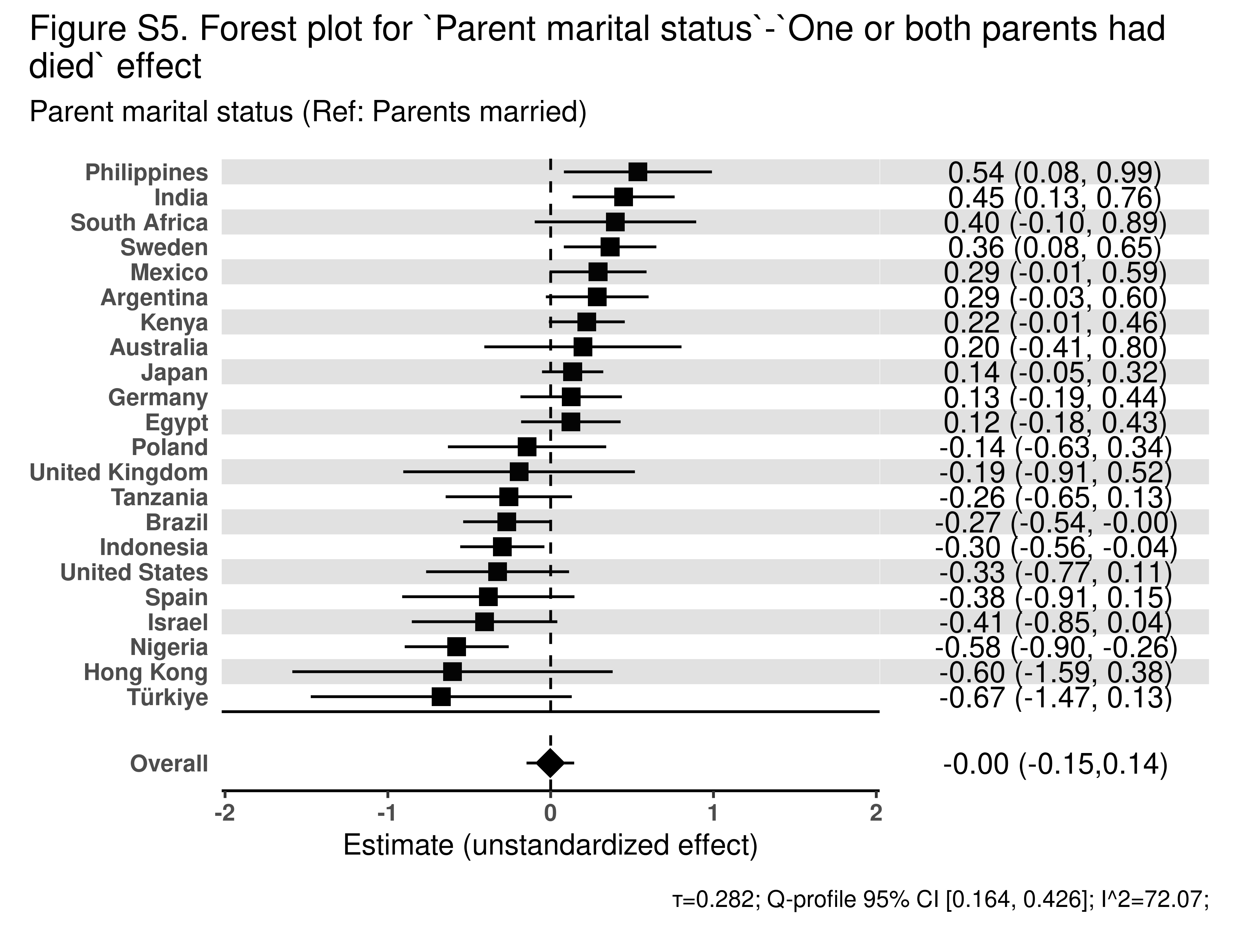

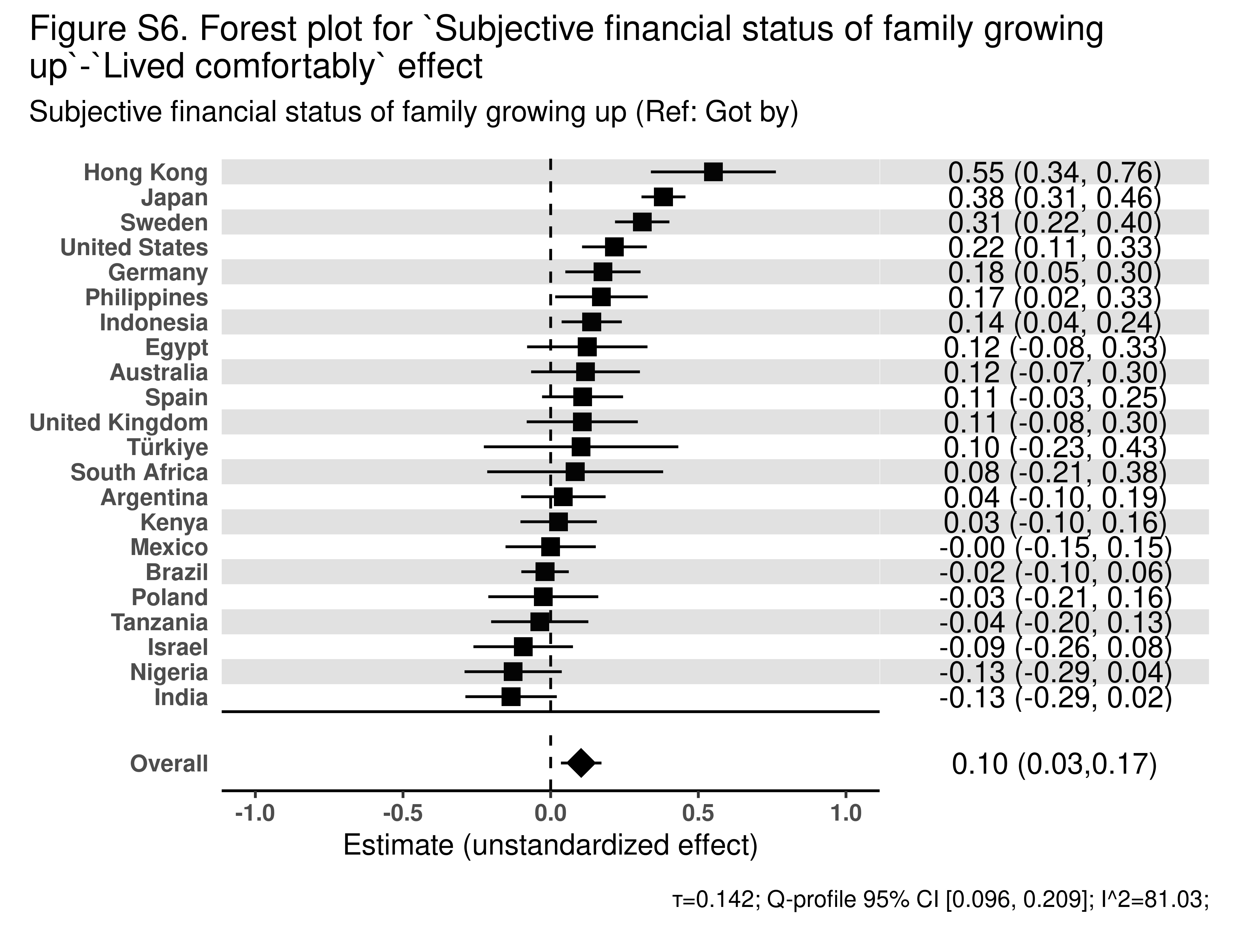

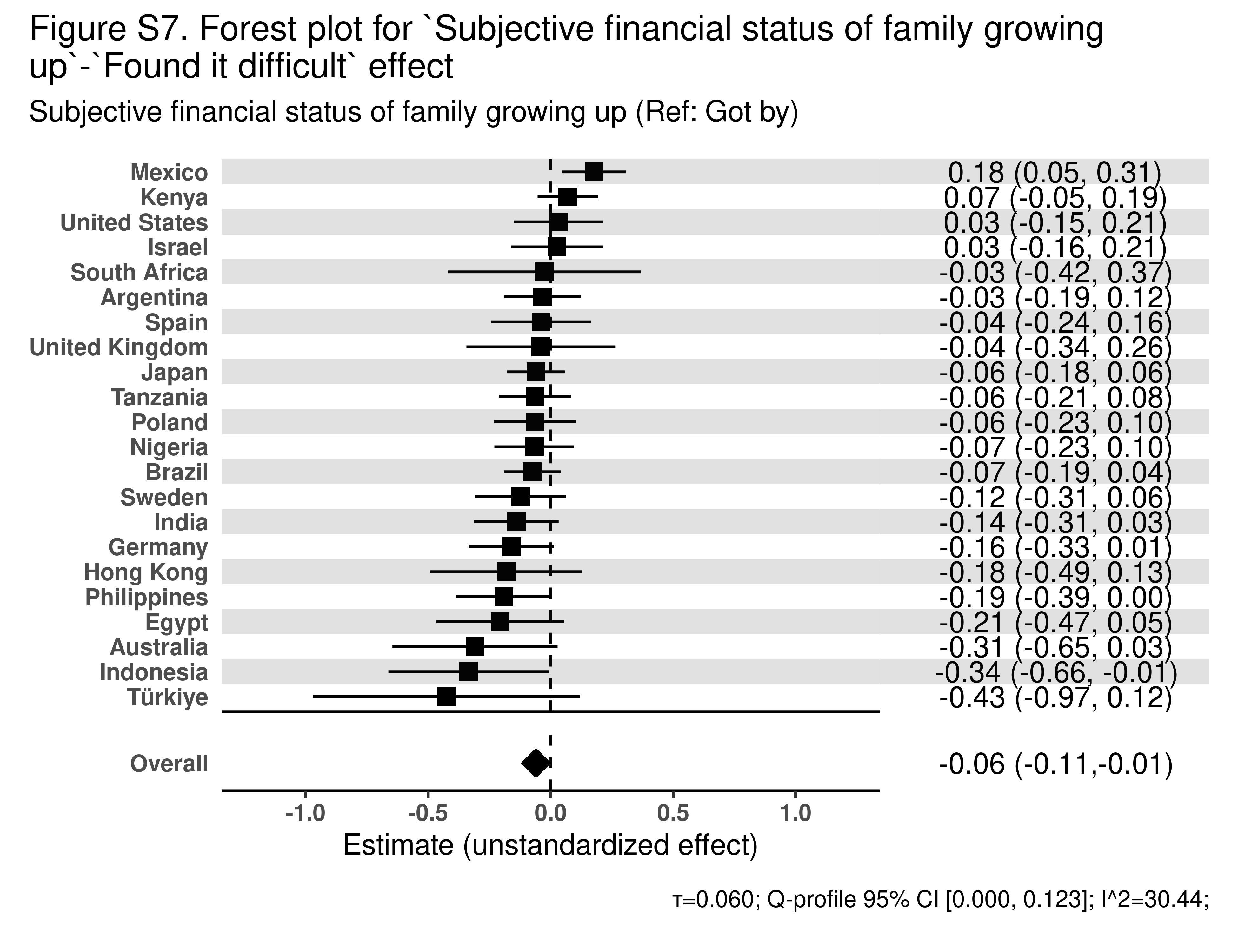

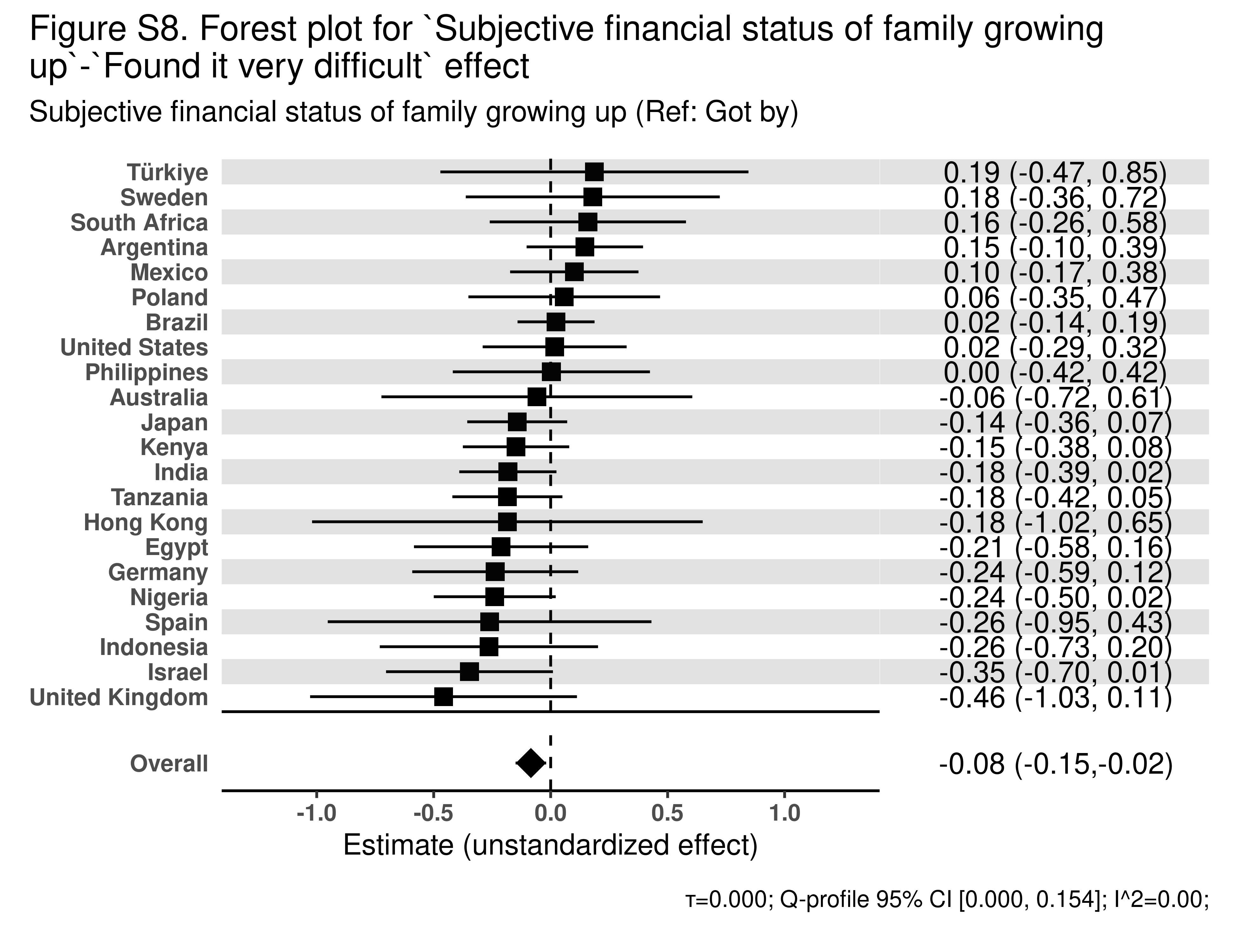

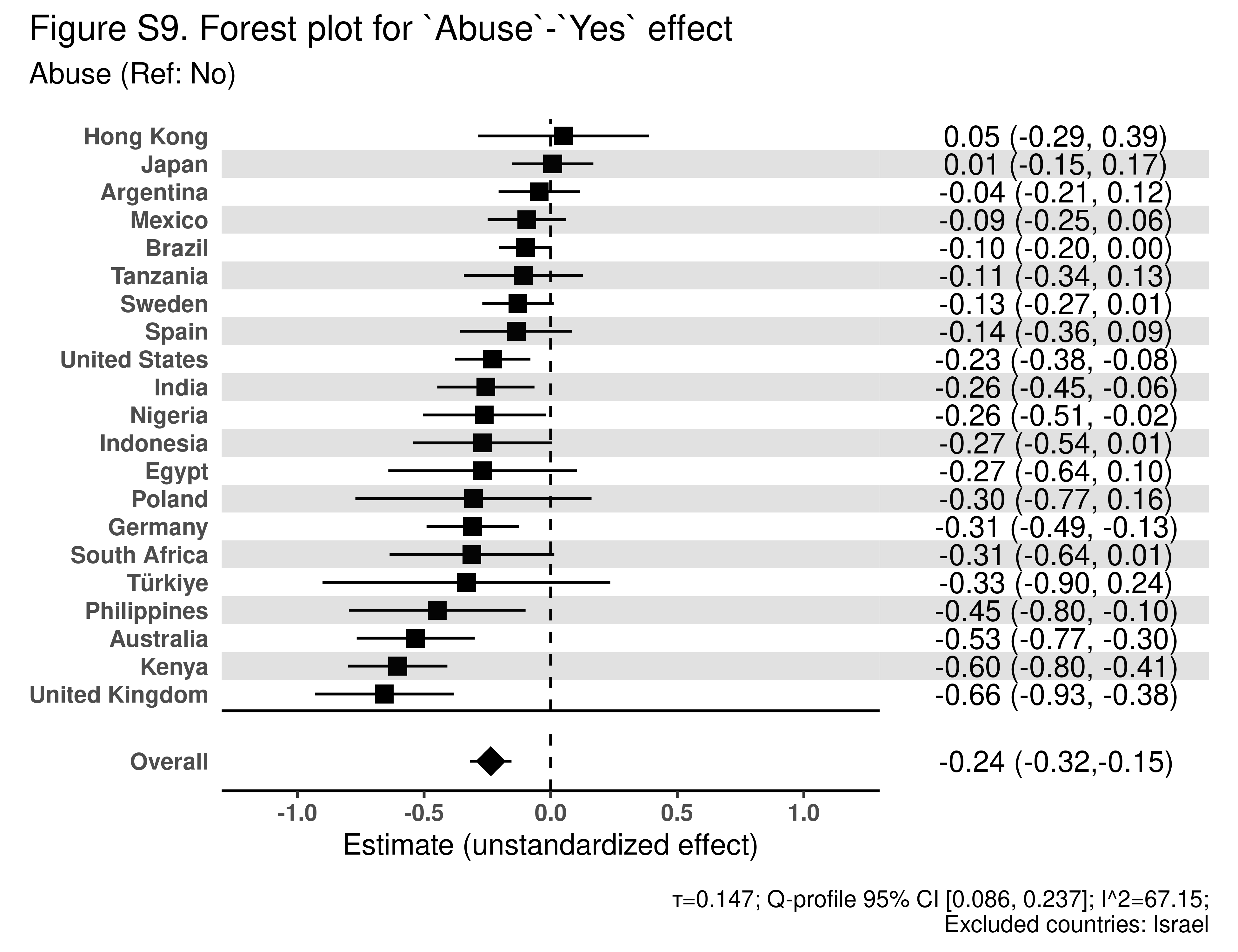

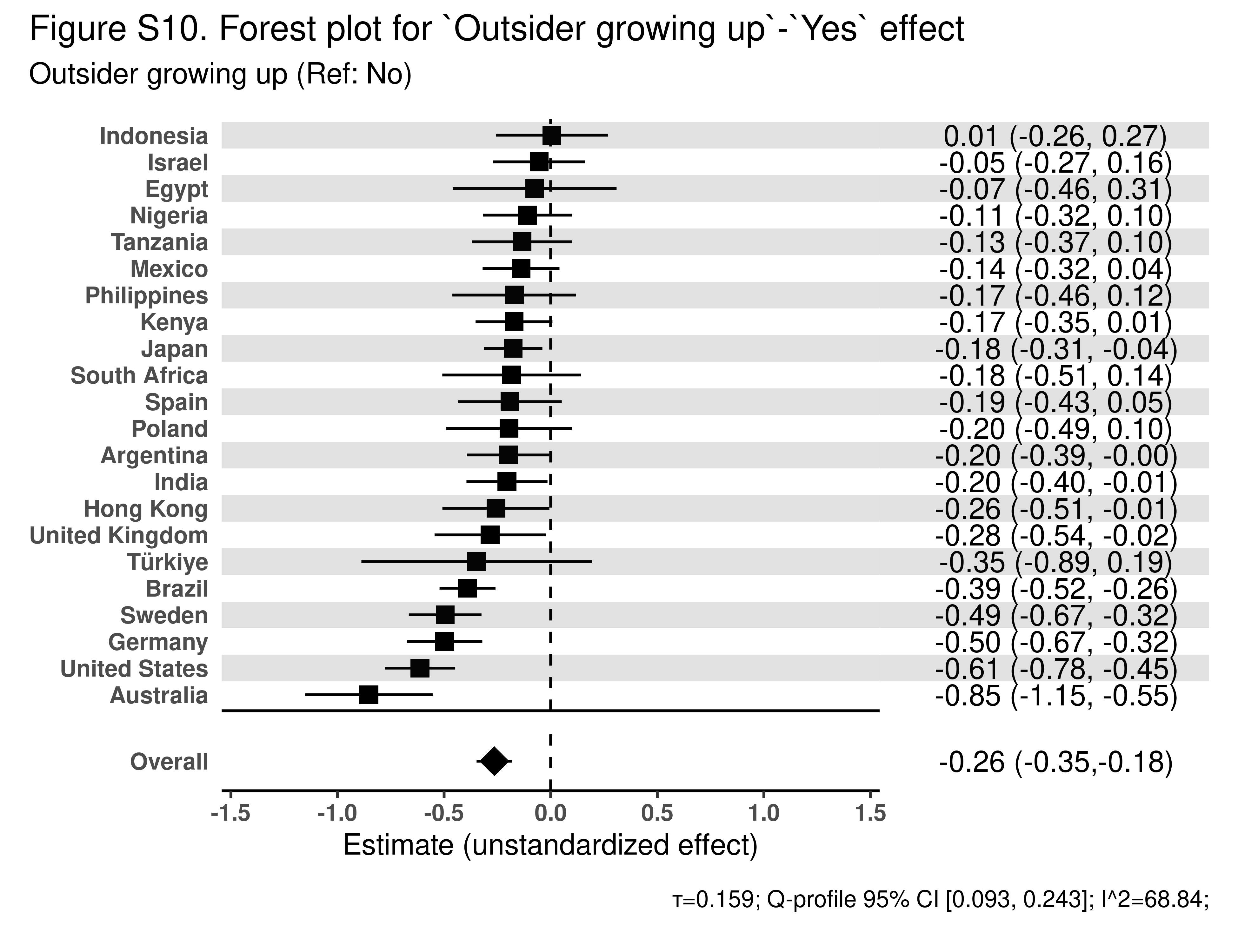

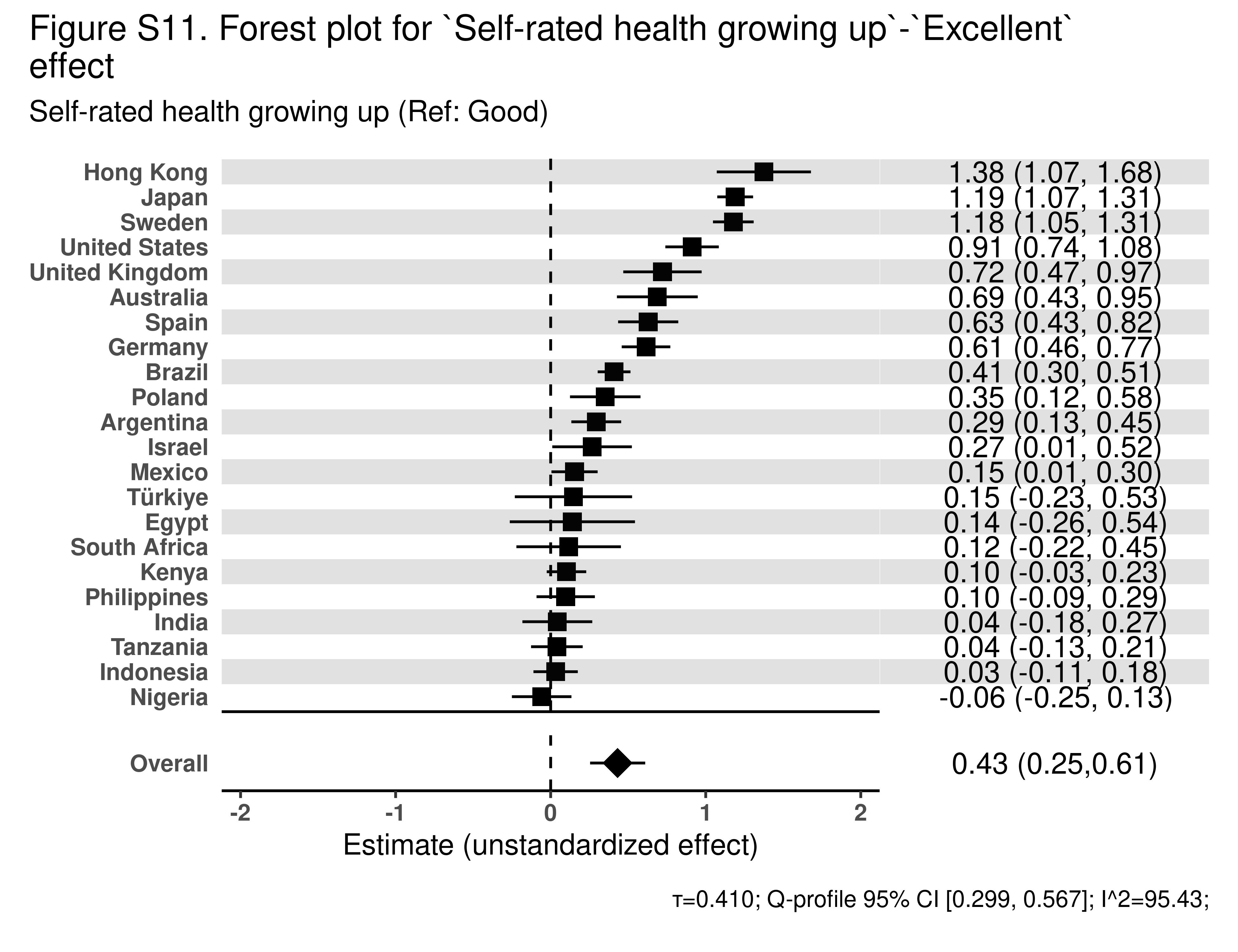

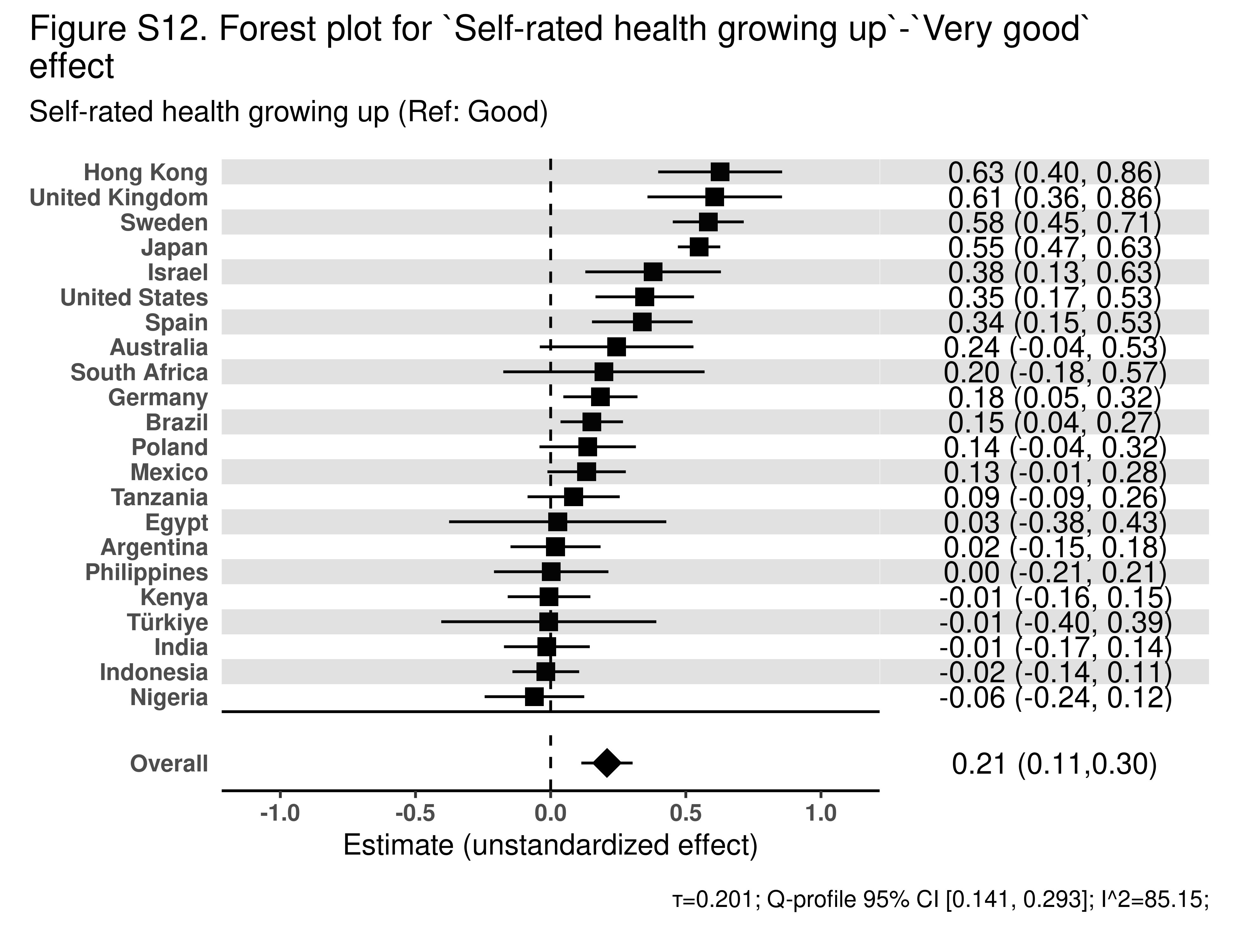

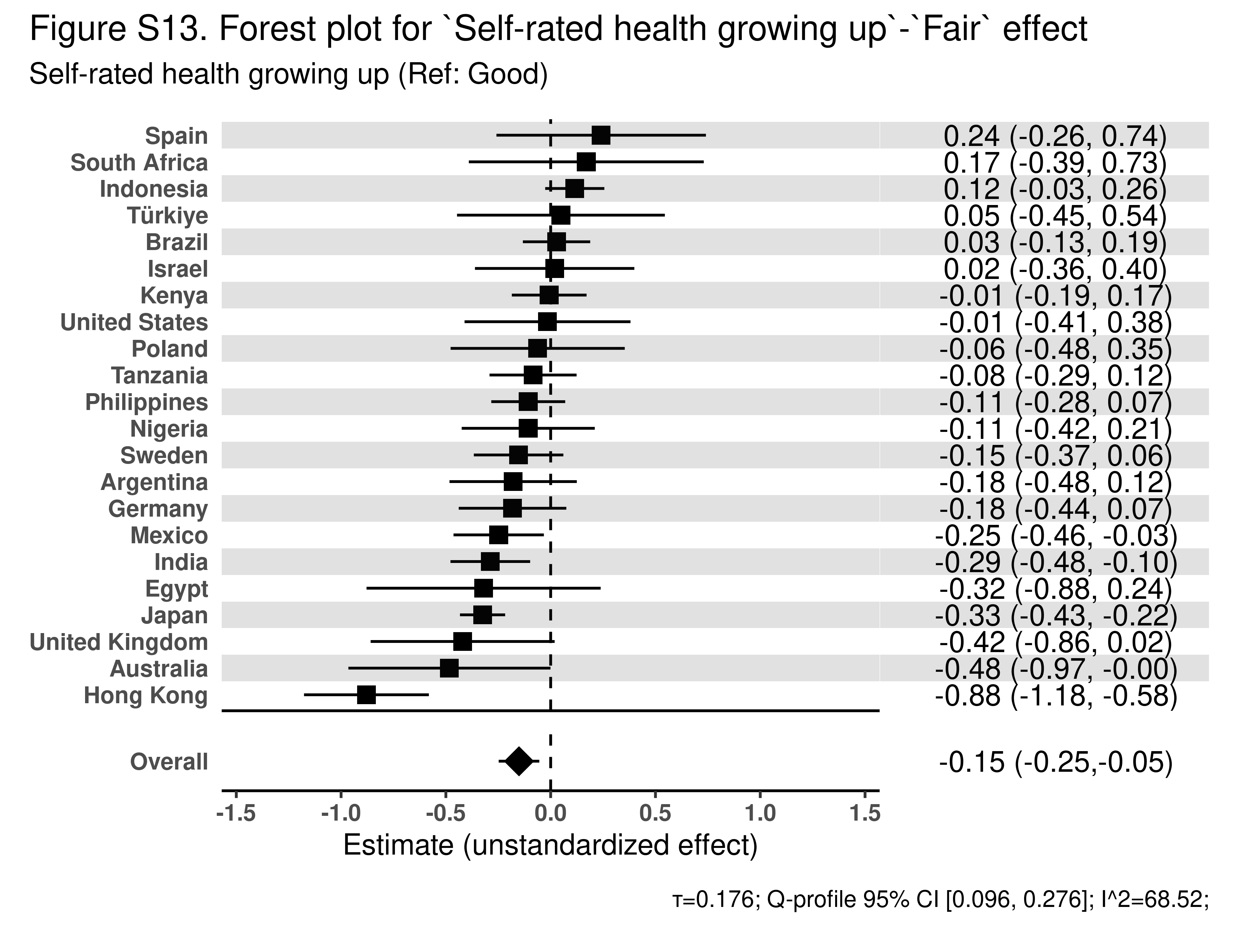

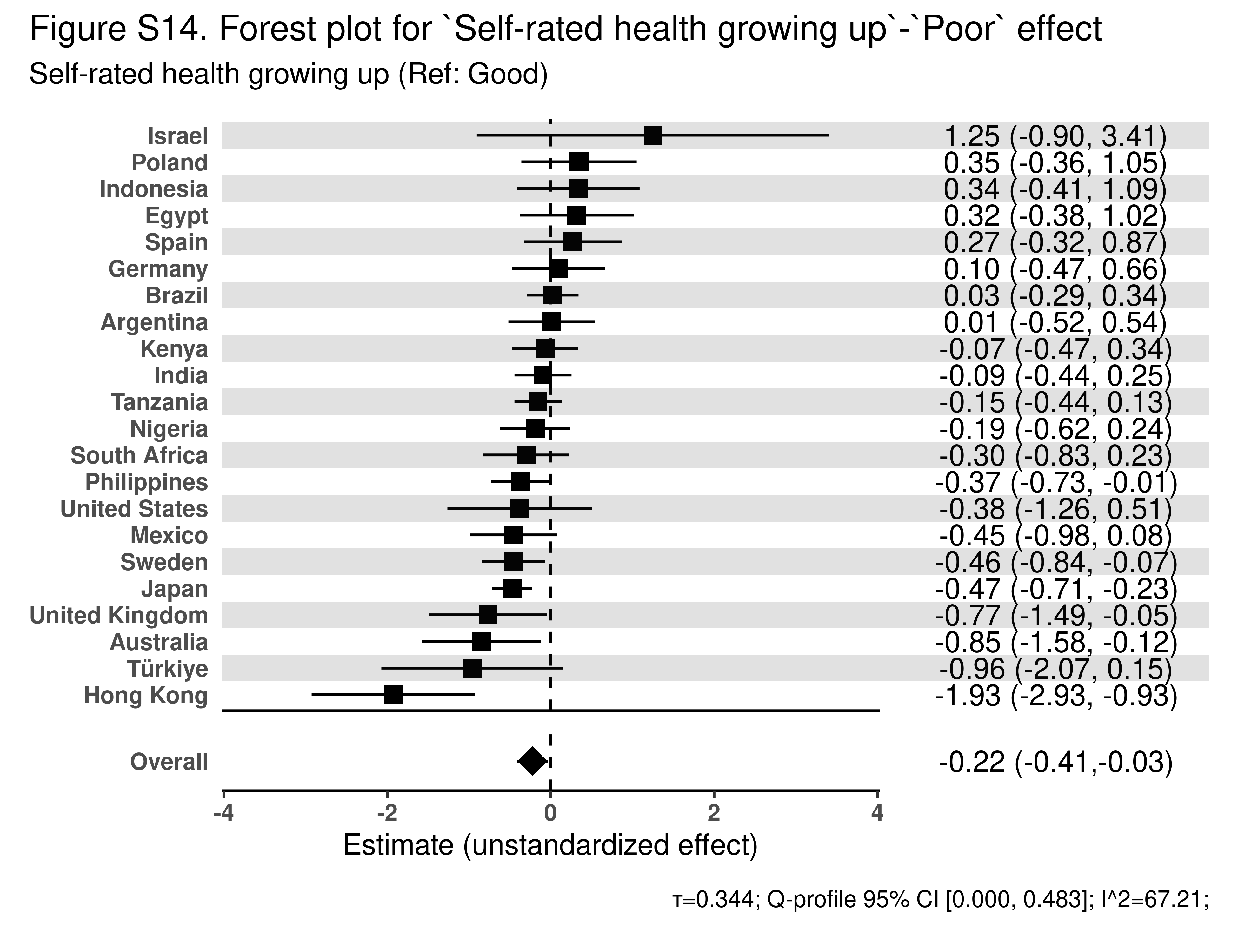

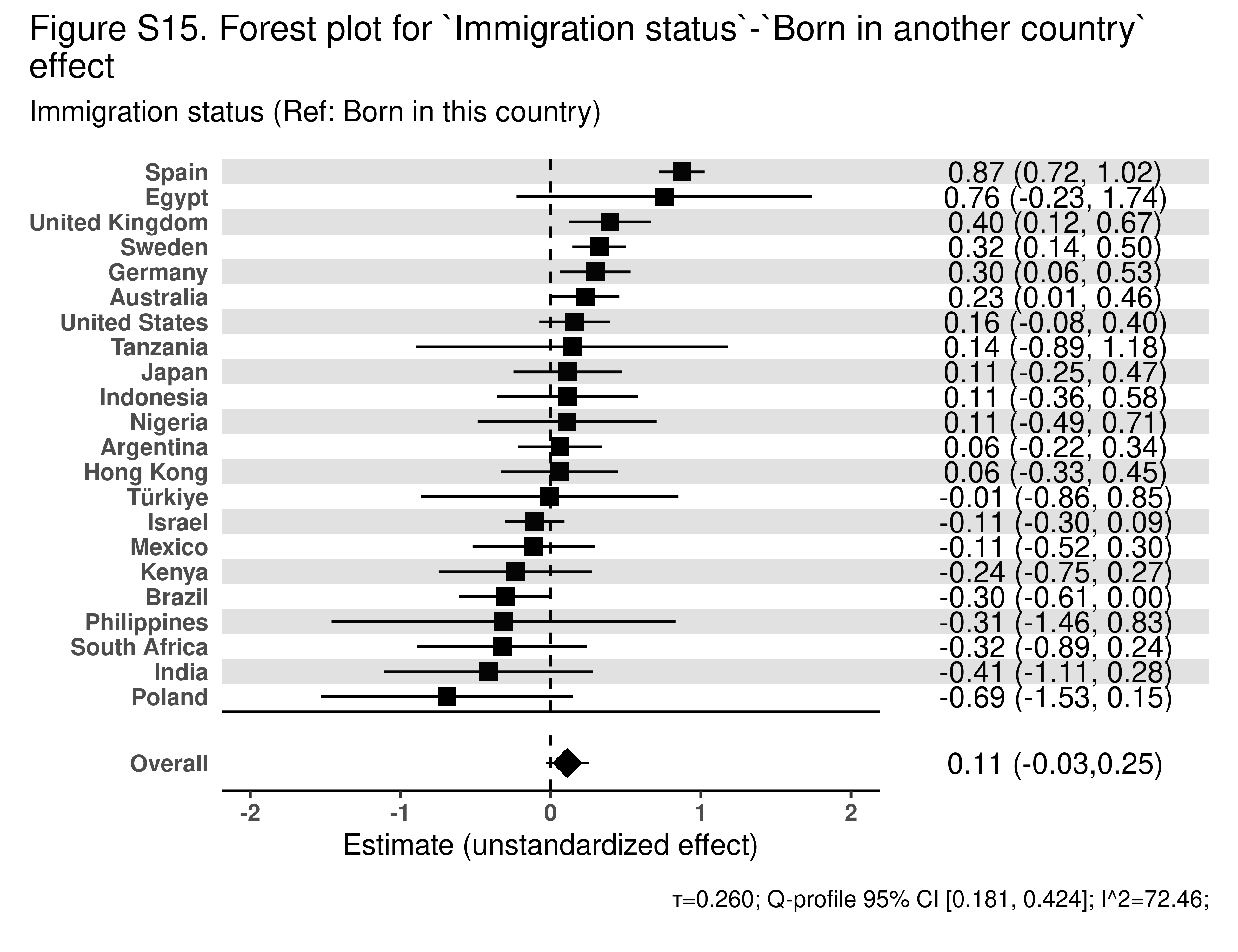

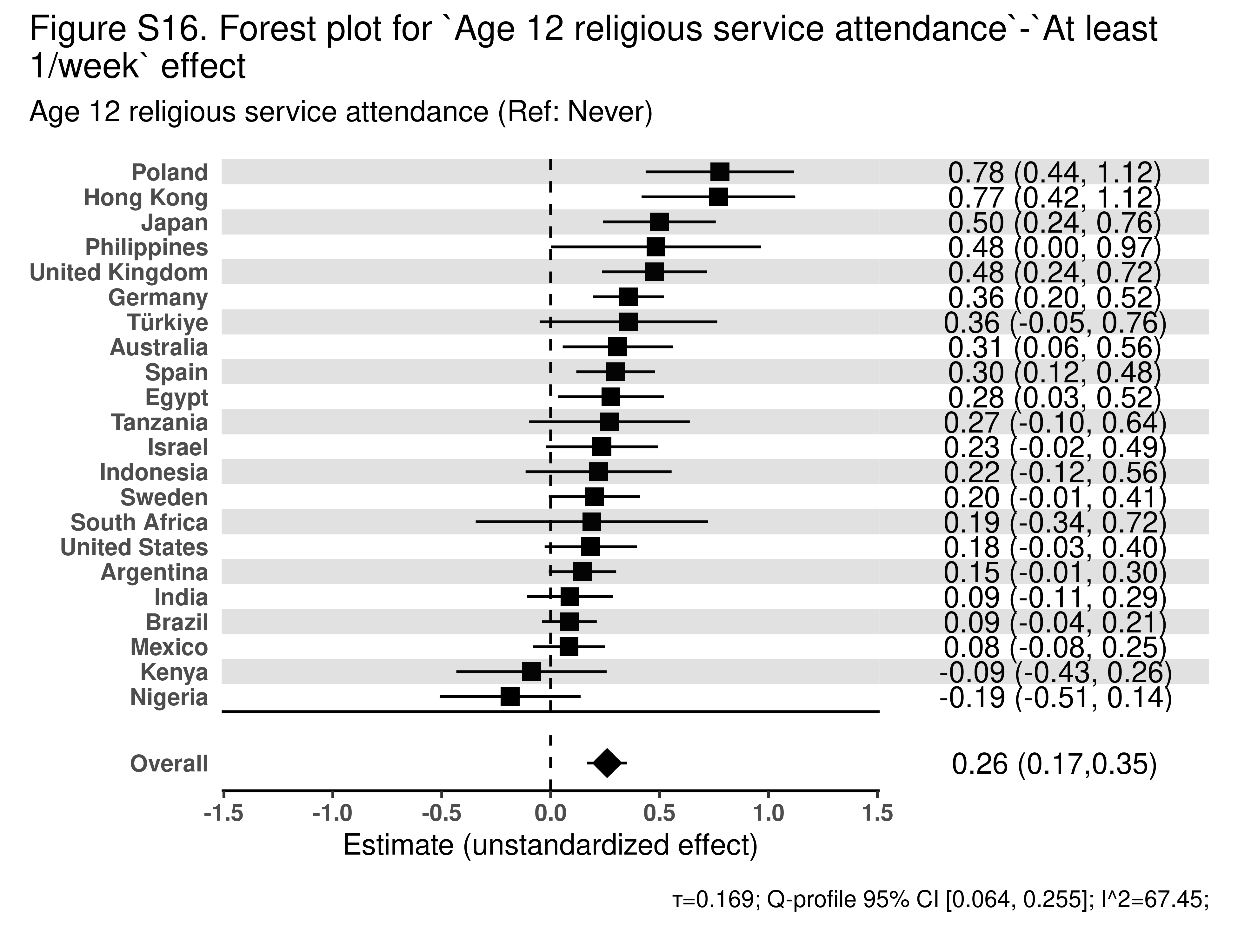

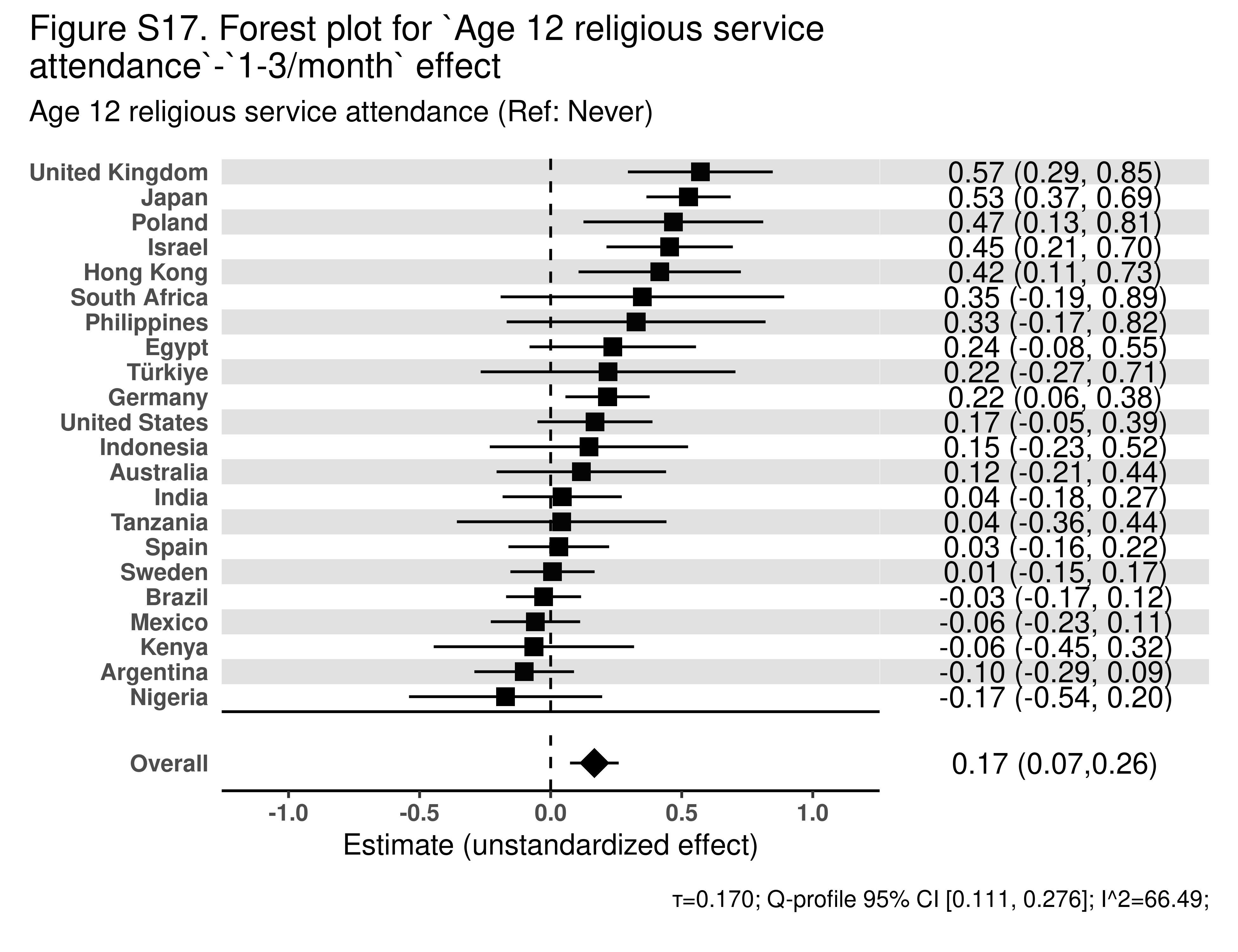

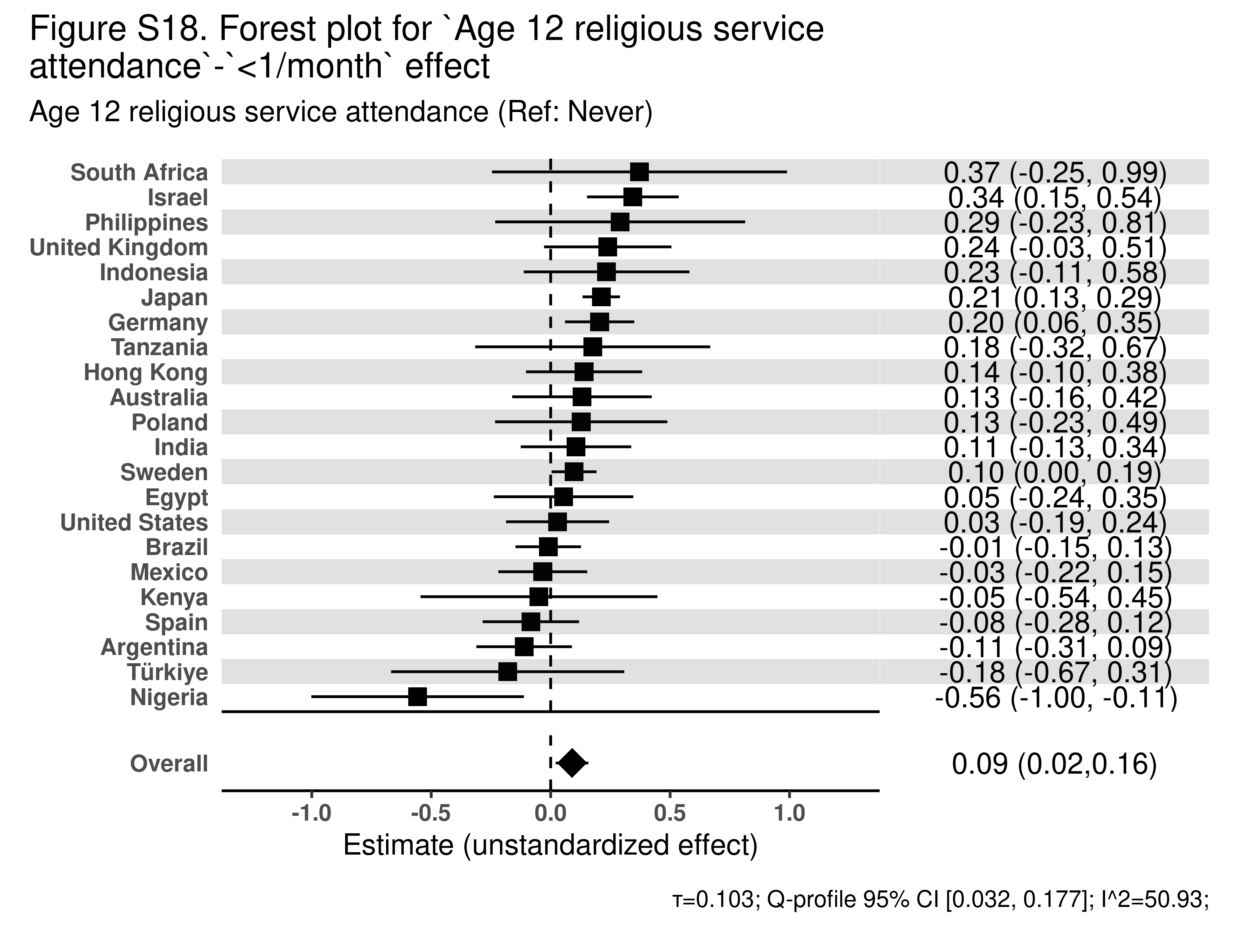

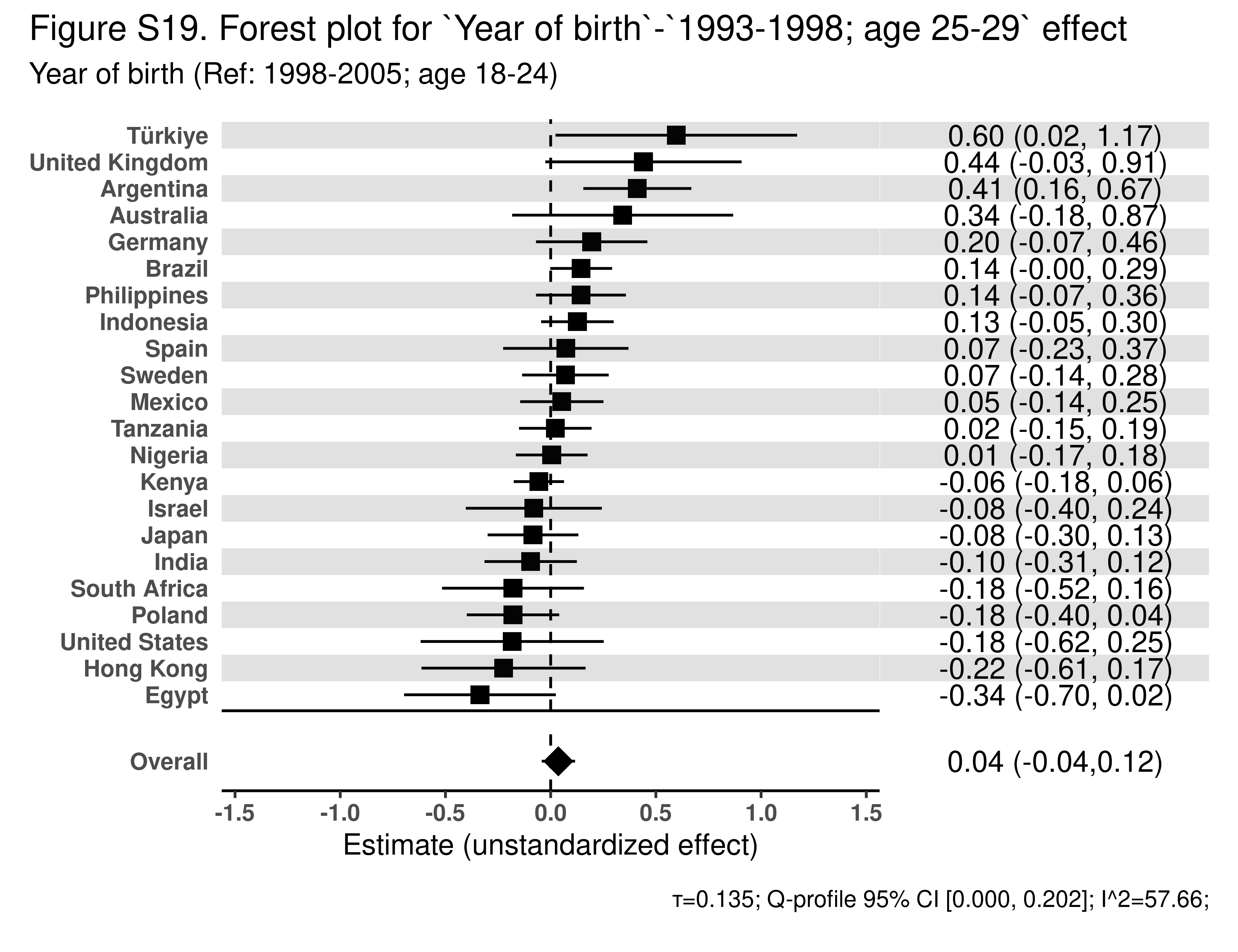

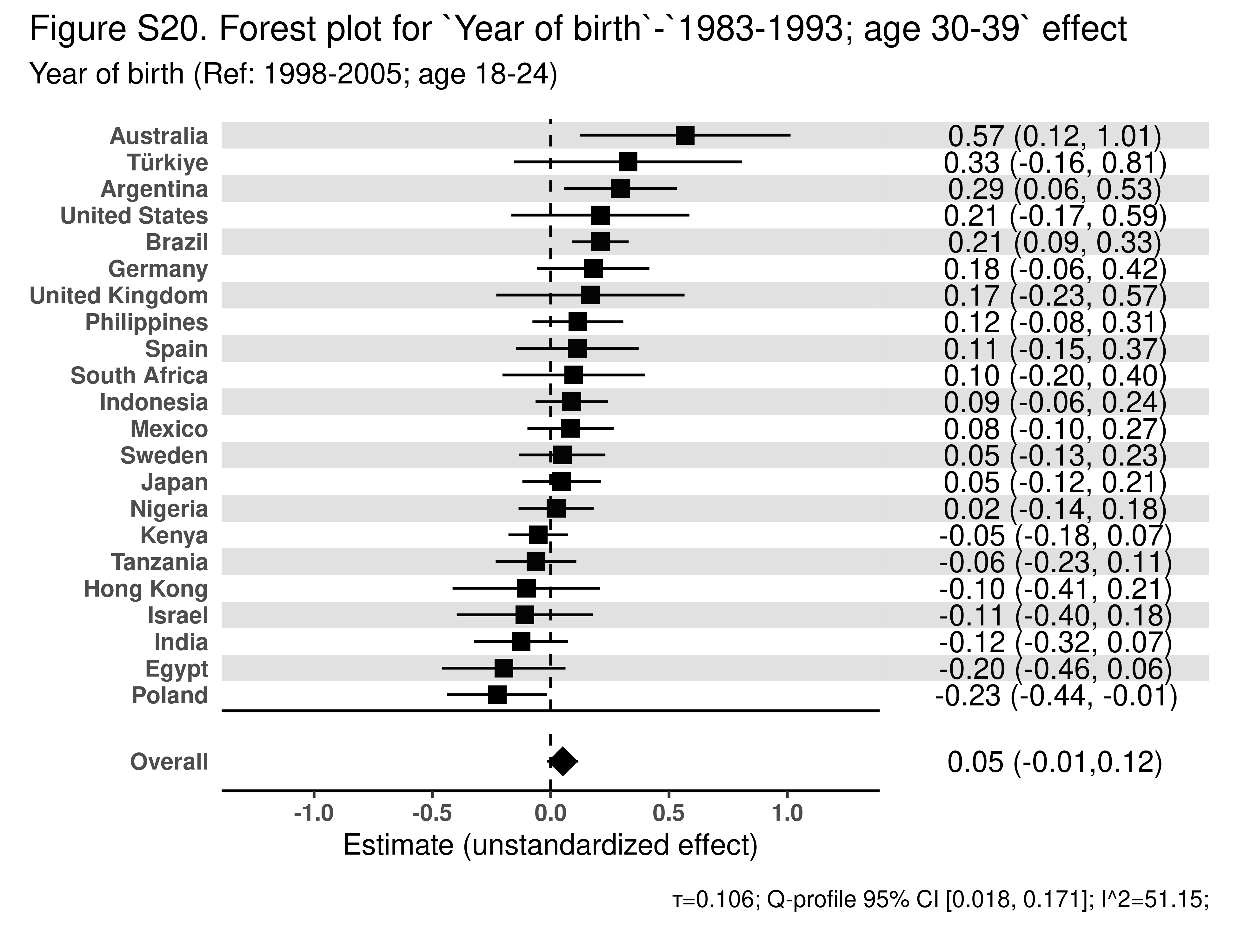

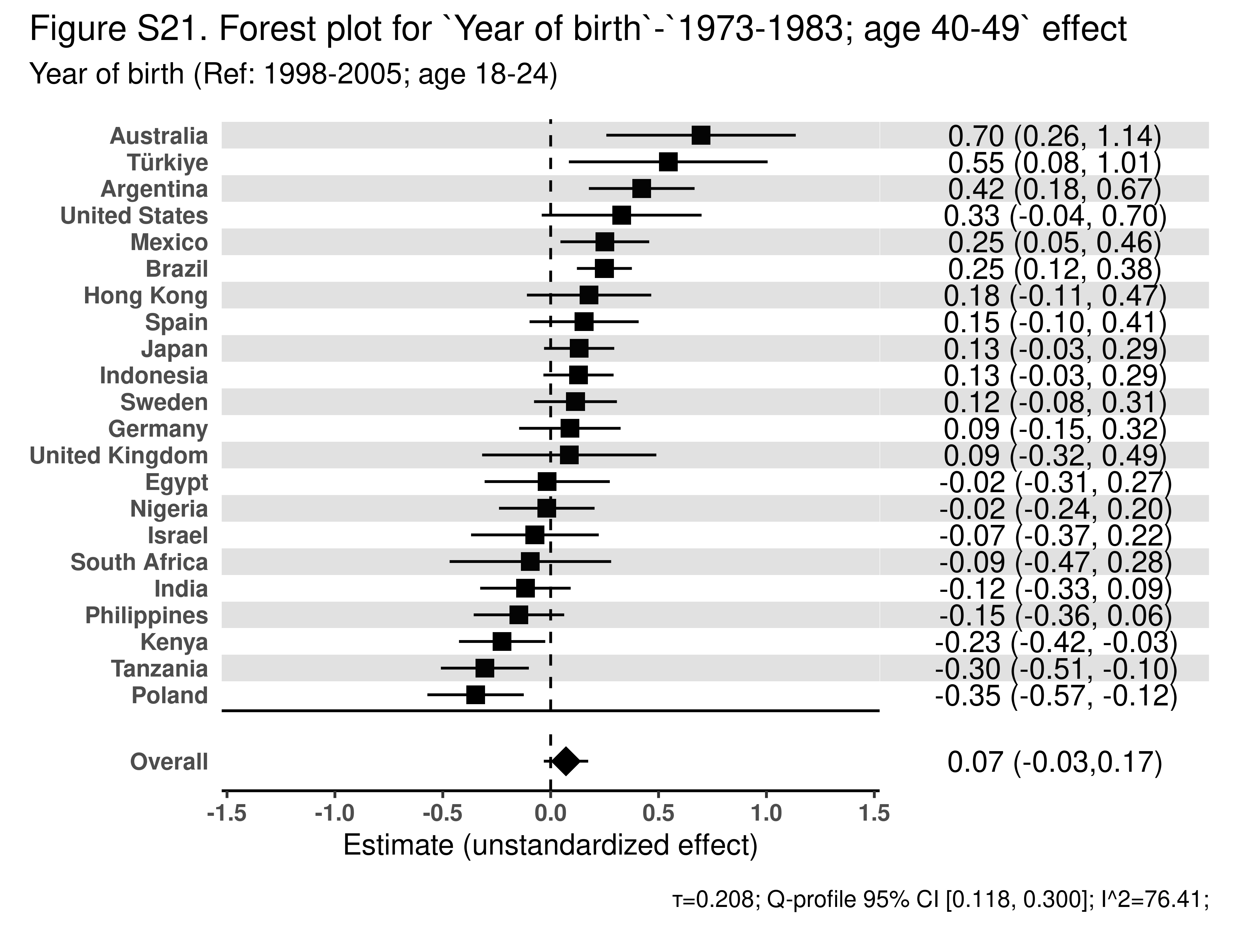

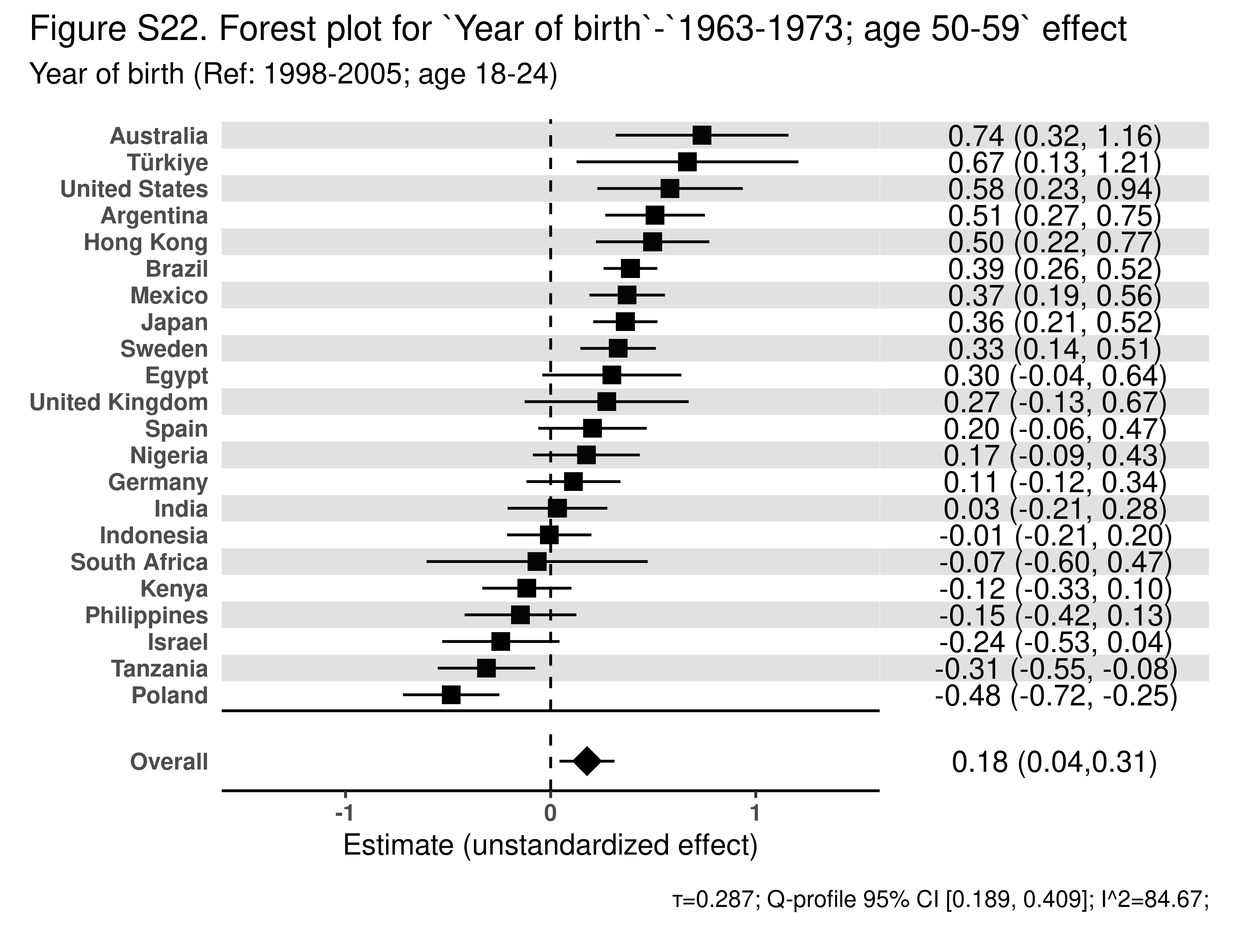

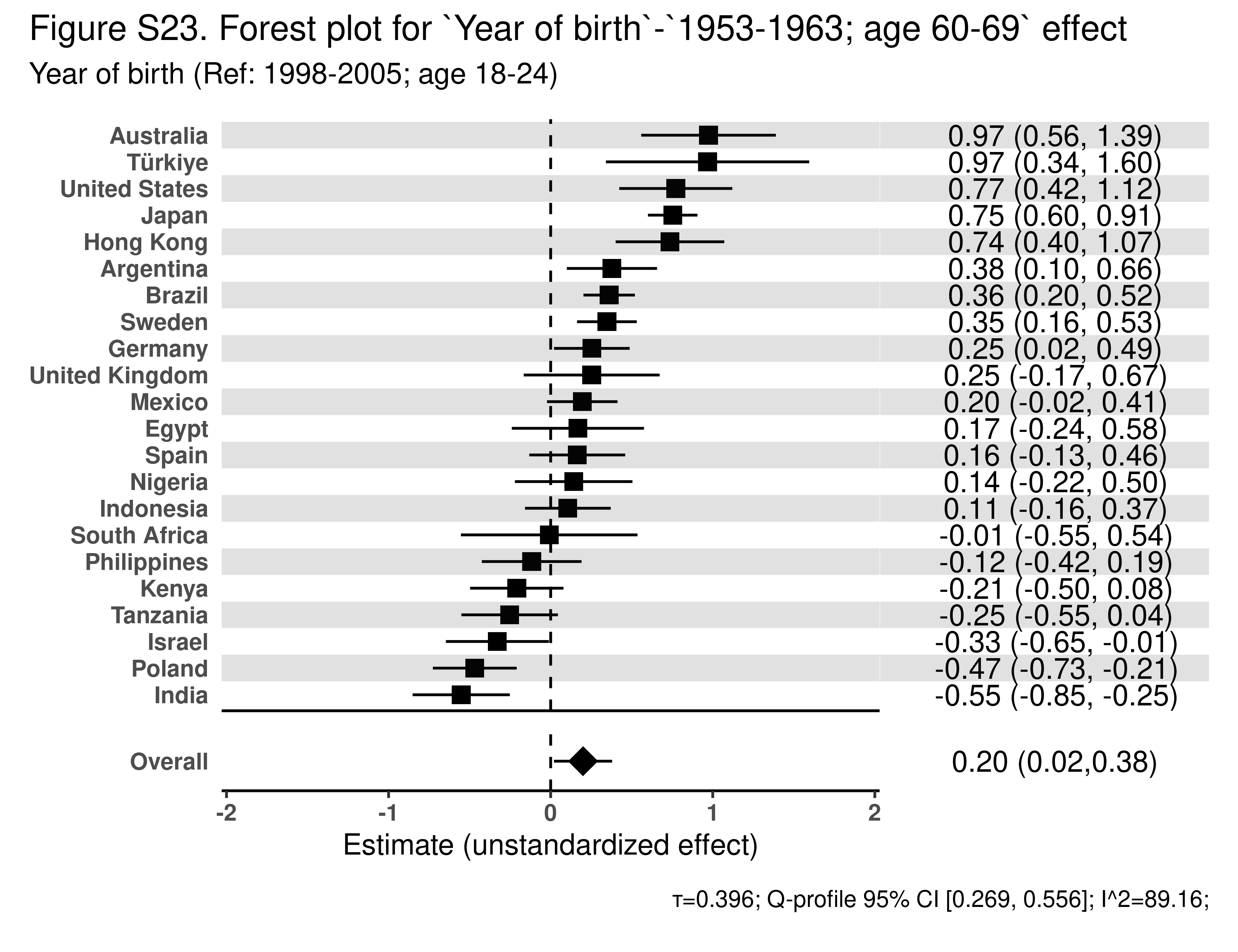

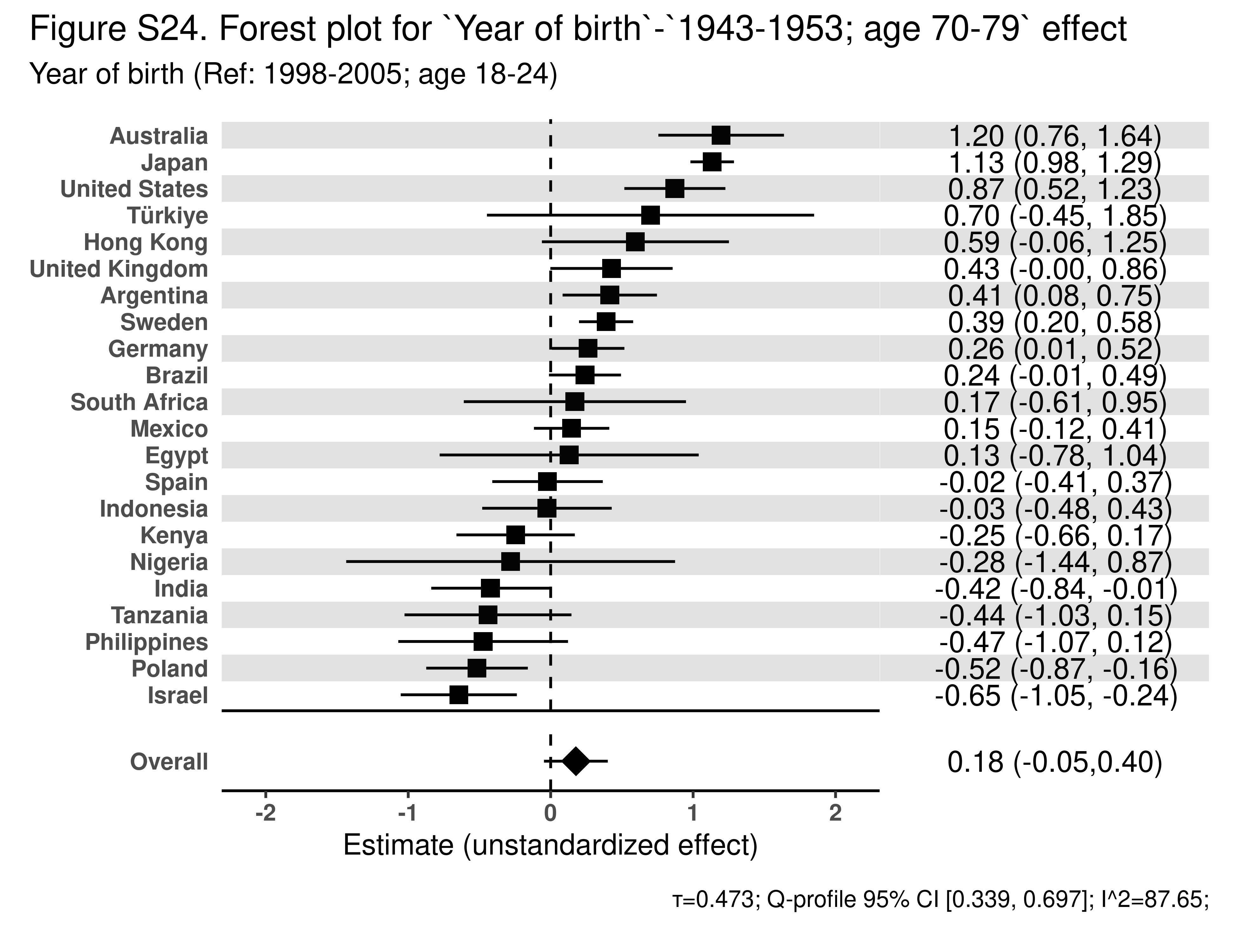

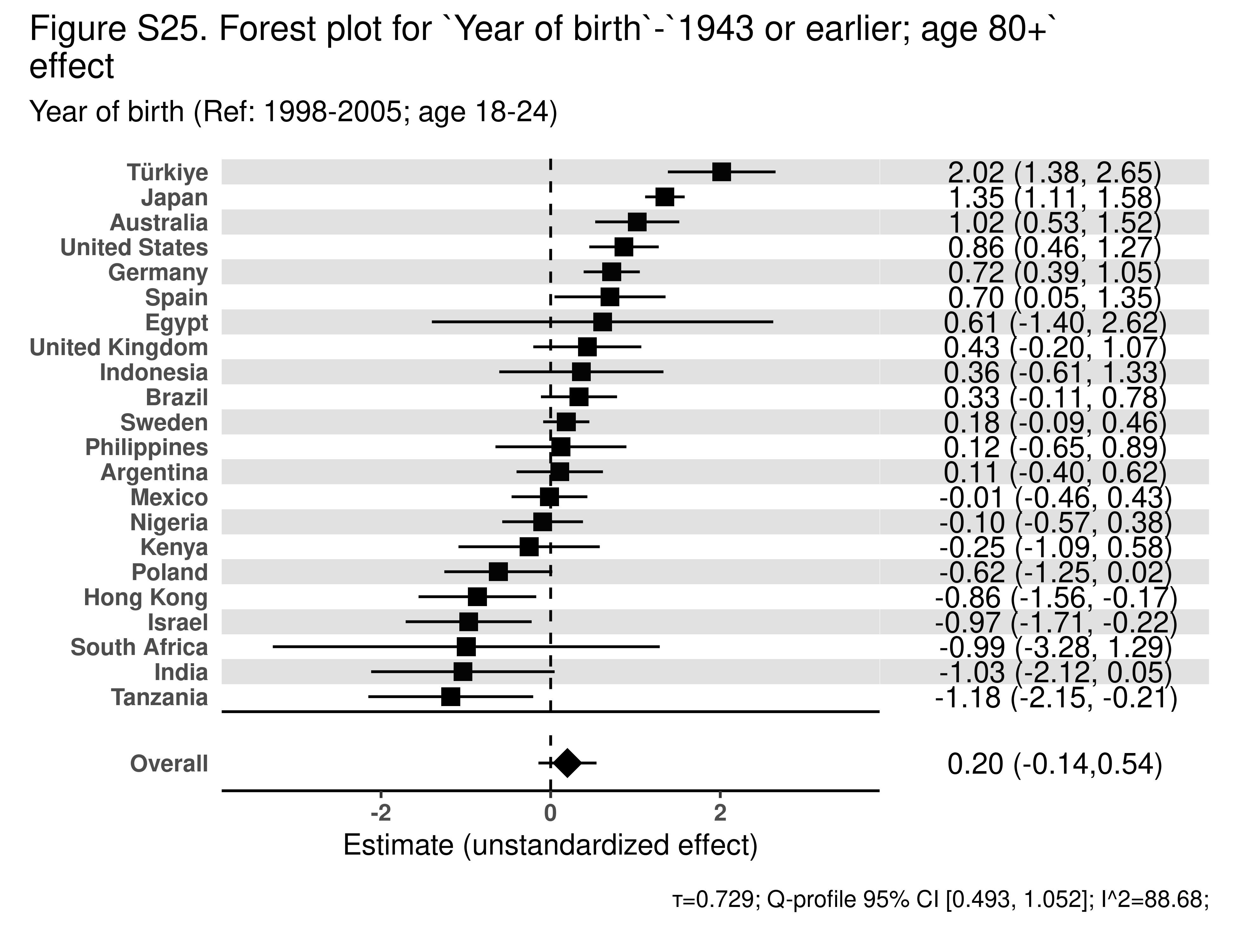

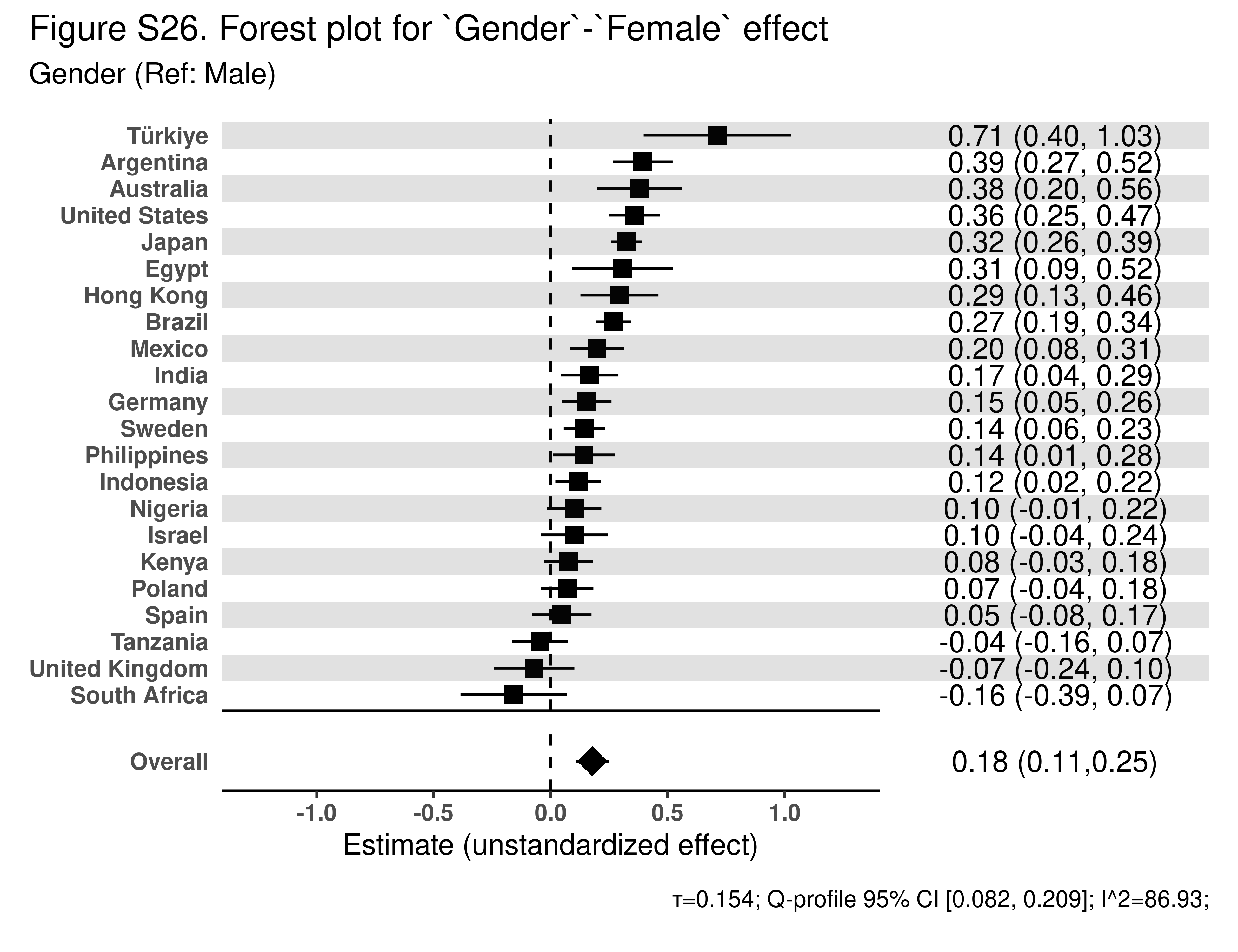

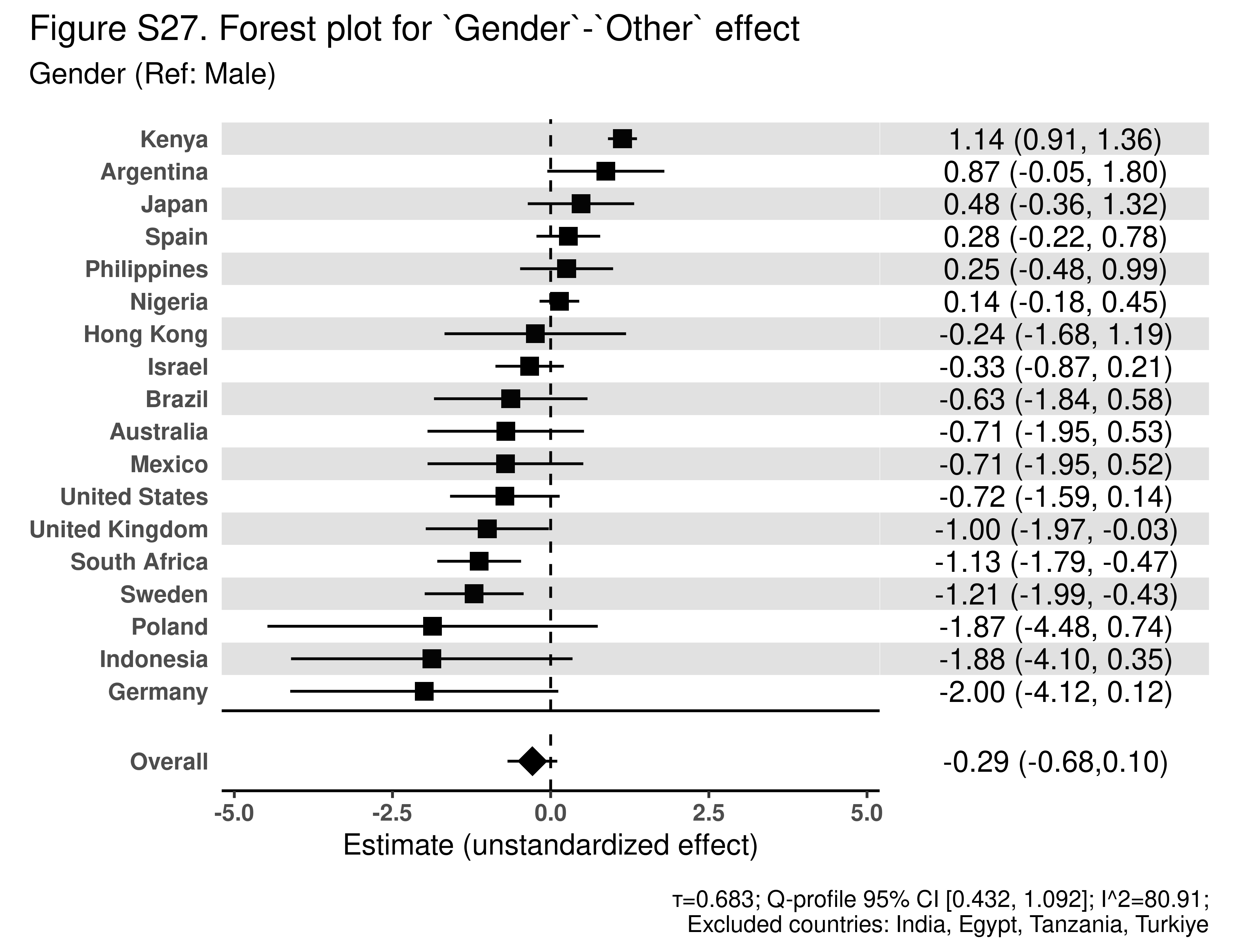
